# Triangulating molecular evidence to prioritise candidate causal genes at established atopic dermatitis loci

**DOI:** 10.1101/2020.11.30.20240838

**Authors:** Maria K Sobczyk, Tom G Richardson, Verena Zuber, Josine L Min, eQTLGen Consortium, BIOS Consortium, GoDMC, Tom R Gaunt, Lavinia Paternoster

## Abstract

**Background:** Genome-wide association studies for atopic dermatitis (AD, eczema) have identified 25 reproducible loci associated in populations of European descent. We attempt to prioritise candidate causal genes at these loci using a multifaceted bioinformatic approach and extensive molecular resources compiled into a novel pipeline: ADGAPP (Atopic Dermatitis GWAS Annotation & Prioritisation Pipeline).

**Methods:** We identified a comprehensive list of 103 accessible molecular resources for AD aetiology, including expression, protein and DNA methylation QTL datasets in skin or immune-relevant tissues. These were used to test for overlap with GWAS signals (including colocalisation testing where possible). This was combined with functional annotation based on regulatory variant prediction, and independent genomic features such as chromatin accessibility, promoter-enhancer interactions, splicing sites, non-coding RNA regions, differential expression studies involving eczema patients and fine-mapping of causal variants. For each gene at each locus, we condensed the evidence into a prioritisation score.

**Results:** Across the 25 AD loci investigated, we detected significant enrichment of genes with adaptive immune regulatory function and epidermal barrier formation among the top prioritised genes. At 8 loci, we were able to prioritise a single candidate gene (*IL6R, ADO, PRR5L, IL7R, ETS1, INPP5D, MDM1, TRAF3*). At a further 2 loci, 2 candidate genes emerge (*IL18R1/IL18RAP, LRRC32/EMSY*). For the majority of these, the prioritised gene has been previously proposed as a plausible candidate, but the evidence we combine here, strengthens the case for many of these. In addition, at 6 of the 25 loci, our ADGAPP analysis prioritises novel alternative candidates (*SLC22A5, IL2RA, MDM1, DEXI, ADO, STMN3*), highlighting the importance of this comprehensive approach.

**Conclusions:** Our ADGAPP analysis provides additional support for previously implicated genes at several AD GWAS loci, as well as evidence for plausible novel candidates at others. We highlight several genes with good/converging evidence of involvement in AD that represent potential new targets for drug discovery.

## Background

Defined by inflamed dry, hyperplastic eczematous skin and pruritus, atopic dermatitis (AD) is among the world’s top 50 common disease, with prevalence in 2010 estimated at close to 230 million cases and increasing[1]. AD is particularly common in childhood, with 10-16.5% prevalence in the first 5 years of life[2,3], 80% of which will then progress in the so-called atopic march and develop other allergic conditions[4]. In 2015, the burden of AD in terms of cost born by individuals, their families and society at large, was estimated at close to 5.3 billion dollars[5,6]. AD comorbidities include conditions such as: allergic rhinitis, asthma, allergic conjunctivitis, eosinophilic esophagitis and food allergy[7]. Due to the systemic immune nature of the disease, AD is also associated with increased risk for many inflammatory diseases: rheumatoid arthritis (RA), inflammatory bowel disease (IBD), systemic lupus erythematosus (SLE) and decreased risk for type 1 diabetes (T1D) [8,9].

AD is highly heritable - with estimates of up to 75% in twin studies[10]. The largest and most recent genome-wide association study (GWAS) of AD, undertaken by the EAGLE consortium in 2015 identified 25 loci (explaining 14.9% of the variance) that were associated with AD in individuals of European descent and replicated in an independent sample[11]. The majority of disease associated variants are located in non-coding regions. This observation implies that many disease-associated variants have a regulatory role rather than affecting protein function. It has now been widely recognised that integrating various biological data resources can provide complementary evidence about GWAS causal genes[12]. Across several disease it has been estimated that ∼50% of the time, the gene through which the causal SNP acts is not the closest gene to the index SNP[13,14].

The EAGLE GWAS utilised small eQTL resources (TwinsUK LCL and skin microarray samples[15]), fine-mapping tools (MANTRA[16]), variant annotation software (RegulomeDB[17] and HaploReg[18]) and two differential expression studies on eczema to investigate potential causal genes at the identified loci. Since 2015 there has been an explosion of new datasets from many cell types (such as eQTLGen[19], full GTEx release[20], Blueprint[21], GoDMC, promoter-enhancer HiC datasets) and new methods (such as finemap[22], JAM[23], fastPaintor[24], TWAS[25]) being made available that offer an opportunity to refine prioritisation of genes at the GWAS loci.

Several excellent computational approaches to aid identification of causal genes at GWAS loci exist, but these methods were not deemed suitable for our purpose as they either apply a single method across a large number of GWAS (such as SMR[26] and PhenomeXcan[27]) but do not integrate evidence from across many sources, are out of date (e.g. GWASrap[28], last updated in 2013), assemble a lot of data but do not show clear target prioritisation such as FUMA[29], INFERNO[30], or their source datasets are less relevant to our disease in terms of tissue selection because they target another specific disease, e.g. INQUISIT and breast cancer[31].

In this paper, we aim to comprehensively dissect AD GWAS loci by prioritising candidate causal genes and illuminating biological mechanisms through which candidate genes can impact AD risk. We applied a number of fine-mapping approaches and integrate evidence from across many sources relevant for skin disease. We developed ADGAPP (Atopic Dermatitis GWAS Annotation & Prioritisation Pipeline) which offers a consistent method of prioritising AD candidate genes and variants by devising a ranking system. ADGAPP which explicitly models our assumptions about the importance of different types of evidence as well as strength of the associations relating the features to genes and variants.

In our GWAS target prioritisation pipeline, we use methods (coloc, TWAS) to formally compare the association patterns in QTL studies and GWAS to test for their colocalisation whenever full summary statistic are available, as ∼50% of common variants are associated with one eQTL or more in GTEx[32] so simple lookups for variant overlap alone will result in many false positives. We also integrate the results of two independent pipelines: enrichment-based regfm[33] and network-based PrixFixe[34] for GWAS-wide prioritisation of target genes.

We aimed to select relevant datasets to reflect the full molecular aetiology of AD. Broadly speaking, AD pathogenesis is thought to stem from epidermal barrier disfunction, as well as abnormal immune system activation[35]. In addition to skin lesions, AD abnormalities are also seen in non-lesional skin and blood[36,37]. Therefore, we made it a priority to include molecular QTLs from skin, blood and immune-specific cell types in ADGAPP. Direct importance in AD physiology is evaluated by including differential gene expression and DNA methylation studies and proteome comparisons involving eczema patients.

As the lead SNPs identified in the initial GWAS analysis may not be causal[38], we use three Bayesian fine-mapping algorithms to initially prioritise causal SNPs.∼90% of index SNPs are placed in the non-coding regions of the genome, and so causal variants are expected to have mainly regulatory functions[18]. In order to include the functional potential of each variant in our ranking, we directly look up overlap of potential GWAS causal variants with experimentally generated functional annotations as well as predictions for regulatory impact of the variant generated by machine learning models. Integration with high-throughput chromatin conformation capture (3C) data allow us to check if our variants of interest are located in enhancer regions or promoters of genes highlighted in eQTL analysis in the matching tissue.

We integrate several established fine-mapping and gene prioritisation methods in a unique AD-focused gene prioritisation pipeline to comprehensively evaluate the causal genetic evidence at each locus and utilise an exhaustive set of AD-related molecular datasets to best support these methods. Our pipeline in combining these methods generates a global score, which can be used to assess the magnitude of evidence for (and relative evidence between) tested genes as the causal gene at a particular locus. Such a score can serve as a metric which allows rapid gene prioritisation by molecular biologists and other interested parties, such as pharmaceutical companies.

## Methods

### Source GWAS

Paternoster et al. (2015)[11] is the biggest GWAS on AD to date, consisting of EAGLE Consortium data of 21,399 cases and 95,464 controls from populations of mostly European, but also African, Japanese and Latino ancestry. In our analysis, we investigate 25 loci, which show either genome-wide significance and for novel loci are replicated in independent European ancestry sample (21 loci), or are significant loci prioritized by the MAGENTA gene set enrichment analysis[39] presented in the original paper (4 loci)[11].

### Bayesian fine-mapping

To identify likely causal genetic variants in the regions harbouring AD GWAS signals, using only information on association strength and LD structure, we used three different Bayesian fine-mapping methods: Finemap[22], fastPaintor[24] and JAM[23]. Each method relies on different prior assumptions and model formulation.

As input data we used, the association statistics from the AD GWAS in individuals of European ancestry, published in Paternoster et al. (2015). For LD structure, in Finemap and Paintor analysis, we used *r* correlations calculated from the 1000 Genomes[40] European reference (*n*=503), as it allowed inclusion of more high confidence SNP calls compared to UK Biobank[41] - 9,265,840 versus 8,391,826. UK Biobank panel[41] filtered for European ancestry (*n*=48,167) with standard QC applied[42] was used as LD reference panel in JAM analysis. This was because JAM requires a high number of individuals in the reference and removal of highly correlated SNPs for the genotype matrix to be invertible. To that end, we also pruned SNPs prior to feeding them to JAM. We used a threshold of r^2^ > 0.95 in Priority Pruner (http://prioritypruner.sourceforge.net/) and set minor allele frequency (MAF) threshold at 0.05 and minimum SNP call rate of 0.9, while force selecting all the index SNPs.

Our fine-mapping integration protocol involved running all the 3 programs with all the SNPs within the interval of 10kbp, 100kbp, 500kbp, 1Mbp, 3Mbp centred on the index SNP. We also used r^2^ -based and D’-based haploblock intervals defined with the BigLD[43] and Gpart[44] algorithms, respectively, in 1000 Genome EUR panel.

We ran our Finemap analysis using shotgun stochastic search, whereas for Paintor, we varied the algorithm from exact exhaustive search to MCMC when considering from up to 2 or max. 5 causal SNPs in the region, accordingly. Maximum number of algorithm iterations was set at 1000 in Paintor and 10 million in JAM. In order to permit analysis of a binary trait in JAM, linear mapping of log-odds ratios was performed[45] and the residual variance inverse gamma hyperpriors were set to “GaussianResidualVarianceInvGammaPrior_a” = 2, “GaussianResidualVarianceInvGammaPrior_b” = proportion of cases * (1-proportion of cases).

When comparing output of Finemap, Paintor and JAM we only considered top fine-mapped SNPs with Bayes Factor > 100 and posterior probability of being causal of at least 0.1.

### Variant filtering

In subsequent gene analyses, shown below, we limited ourselves to SNPs within the region in significant LD with the index SNP in 1000 Genomes EUR population, henceforth referred to as the GWAS locus interval. The region in each case was defined by the furthest removed SNP with r^2^ >= 0.2 within 1 Mbp interval centred on the index SNP and all the SNPs within the boundary were considered. We confirmed that we captured all the SNPs within the broadly defined haploblock by re-defining the boundaries based on maximum 3 Mbp interval. Definition of our haploblock changed only in the case of 3 index SNPs: however in all cases there was a stretch of at least 100 kbp (rs61813875 with a very sharp LD decay and rs41293864 situated in the MHC region with complex LD), up to 350 kbp (rs77714197) of SNPs with no r^2^ > 0.2 so we dismissed those as outliers and used the 1 Mbp interval-defined region in all cases.

### Identification of key tissues and cell types

In order to focus on key tissues/cell types associated with eczema variants, we used gene set enrichment in SNPSea[46] with the supplied gene expression datasets: Gene Atlas Affymetrix expression in 79 human tissues[47], Immunological Genome Project[48] Affymetrix expression in 249 murine blood cell types and FANTOM CAGE[49] in 533 human cell types. We ran SNPSea using recommended settings and used index SNPs as input. SNPsea considers genes in LD with the index SNPs and for each locus and cell-type combination, selects one gene which shows highest tissue-specificity of expression. The merged gene set across loci is then scored for its cell-type specificity and the resulting score compared to the ones obtained from the null distribution of results for random SNP sets matched on the number of genes in LD in order to obtain a permutation *p*-value.

Secondly, we used MAGMA[50] gene enrichment analysis on GTEx 7.0[20] data as carried out by FUMA[29]. Briefly, MAGMA gene-based analysis was done using SNPs in the locus interval to identify all the genes associated with our GWAS hits, and subsequently MAGMA gene-property analysis was applied to test tissue type specificity in expression of identified genes.

### TwinsUK eQTL identification

We used genotype array data and RPKM -normalised expression in lymphoblastoid cell line (LCL) and skin tissue for females in the TwinsUK cohort[51]. RPKM values were rank-transformed to normality using GenABEL[52] R package before eQTL mapping. *cis*-eQTLs 1.5 Mbp upstream and downstream of TSS were identified using linear mixed model implemented in GEMMA[53]. We used age as covariate in the analysis involving all samples and centred relatedness matrix as random effects. PEER analysis was run to identify any additional hidden covariates not captured above[54]. eQTL associations were identified using the Wald test.

### CEDAR eQTL re-identification

In the analysis involving the CEDAR cohort (e.g. Momozawa et al. (2018)[55]), we used the publicly available data: imputed genotypes and normalized gene expression values from blood and intestinal cell types (CD4^+^ T lymphocytes, CD8^+^ T lymphocytes, CD19^+^ B lymphocytes, CD14^+^ monocytes, CD15^+^ granulocytes, platelets, ileum, colon, rectum) adjusted for 4 top PCs and covariates (sex, age, smoking status, batch). We used GEMMA’s linear mixed model and Wald test to re-identify *cis*-eQTLs within 1.5 Mbp upstream and downstream of TSS.

### Colocalisation with coloc and TWAS

We obtained full summary statistic results for cis-eQTLs detected in whole blood in the eQTLGen dataset[56] – accessed on 08/08/2018, eQTLs from GTEx ver. 7 dataset identified in the following tissues: whole blood, spleen, sun-exposed and unexposed skin, transformed fibroblasts and EBV-transformed lymphocytes[57], eQTLs published from the Kim-Hellmuth et al. (2017) study investigating monocyte response to microbe-associated molecular patterns[58], eQTLs in the monocytes, neutrophils and CD4+ T cells from the BLUEPRINT project[21], and pQTLs from whole blood in the Sun *et al*. (2018) dataset[59] as well as TwinsUK and CEDAR eQTLs identified above (Dataset S1). Subsequently, colocalisation signal between betas from GWAS and eQTLs/pQTLs for genes within 1.5Mbp upstream and downstream of index SNP was evaluated with the coloc[60] R package. In addition, three-way colocalisation of GWAS and whole blood molecular phenotypes: pQTLs and eQTLs was investigated with moloc[61] with default priors but brought no significant results. In coloc analysis, we considered loci with posterior probability of H4 (PPH4) > 0.5 as informative enough to be included in ADGAPP, as done previously[62]; with H4 stating the hypothesis of both traits being associated and sharing a single causal variant.

We also carried out a TWAS[25] analysis, where reference datasets with gene expression and genotype data (GTEx ver. 7.0, CEDAR and TwinsUK) were used to predict gene expression in our target GWAS. We used 100 permutations to conservatively calibrate the imputed gene expression association statistic conditional on the GWAS strength of association and used provided European1000 Genomes panel for LD reference. Any significant gene expression associations with AD were then post-processed to identify conditionally independent associations. In addition, coloc analysis was carried out, based on marginal TWAS weights with provided scripts.

The analysis pipeline for the SMR analysis has been described previously[26]. In brief, eQTL datasets from the eQTLGen consortium[19], BLUEPRINT[21] across 3 cell types (monocytes, neutrophils and T cells) and GTEx ver. 7[20] across 48 tissue types were downloaded and. The SMR method applies the Wald Ratio method systematically for all genes with an eQTL at P<1×10^−4^, using lead eQTL as instrumental variables and eczema estimates as the outcome. The HEIDI (heterogeneity in dependent instruments) test was applied to filter out genetically predicted effects which may be attributed to heterogeneity in a region which may lead to spurious results.

### Regfm and PrixFixe analysis

To further prioritize GWAS gene targets, we used two gene prioritisation methods: regfm[33] and PrixFixe[34]. PrixFixe strategy relies on prioritisation of groups of candidate genes from multiple GWAS loci based on ‘cofunction’ networks (CFNs). A genetic algorithm optimisation approach selects optimal sets consisting of one gene per locus where there exists a dense subnetwork of functional relationships with genes at other GWAS loci. Each gene is then scored to reflect its contribution to the top subnetworks. Regfm’s workflow involves intersection of fine-mapped credible interval SNPs with consensus DHS sites and genes whose expression they control predicted based on ROADMAP[63] chromatin accessibility and gene expression data to prioritise target genes.

### Variant functional prediction

KGGSeq[64] was used to measure non-coding variant regulatory potential and coding variant deleteriousness using functional scores derived by combining scores from CADD[65], DANN[66], Funseq2[67], fathmm-MKL[68], GWAS3D[69], SuRFR[70], GWAVA[71] algorithms. The new extended version of fathmm: fathmm-XF[72], GWAS4D[73] and fitCons[74] were also used independently. Overlap with ChIP-Seq defined binding sites of transcriptional regulators was cross-referenced in the ReMap2018 database[75]. Splicing regulatory potential of variants was evaluated with SPIDEX[76].

We also looked at variant overlap within different regulatory regions: insulator[77], promoter-enhancer interactions[78],[79],[80],[81],[82],[83],[84],[85],[86], regulatory non-coding RNAs[87],[88],[89],[90],[91], topologically associating domains (TADs)[92],[93],[79],[94],[81],[95], and CTCF binding sites[96] culled from various publications, using giggle[97] search engine. Independently, we looked for overlap inside Roadmap regions classified as containing active chromatin state (states 1-8)[63] and FAIRE-Seq-determined regions of accessible chromatin in human epidermis during barrier maturation and disruption[98]. Cell-type specific regulatory elements[99] were also annotated based on histone marks and chromatin state.

### Independent lookups

We have also performed gene and variant lookups among published significant results from various eQTL[21,51,99–125], mQTL (including GoDMC results[126])[121,124] pQTL[127,128],, hQTL[21,129], caQTL[130], where full GWAS results were not available, as well as differential expression[131–135], DNA methylation[136,137] and proteome[138,139] comparisons between skin in AD patients and healthy controls (Dataset S1). We also interrogated the GWAS Catalog[140] (accessed on 11/01/2019) for any variants that have been identified as genome-wide significant in previous GWAS studies on related inflammatory conditions. We used the significance threshold defined in each paper, as it varied due to a number of comparisons made.

### Generation of candidate gene and SNP rankings

The results of analyses and lookups listed above were then integrated to provide two rankings of: 1) all the SNPs within each GWAS locus interval and 2) all the genes within 3 Mbp window centred around index SNP. This was achieved by given a score to each piece of evidence and summing across these sources to generate a causal prioritisation score for every SNP and every gene tested. These scores represent the strength of evidence for a causal role of the SNP or gene in AD based on the evidence assimilated.

Detailed method of calculation of basic score per gene or variant in a given experiment/analysis is presented in Additional File 1: Supplementary Text. Briefly, the score is proportional to the magnitude of result effect or significance, down-weighted based on total SNP/gene hits in a locus in the experiment/analysis, *n* experiments in the study, and up-weighted based on pre-assigned evidence weight of the study (Dataset S2). Finally, after summing up scores from individual analyses/experiments, the score is further adjusted based on average number of studies and study types showing evidence for a given SNP/gene.

We down-weighted evidence from any single experiment/analysis which highlighted multiple genes/variants as being less specific. Down-weighting based on *n* experiments in the study was introduced since experiments in the same study are not independent, and so we wanted to attenuate the importance of multiple hits to experiments within the same study.

We also scaled the score by the weight of evidence, ie. subjective prior belief in strength of the evidence. We strongly prioritised hits with evidence weight 1 (formal test of association using the full set of summary statistics from experimental data - here finemapping, coloc, TWAS). The other two evidence weight categories included direct lookups among significant results obtained in experiments and predictions using a machine learning model trained on experimental data, for instance variant pathogenicity prediction.

Lastly, the total score for a given gene or SNP was adjusted by the average number of studies and study types showing evidence for a given SNP/gene, which aimed to promote heterogeneity of evidence sources and their types, such as different types of molecular markers. The decisions made in calculating the final score reflected various trade-offs. For example, for gene rankings, we wanted to reward consistent association of a given gene with SNPs in LD with lead variant, but at the same time did not want the score to be artificially inflated by repeated associations of different variants to the same gene in just one study.

## Results

### Identification of key tissues and cell types in AD GWAS loci

Skin, as the primary affected organ, and blood cell types, given the established immune component, are of relevance for AD[141]. We also attempted to identify additional tissues of importance by using gene-set enrichment methods on gene expression datasets across available tissue/cell types to identify any tissue/cell-specificity enriched in genes linked to our GWAS loci.

MAGMA gene-property analysis on 53 tissue types from GTEx ver. 7 identified significant enrichment (at *p* <0.001) for genes with tissue-specific expression in EBV-transformed lymphocytes, whole blood, spleen, sun-exposed and non-exposed skin at our GWAS loci. Despite not reaching statistical significance, we also included transformed fibroblasts (*p* = 0.1), due to the role of dermal fibroblasts in skin maintenance and repair[142].

SNPSea enrichment analysis in the Gene Atlas dataset prioritised (at permuted *p* <0.05, 79 tissues tested) whole blood and blood cell types: CD4+ T cells, CD14+ monocytes, CD8+ T cells, and dendritic cells. In addition, Immunological Genomics dataset (249 cell types) prioritised Natural Killer T cells, while FANTOM CAGE dataset (533 cell types) prioritised stimulated monocytes, basophils, Mast cells, eosinophils, neutrophils, Langerhans cells, CD34+ progenitor cells, hair follicle outer root sheath cells as well as spleen. For that reason, we decided to include datasets involving all possible types of immune cell datasets in our analysis, in addition to skin (including fibroblast), spleen and whole blood.

We reviewed the literature to identify 103 separate datasets from these tissue-types with relevant data for the prioritisation of genes (and SNPs). We identified 83 SNP-based datasets with individual variant-based (molecular QTLs, variant functional prediction software) or interval-based (TAD, promoter-enhancer interactions) information that we used to aid variant prioritisation. We identified 73 gene-based datasets with SNP-gene associations (molecular QTLs, promoter-enhancer interactions) or gene-centric (differential expression, GWAS target prioritisation pipelines – PrixFixe, regfm) information (Figure 1 subpanel 2, Additional file 2: Supplementary Figure 1).

**Figure 1.**
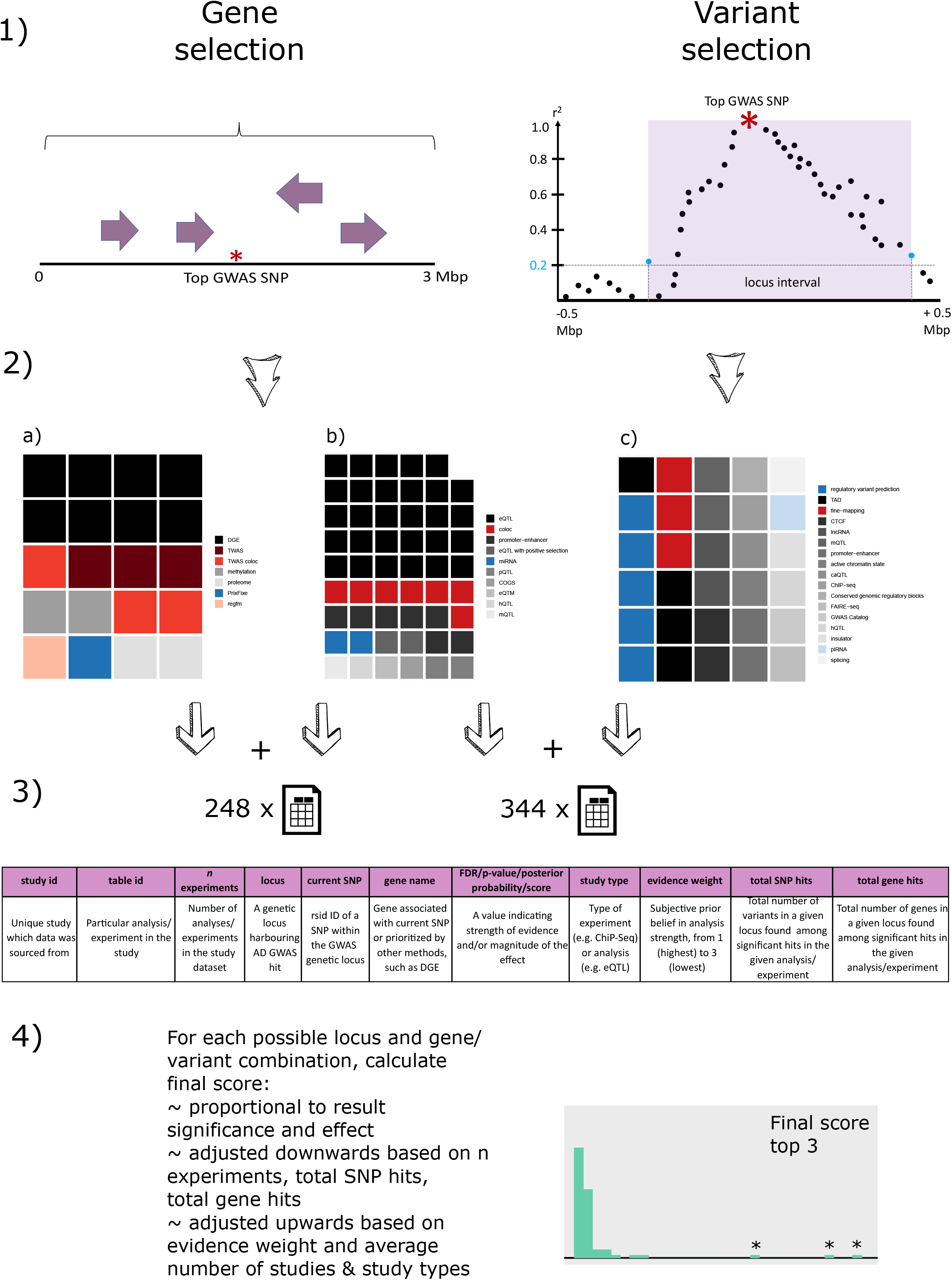
Outline of ADGAPP workflow used to prioritize candidate genes and variants based on AD GWAS. 1) *Left hand-side:* We considered all the genes positioned within a 3Mbp interval centred on index GWAS SNP. *Right hand-side:* We considered variants within the interval around top GWAS SNP defined as follows: consider all the SNPs within 1 Mbp interval around the index SNP and find the furthest SNP (blue circle) in either direction with r^2^ >= 0.2 in 1000 Genomes EUR population – these variants define the boundaries of the locus interval (purple shading) within which all SNPs are considered. For locus interval length, we found it ranged from 28,133 bp to 915,373 bp, with median at 228,670 bp. The number of candidate SNPs contained within a locus interval varied from 93 to 10,710, with a median of 758 SNPs. 2) We assembled different types of datasets showing significant results for genes within the loci (*left panel*), both genes and SNPs (*middle panel*) and just SNPs (*right panel*). Tiles represent number of datasets in each category and are further coloured according to subjective evidence strength: red (highest): statistical tests based on full summary statistics, gray (middle): lookups among significant results in experimental studies, blue (low): predictive machine learning models. 3) We summarised the output of for each experiment/analysis in a set of standardised summary tables. 4) We calculated a final score which allowed ranking of all the considered genes and SNPs for a given locus, and prioritisation of targets for downstream research.

### Prioritisation of candidate genes

ADGAPP generates a causal gene prioritisation score for every gene within a 3Mb window centred on each of the 25 index GWAS SNPs and a causal SNP prioritisation score for every SNP within the LD-generated boundary, which allows subsequent ranking of genes and variants at each locus (Figure 1 subpanel 1). To generate the prioritisation score, each source of evidence is assigned a weight based on subjective strength of evidence: highest for results from statistical tests using full set of summary statistics, such as molecular QTL colocalisation methods; lowest for prediction results from machine learning models such as variant functional prediction software and intermediate for positional overlap with significant experimental results, such as identified promoter-enhancer loops (Figure 1 subpanel 3 & 4). In calculating the final score, we also took into account the magnitude of result significance or effect, specificity (overall number of SNPs/genes significant in a given experiment), independence of evidence (number of experiments conducted in the same study, such as measuring both expression and DNA methylation levels). The final score was adjusted by heterogeneity of evidence (I.e. genes or variants consistently supported by a range of evidence sources - alternative functional assays and statistical methods – are upweighted in proportion to the square root of mean number of unique study types and unique study IDs), as well as absolute number of studies providing supportive evidence, consistent with criteria used in triangulation and assessment of causal links in epidemiology, such as Bradford-Hill criteria[143].

Gene prioritisation scores ranged from 0 to 1405 while for SNPs from 0.5 to 968 (Dataset S4). For 8 loci the top prioritised SNP was not the index SNP, and for 10 loci the closest gene did not score best (Table 1). In detailing the results, we focus on genes ranked in the top 3 and SNPs ranked in top 10 at each locus as although quite arbitrary, this limit agrees with the sharp score decay observed in ADGAPP scores (Additional File 3 & 4).

**Table 1.**
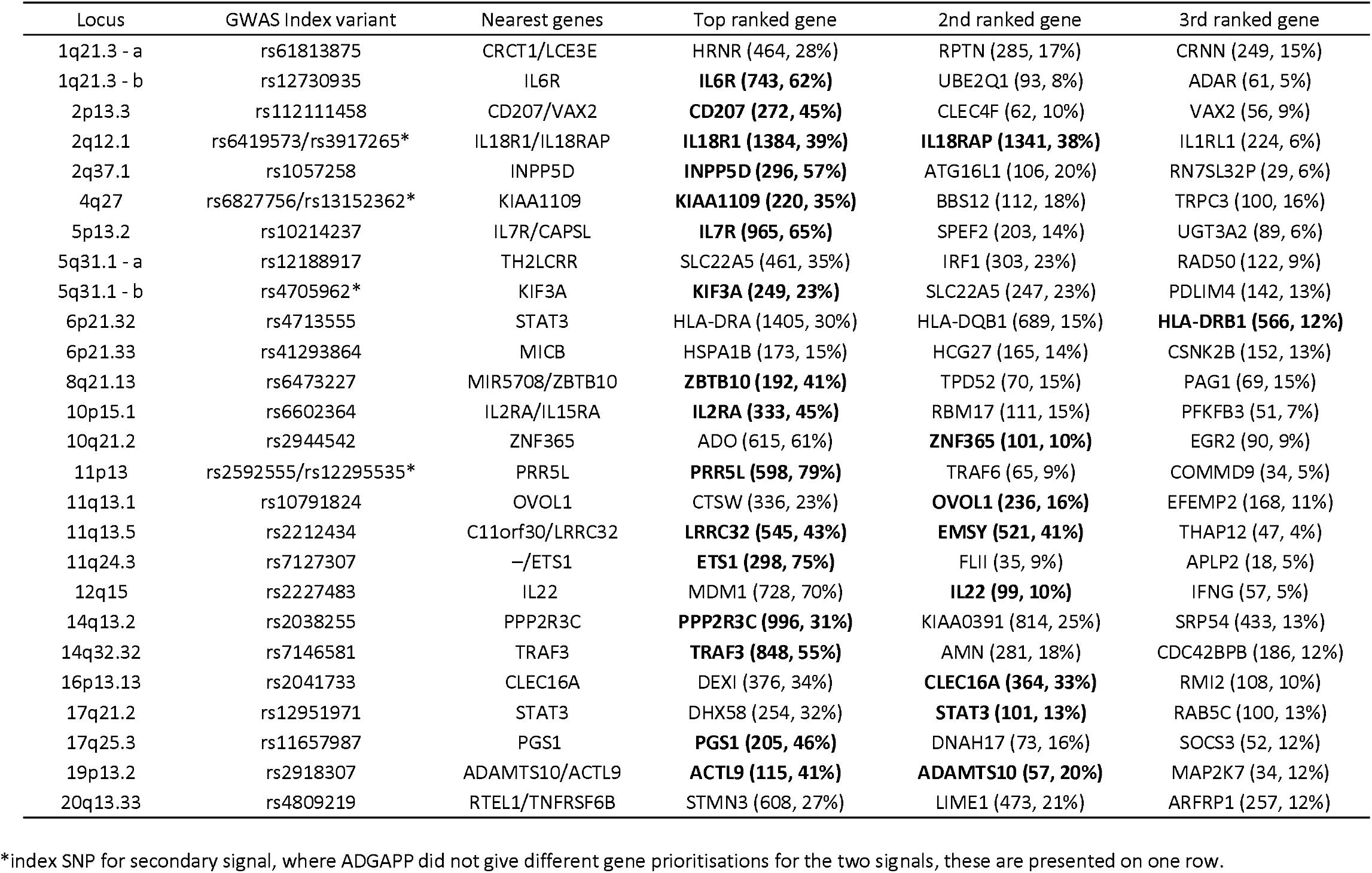
Genes prioritised by ADGAPP at atopic dermatitis GWAS loci. The closest genes to the index variant (in either direction) are marked in **bold**. The two values given in parentheses in top 3 ranked gene columns correspond to the ADGAPP gene prioritisation score and percentage of the total score for locus top 10 genes, respectively.

Excluding the complex MHC locus, the highest gene scores were seen for genes at 5 loci: *IL18R1* (score=1384) and *IL18RAP* (score= 1341) at the 2q12.1 locus, *PPP2R3C* (score= 996) at the 14q13.2 locus, *IL7R* (score= 965) at the 5p13.2 locus, *TRAF3* (score= 848) at 14q32.32 locus and *IL6R* (score= 743) at 1q21.3 locus (Table 1). Assuming that the true model is one of a single causal gene at each locus (unlikely to always be true), prioritisation can also be evaluated by comparing the score of the top prioritised gene at a locus with all other genes at that locus. Eight loci (1q21.3 - *IL6R*, 10q21.2 - *ADO*, 11p13 - *PRR5L*, 5p13.2 - *IL7R*, 11q24.3 - *ETS1*, 2q37.1 - *INPP5D*, 12q15 - *MDM1*, 14q32.32 - *TRAF3*; Table 1) have a single stand-out candidate for causal gene – with the top gene contributing more than 50% of the total score of top 10-ranked genes. The top candidate by that metric is *PRR5L* (79% of top 10 genes at 11p13 locus), with a score of 598 compared to 65 for the second-ranked gene at this locus. Most top-prioritised genes by the total score are also prioritised by this metric.

Two further loci show good evidence (>75% cumulative score) shared across two candidate genes (*IL18R1* and *IL18RAP at 2q12*.*1 and EMSY* and *LRRC32* at 11q13.5, which share 77% and 84% of the cumulative score respectively). At 2q12.1 (where *IL18R1* and *IL18RAP* reside) there is evidence for two independent genetic signals, and these may affect each of these genes.

Out of genes at these 11 loci with good prioritisation evidence, 6 have strong supportive eQTL colocalisation evidence (coloc posterior probability of colocalisation (PPH4) >90% or TWAS *p*-value <1 x 10^−5^, Figure 2, and Additional File 5: Supplementary Table 1): *PRR5L* in whole blood, LCL, skin and CD4+ T cells, *TRAF3* in whole blood, *IL6R* in whole blood, *IL18R1* in whole blood and LCL, *IL18RAP* in whole blood and CD4+ cells, *LRRC32* in whole blood, *PPP2R3C* in skin, CD15+ granulocytes, and whole blood. In addition, 20 other genes ranked 1-3 in ADGAPP have strong colocalisation evidence.

**Figure 2.**
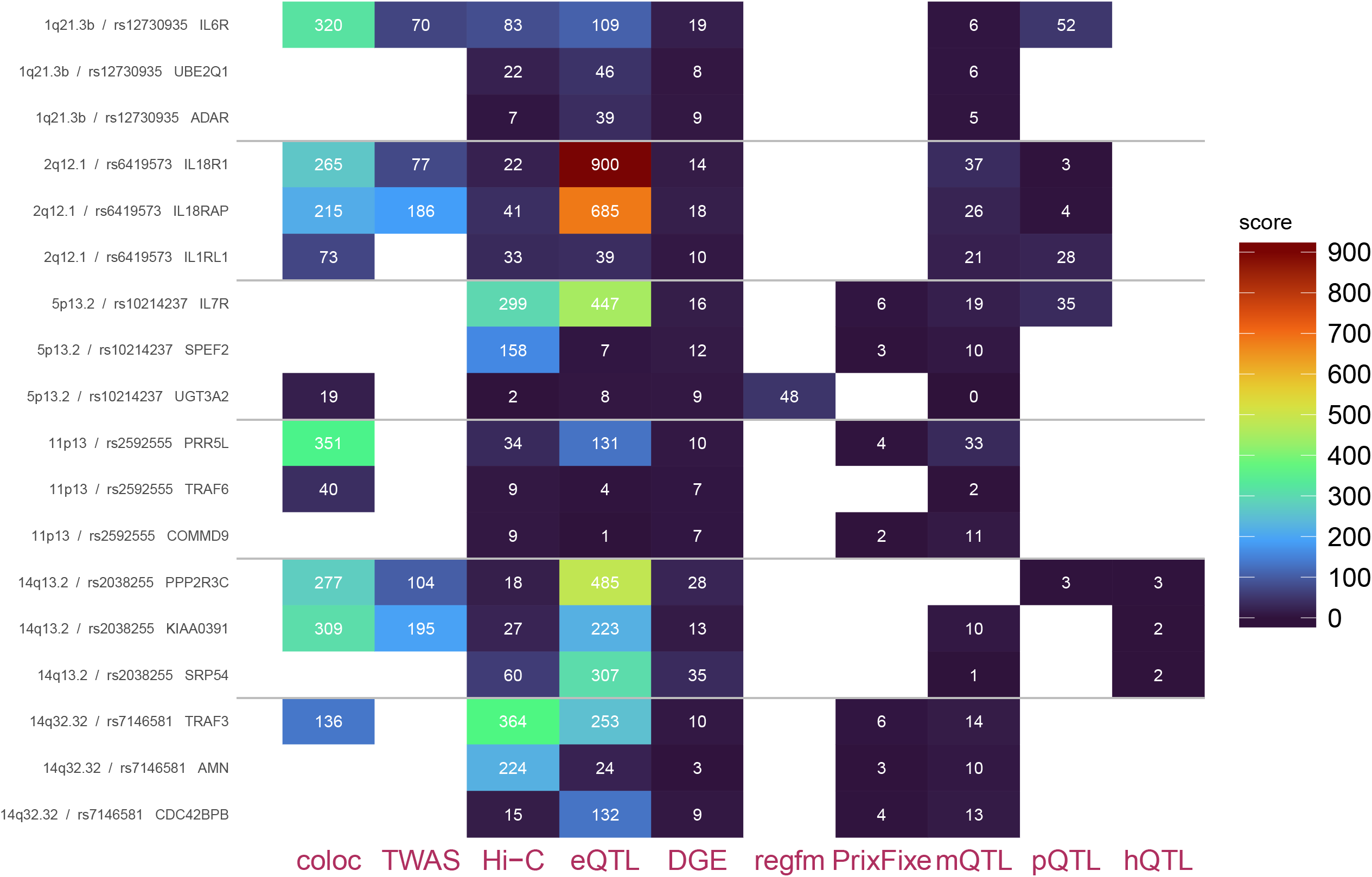
Score by type of evidence for top 3 ranked genes in the 6 highlighted loci. Scores for top 3 ranked genes at each locus are shown partitioned by category of evidence – here including the top 10 categories contributing the highest proportion of total score at the top 10 ranked genes for all loci. Order of loci corresponds to the order in Table 1.

For the majority of the loci, we prioritise genes that have previously been considered in annotation of these GWAS loci[11], but ADGAPP provides additional resolution on the likely causal genes through the integration of new evidence. Thus, for many loci we are able to strengthen the evidence for existing candidates. However, for some loci, our present analysis prioritises an alternative gene to the original GWAS. At 10p15.1 *IL15RA* was previously reported as the most likely candidate. Our ADGAPP analysis ranks *IL2RA* in the top spot (score=331). Subsequent to the EAGLE GWAS, CRISPR experiments have been conducted which have established that the SNP which ADGAPP ranks as top SNP at this locus, regulates *IL2RA* expression, providing validation for our prioritisation approach[144].

In addition, for five loci, ADGAPP prioritises genes in top position (and with a score >300) that were not considered in the original annotation of these loci in the GWAS paper[11]; *MDM1* at 12q15 (score=728), *ADO* at 10q21.2 (score=615), *STMN3* at 20q13.33 (score=608), *SLC22A5* at 5q31.1 (score=461) and *DEXI* at 16p13.13 (score=376). Some in this list (such as *SLC22A5*) represent promising looking candidates, whilst others are in loci where there are far more promising biological candidates (e.g. at 12q15, where *IL22* and *IFNG* appear to represent more plausible candidates than the top ranking *MDM1*). It will therefore be of interest to prospectively follow these loci, as more evidence is gathered and experimental work done, to assess whether any of the novel candidates presented here do in fact transpire to be the causal target genes.

In order to independently establish if restriction of the tissues to those known to be mechanistically linked to eczema and enriched in our GWAS signal in MAGMA and SNPSea analysis was likely to have influenced the final gene ranking, we compared our results to those run on the full set of 53 GTEx tissues, Blueprint and eQTLGen datasets using Mendelian randomization-based colocalisation method, SMR. Twelve out of 20 (Additional File 6: Supplementary Table 2) genes showing strong evidence (p<6.5 x 10^−6^) for colocalisation using SMR were among the top 3 hits for a given locus from our ADGAPP analysis. Of the remaining 8, 3 were prioritised by SMR in tissues absent in ADGAPP: the established eczema filaggrin gene in aorta artery (ranked 6 at 1q21.3 - a locus in ADGAPP), its antisense transcript *FLG-AS1* in esophagus muscularis (ranked 21 at the same locus), and *RTEL1* helicase in tibial and aorta artery (ranked 15 at the 20q13.33 locus). However, further investigation of these eQTL signals suggest some may not be biologically meaningful due to very low expression levels of these genes in the respective tissues (FLG TPM 0.59 in aorta artery and FLG-AS1 TPM 0.62 in esophagus muscularis in GTEx[20]) and so inclusion of likely irrelevant tissues may increase the chance of spurious results. Therefore, restriction to six GTEx tissues is unlikely to have had significant impact on the final gene ranking. On balance, restriction of tissues will retain eQTLs shared across tissues[14] but removes the problem of increased false positive rate for eQTLs in tissues less related to the pathophenotype[145].

### Prioritisation results for choice loci

A full discussion of each locus in Table 1 is available in Additional File 7: Supplementary Text 2. Here we highlight several loci with especially compelling prioritisation evidence (Figure 2 & 3, see Dataset S3 and Additional File 8: Supplementary Figure 4 for all loci) and integrate this with knowledge from literature.

**Figure 3.**
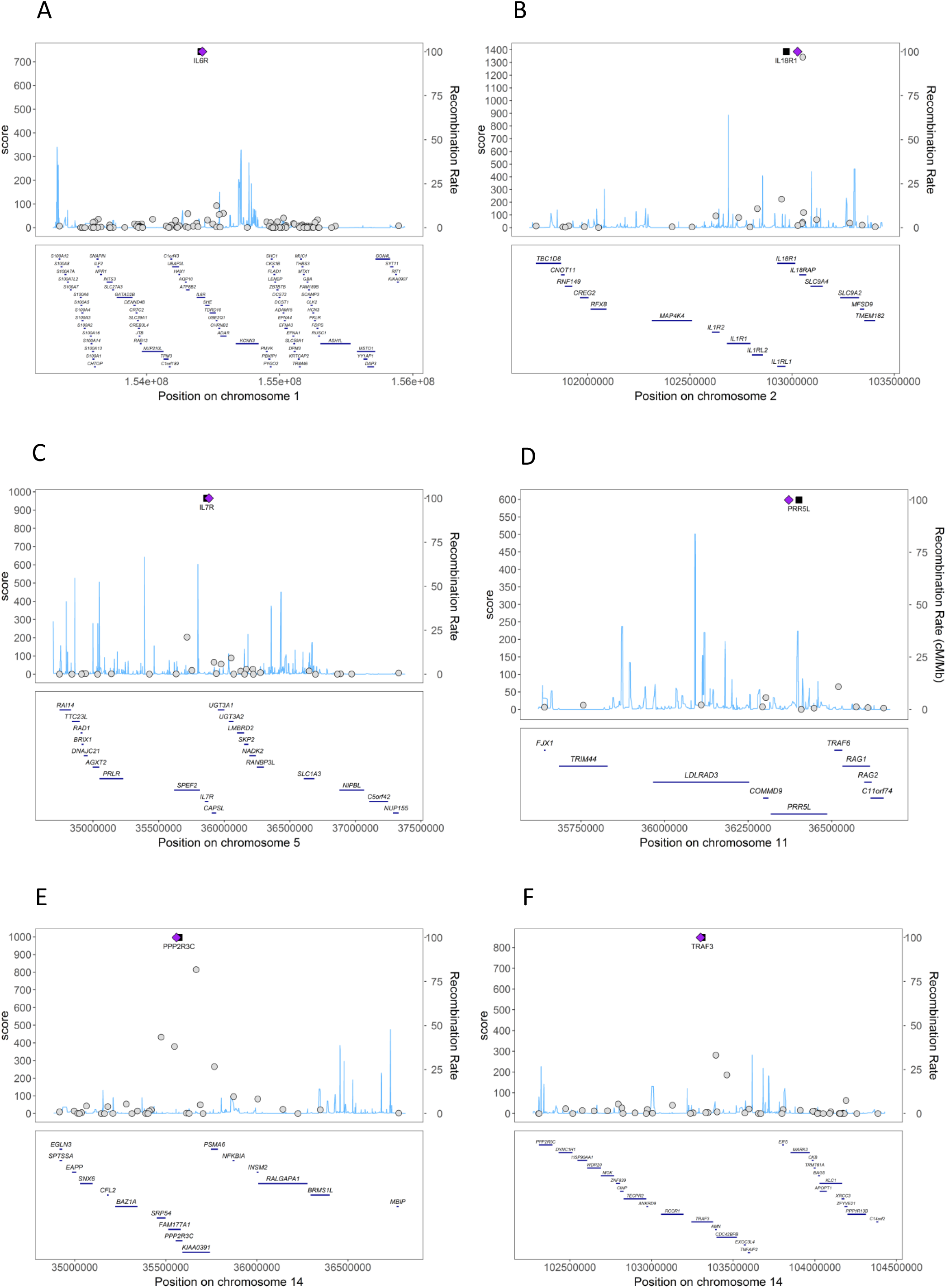
Gene scores within the 3 Mbp interval of lead SNP in the 6 highlighted loci. Top prioritised gene marked with a black square. A) locus 1q21.3 – b; B) locus 2q12.1; C) locus 5p13.2; D) locus 11p13; E) locus 14q13.2; F) locus 14q32.32.

### Locus 1q21.3 - b

At this locus we tested 104 genes within the 3 Mb interval (Figure 3A & Dataset S4). Interleukin 6 receptor (*IL6R)* had the highest prioritisation score – 743 (62% of top 10 cumulative score; Table 1 & Figure 3A), with the next best gene being *UBE2Q1*-score of 93 (only additional 8% of top 10 cumulative score). The high prioritisation of *IL6R* by ADGAPP is mostly driven by colocalisation (Figure 2) in whole blood in eQTLGen and GTex (coloc posterior probability of colocalisation (PPH4) = 63%-88% and TWAS *p*-value = 5 x 10^−6^), in lipopolysaccharide-/muramyl dipeptide-primed monocytes in Kim-Hellmuth *et al*. (PPH4 = 75%-77%) and sun-unexposed skin in GTEx (PPH4 = 64%) (Additional File 5: Supplementary Table 1[11]. Possible placement of many locus interval SNPs within an enhancer of the gene is highlighted by HiC interaction data in human embryonic stem cells[79], whole blood[81], CD34+ hematopoietic cells and lymphoblastoid cell lines[146] and epidermal stem cells along with keratinocytes[84]. Previously, *IL6R* was also implicated as the potential causal gene at this locus in the EAGLE GWAS[11], as one of the variants in the MANTRA-based credible set of 23 variants, rs2228145, represents a missense (Asp358Ala) mutation in the gene [147]. This variant is ranked 3^rd^ in ADGAPP and is associated with blood serum levels of IL6R protein as well as transcript[127]. However, the actual action of rs2228145 is to shift IL6R protein from membrane-bound state to soluble cell fraction, or in other words, from the anti-inflammatory classical signalling pathway to pro-inflammatory trans-signalling [148]. Mendelian randomization analyses indicated increased soluble IL6R levels as causal for higher AD (and asthma) risk and decreased soluble IL6R levels indicative of increased rheumatoid arthritis (RA) risk[149]. This is in agreement with the Asp358Ala mutation being protective towards RA and increasing risk in AD[150]. However, in our colocalisation analyses the AD risk allele is linked to overall reduction in *IL6R* transcript levels, likely due to the complex balance of IL6 classical and trans-signalling. A range of anti-IL6 biologics are in clinical use and development for inflammatory diseases, including, tocilizumab, which inhibits both soluble and membrane-bound IL6R, and is approved for treatment of RA and juvenile idiopathic arthritis[151]. However, AD adverse events have been reported in tocilizumab trials[152], consistent with the opposing effects of this GWAS locus on RA and AD.

### Locus 2q12.1

This locus consists of two independent signals represented by rs6419573 and rs3917265 lead SNPs[11]. Altogether, we considered 46 genes located within the 3 Mbp interval of each SNP (Figure 3B & Dataset S4). As mentioned earlier, the highest scores were obtained by *Interleukin-18 receptor 1* - *IL18R1* (score=1384, Table 1) and *Interleukin-18 receptor accessory protein 1* - *IL18RAP* (score=1341) which together account for 77% of the top 10 cumulative score at the locus, and so are likely to represent the causal genes behind the two signals. *IL18R1* and *IL18RAP*’s roles are supported by abundant expression colocalisation evidence in the whole blood in GTEx (*IL18R1*: PPH4 = 51%-80% and p-value = 7 x 10^−6^; *IL18RAP*: PPH4 = 96%-99% and p-value = 4 x 10^−12^) and immune cell types in CEDAR, TwinsUK and Kim-Hellmuth *et al*. (*IL18R1*: PPH4 = 55%-86% and p-value = 2 x 10^−5^, *IL18RAP*: PPH4 = 55%-97% and p-values = 5 x 10^−5^ to 6 x 10^−8^), as well as skin in TwinsUK for *IL18R1* only (p-value = 1 x 10^−4^; Figure 2 & Additional File 5: Supplementary Table 1). The direction of effect indicates increased expression of both genes for AD risk alleles, with one exception in GTEx whole blood for *IL18RAP* (Additional File 5: Supplementary Table 1). Interestingly, opposing expression effects of the same allele in a variant has previously been reported in monocytes and whole blood for *IL18RAP*[104]. SNPs in LD with the lead SNP at the three top-ranked genes overlapped whole blood mQTL SNPs in the GoDMC study[126], likely influencing the methylation of promoters of those genes. Differential gene expression (DGE) consistently shows significant upregulation of *IL18RAP*[132] and *IL18R1*[135,153] in the skin of AD patients (Dataset S5). Both genes were put forward as candidate genes in the EAGLE GWAS annotation, as the credible set variants span both genes and individual variants linked to *IL18R1* eQTLs in TwinsUK skin microarray data, but previously there was little evidence for colocalisation.

In general, cytokine IL-18, in the formation of whose receptor complex both IL18RAP and IL18R1 participate, has been shown to elicit significant immune gene expression change in keratinocytes from AD lesions[153]. *IL18R1* and *IL18RAP* are both involved in T-cell, especially T helper cell signalling and activation of the NFkB-pathway. IL18R1 and IL18RAP mediate IL-18-dependent signal transduction, which is of special importance in Th1 response. Thus, it would not be surprising if one variant affected the expression of the whole gene cluster, and that has indeed been shown to be the case with SNP rs917997 associated with IBD and coeliac disease[154]; and ranked 8^th^ among the two signals. A variant situated 15 kbp away from rs917997 - rs990171, again associated with celiac disease and lymphocyte counts[140], is ranked as second-best among the two signals in the locus (Dataset S4). There is strong pQTL support for association of rs990171 with IL18R and IL1RL1 resulting in increased protein availability [128] (Dataset S5). In addition, rs990171 overlaps allele-specific eQTLs for IL18RAP and acetylation hQTLs in neutrophils[21] appearing during Th1 polarization[155].

### Locus 5p13.2

We considered 41 potential candidate genes situated within the 3 Mbp window centred on index SNP for this locus (Figure 3C & Dataset S4). Most of the cumulative score was assigned to *interleukin-7 receptor subunit alpha* -*IL7R* (score=965, which contributes to 65% of top 10 cumulative score, Table 1), followed by *SPEF2* (score=203 and a further 14%). While our GWAS results do not directly colocalise with any eQTLs for the gene (in datasets where this could be tested), there are multiple eQTL associations for SNPs in LD with the index SNP: in whole blood[117,121], CD4+ T cells[21,102,103], macrophages[108] and monocytes[101] and pQTLs in whole blood[127,128] as well as promoter-enhancer interactions in human embryonic stem cells [79], CD34+ hematopoietic cells[146], naïve T regulatory cells and T helper 17 cells[156] (Figure 2, Dataset S5). *IL7R* is among the genes found to be upregulated in the skin in eczema patients in a meta-analysis[132]. Our results confirm initial prioritisation of *IL7R* in the GWAS which was supported by 18 credible set variants spanning the gene, including one non-synonymous (Thr>Ile) variant, rs6897932[11]. In ADGAPP, rs6897932 was ranked as the 8^th^ most likely causal variant. The variant affects splicing of the *IL7R* transcript, with the minor C allele, the risk allele for multiple sclerosis (MS), favouring secreted over surface isoform of the protein. Elevated levels of secreted isoform exacerbate symptoms of MS in animal model[157,158]. In common with opposite effects seen in AD compared to autoimmune diseases, the minor allele is protective in eczema[150]. *IL7R* is part of the thymic stromal lymphopoietin (TSLP) receptor/IL-7/IL7R axis required for correct lymphocyte maturation, especially of Th2 lymphocytes of interest in AD, with overexpression associated with acute lymphoblastic leukaemia[159], whereas recessive mutations in the gene resulting in reduction of gene expression is seen in severe combined immunodeficiency (SCID) patients[160].

### Locus 11p13

This locus stands out as having one clearly prioritised target gene - *PRR5L* (score=598, 79% of top 10 cumulative score; Table 1), with its final score nine times higher than the second-ranked gene, *TRAF6* (score=65 - only 9%). In total, we considered 15 genes located in the 3 Mbp region around index SNP (Figure 3D & Dataset S4). We could not tease apart the target behind the secondary signal represented by lead SNP rs12295535, with the same genes prioritised as for the primary signal represented by rs2592555. rs2592555, the top prioritised variant at the locus (score=246, Dataset S4), is situated in the intron of *proline rich 5 like (PRR5L)*. We also note that ADGAPP’s second best SNP rs7925585 (score=132) for the primary signal, also positioned within the intron of *PRR5L*, was independently prioritised for eczema in FINDOR analysis[161]. Strong expression colocalisation was detected with the coloc method (Figure 2, Additional File 5: Supplementary Table 1) in: eQTLGen - whole blood (PPH4 = 95%); TwinsUK – skin (PPH4 = 91%), LCL (PPH4 = 98%); CEDAR - CD4+ T cells (PPH4 = 98%) with protective AD allele associated with increased expression. The gene’s role was also supported using the network method PrixFixe (Dataset S5) and strongest methylation signal detected through mQTL overlap with locus interval SNPs in whole blood in the GoDMC study[126]. *PRR5L* was proposed as the candidate gene in the EAGLE GWAS due to position of the lead SNP in *PRR5L’s* intron and *PRR5L* eQTL overlap with the credible set variants[11]. However, there was previously very little evidence for colocalisation of these signals.

PRR5L is part of the rapamycin complex 2 (mTORC2) which responds to extrinsic stimuli through cytoskeleton re-organisation and cell migration[162]. PRR5L specifically plays a role in regulation of fibroblast migration, and decreased expression of the gene conferred by the risk allele is predicted to lead to increase in fibroblast migration.

### Locus 14q13.2

Out of 70 genes considered as causal at this locus (Figure 3E & Dataset S4), we find 2 of them to have comparably high scores - *Protein phosphatase 2 regulatory subunit B’’Gamma* (*PPP2R3C*) with the score of 996 (31% of top 10 cumulative score, Table 1), and *KIAA0391* with the score of 814 (25% of top 10 cumulative score).

Three (PP2R3C, KIAA0391, *FAM177A1*) out of the four top-ranking ADGAPP genes at this locus display partial co-expression. The top-ranked candidate gene *PPP2R3C* shows colocalisation (Figure 2, Additional File 5: Supplementary Table 1) in sun-exposed (PPH4 = 94%) and unexposed skin (PPH4 = 95%, *p*-value = 2 x 10^−7^), whole blood in GTEx (PPH4 = 97%, *p*-value = 2 x 10^−7^), CD15+ granulocytes (PPH4 = 96%) and colon (PPH4 = 96%) in CEDAR, and neutrophils (PPH4 = 93%) in Blueprint. The next best gene, *KIAA0391* recapitulates the colocalisation in the skin in GTEx (sun exposed PPH4 = 97%, unexposed PPH4 = 96%), and LCL and individual immune cell types: LCL from TwinsUK (PPH4 = 94%, *p*-value = 3 x 10^−9^), CD8+ T cells (PPH4 = 97%) and CD14+ monocytes from CEDAR cohort (PPH4 = 60%, *p*-value = 1 x 10^−4^), in addition to the spleen in GTEx (PPH4 = 95% and *p*-value = 2 x 10^−7^). Moreover, we find hundreds of individual variants in LD with the index SNP to overlap blood and skin eQTLs for those genes (Dataset S5). The orchestrated expression of these genes is underscored by their differential expression in atopic dermatitis skin: *PPP2R3C* is upregulated regardless of *FLG* genotype[135,137] and *KIAA0391* is strongly downregulated in homozygous *FLG* mutation AD patients[135]. Comparing that with our colocalisation results, we see tissue-specific regulation for *PPP2R3C*, upregulation in the skin (in line with the differential expression results) but downregulation in the blood associated with the AD risk allele. For *KIAA0391*, the downregulated expression seen in AD patients is at odds with the eQTL result, where the AD risk allele is associated with increased expression; this conflict could indicate that *KIAA0391* is not an AD susceptibility gene. The original GWAS annotation[11] also suggested *PPP2R3C, KIAA0391* and *FAM177A1* as plausible causal genes, with the 47 credible interval SNPs scattered throughout the three genes and the lead SNP mapping to an intron within *PPP2R3C*; strong colocalisation with TwinsUK microarray eQTLs in that paper was found only for *KIAA0391*.

Considering gene function, *PPP2R3C* is the most viable candidate. Targeted B-cell mouse knockout mutants show profound abnormalities in humoral immune response: reduced B cell proliferation, maturation, abnormal activation and small spleen[163]. Similarly, loss of *PPP2R3C* in T cells results in atrophy of the thymus, decreased thymocyte abundance, especially of CD4+ and CD8+ double-positive thymocytes[164]. Variants at the locus related to pleiotropic phenotype of chronic inflammatory diseases (AS, CD, psoriasis, primary sclerosing cholangitis, UC)[165] map to intron positions within *PPP2R3C*. Next, linking the gene’s mutant phenotype established in mouse to human GWAS, it has been reported that decrease in lymphocyte counts[166] significantly correlates with the risk allele at rs2038255, the index variant in AD GWAS and ADGAPP top prioritised variant (score=389, Dataset S4). *PPP2R3C* encodes a regulatory subunit of protein phosphatase 2A, known as G5PR, which associates with phosphatases PP2A, PP5, GANP protein and represses JNK and IKK*β* (inhibitor of NF-*κ*B) phosphorylation[167]. It is involved in regulating antigen-based B-cell and early T cell selection in the thymus promoting thymocyte and B cell survival. G5PR becomes upregulated in activated B cells and prevents B-cell receptor-mediated activation-induced cell death in B cells through suppression of late-phase JNK activation[168]. Overexpression results in the increase of production of non-specific B cells after immunisation and generation of autoantibodies in non-stimulated mice[169]. Therefore, *PPP2R3C* upregulation in the skin could be a contributing factor to autoimmune activation seen in a subset of severe AD patients and particularly directed against epidermal proteins[170].

*KIAA0391* in contrast does not appear to be so directly functionally linked to the AD phenotype. It encodes a component of mitochondrial RNase P complex which catalyses the last step in pre-tRNA maturation process: removal of the tRNA 5’ leader sequence[171].

### Locus 14q32.32

Accounting for 55% of the cumulative score at the locus, *TNF receptor associated factor 3 (TRAF3)* is clearly prioritised among the 59 genes positioned within 3 Mbp of the index SNP at the locus (Figure 3F & Dataset S4). While TRAF3’s score is 848, the second-ranked AMN scores only 281 (18%) (Table 1). *TRAF3* is the only gene at the locus with direct colocalisation evidence (Additional File 5: Supplementary Table 1): in the whole blood in eQTLGen (PPH4 = 93%) and with lower confidence (PPH4 = 85%) in CD4+ T cells in Blueprint. In addition, many locus interval SNPs are possibly situated within an enhancer interacting the gene’s promoter in human embryonic stem cells[79], whole blood[81], CD34+ hematopoietic cells and lymphoblastoid cell lines[146], naïve T regulatory cells and T helper 17 cells[156] and epidermal stem cells along with keratinocytes[84] as shown by Hi-C data. In AD GWAS, risk alleles correlate with upregulated expression of TRAF3 and in IBD there is increased expression of the gene in inflamed intestinal mucosa[172]. However, changes in expression have not been consistently observed in AD lesions relative to healthy skin (Dataset S5).

The original EAGLE GWAS annotation also presents *TRAF3* as the candidate gene at the locus due to the location of the index SNP, pathway enrichment in MAGENTA gene set analysis and mouse knockout phenotype, but no eQTL evidence[11]. Two out of the 3 top prioritised SNPs at the locus in ADGAPP (1^st^ ranked rs79589176 and 3^rd^ ranked rs12880641) as well as index SNP (rs7146581) are intronic, situated within the *TRAF3* gene, whereas the remaining top 3 SNP (2^nd^ ranked rs71421262) is 2 kbp 5’ upstream of *TRAF3. TRAF3*’s role in signal transduction in immunity is well-established, with early studies describing a serious imbalance in T cell composition in mouse model knockouts which eventually leads to their perinatal death[173]. TRAF3 is a repressor of CD40- and B cell-activating factor-mediated signalling and limits homeostatic B cell survival[174]. *TRAF3* is also related to possible candidates at two other GWAS loci (*TRAF6* at 11p13 and *IL6R* at 1q21.3). TRAF6 positively regulates MAPK signalling and production of inflammatory cytokines and chemokines, whereas TRAF3 needs to be degradatively ubiqutinated during MyD88-dependent Toll-like receptor signalling to activate the JNK and p38 MAPK cascade[175]. TRAF3 exerts a negative effect on Th17-based inflammation by sequestering IL-17R, however, normally this is prevented by competitive binding of TRAF3 by NRD1. This allows formation of IL17R-Act1-TRAF6 complex and subsequent propagation of IL-17-induced signal down through MAPK and NF-κB pathways leading to production of pro-inflammatory molecules, including cytokine IL-6, whose receptor is prioritised in the AD GWAS at locus 1q21.3[176].

### Validation of ADGAPP gene prioritisation – enrichment and network analysis

Due to limited knowledge of heritable atopic dermatitis loci, we did not have any significant number of “gold standard” true positive genes to which we could compare our ranking of prioritised genes in the atopic dermatitis GWAS. For that reason, we evaluated the quality of our results in two indirect ways.

Firstly, many GWAS gene prioritisation algorithms focus on prioritising genes which share similar profiles, be it in membership in gene sets, co-expression or protein-protein interaction networks[177]. Here, we work this approach in reverse and ask if our prioritised genes are enriched for their presence in any gene sets and networks. Using enrichr[178], we carried out gene set enrichment tests of our top 3 prioritized genes, to see if they align well with categories from additional previously implicated AD genes (Additional File 9: Supplementary Table 3, Additional File 10: Supplementary Table 4). In general, highly-prioritised genes had functions related chiefly to the immune system but also dermis structure, lipid metabolism and cytoskeleton organisation.

We find that both lists are significantly enriched for immune system-related genes (Figure 4). In particular, cytokine categories were overrepresented: GO cytokine-mediated signalling pathway (adjusted *p*-value for ADGAPP prioritised genes = 1×10^−9^ versus 0.004 for other previously implicated AD genes), positive regulation of cytokine production (GO, 0.009 ver. 9 x 10^−4^), cellular response to cytokine stimulus (GO, p=0.011 ver. 0.009), cytokine-cytokine receptor interaction (KEGG, p=1 x 10^−4^ ver. 7 x 10^−4^), interleukin-7-mediated (GO, p=0.053 ver. 0.048) and interleukin-4-mediated signalling pathways (NCI, p=0.011 ver. 0.007), interleukin-2 (Jensen, p=0.003 ver. 0.035), interleukin-12 (Jensen, p=1 x 10^−5^ ver. 0.001) and interleukin-23 complex (Jensen, p=7 x 10^−6^ ver. 0.010). The genes in the cytokine pathways identified by ADGAPP include *IL6R, IL22, INPP5D, IL2RA, IFNG, IL18R1, IL18RAP, IL1RL1* and *IL7R*.

**Figure 4.**
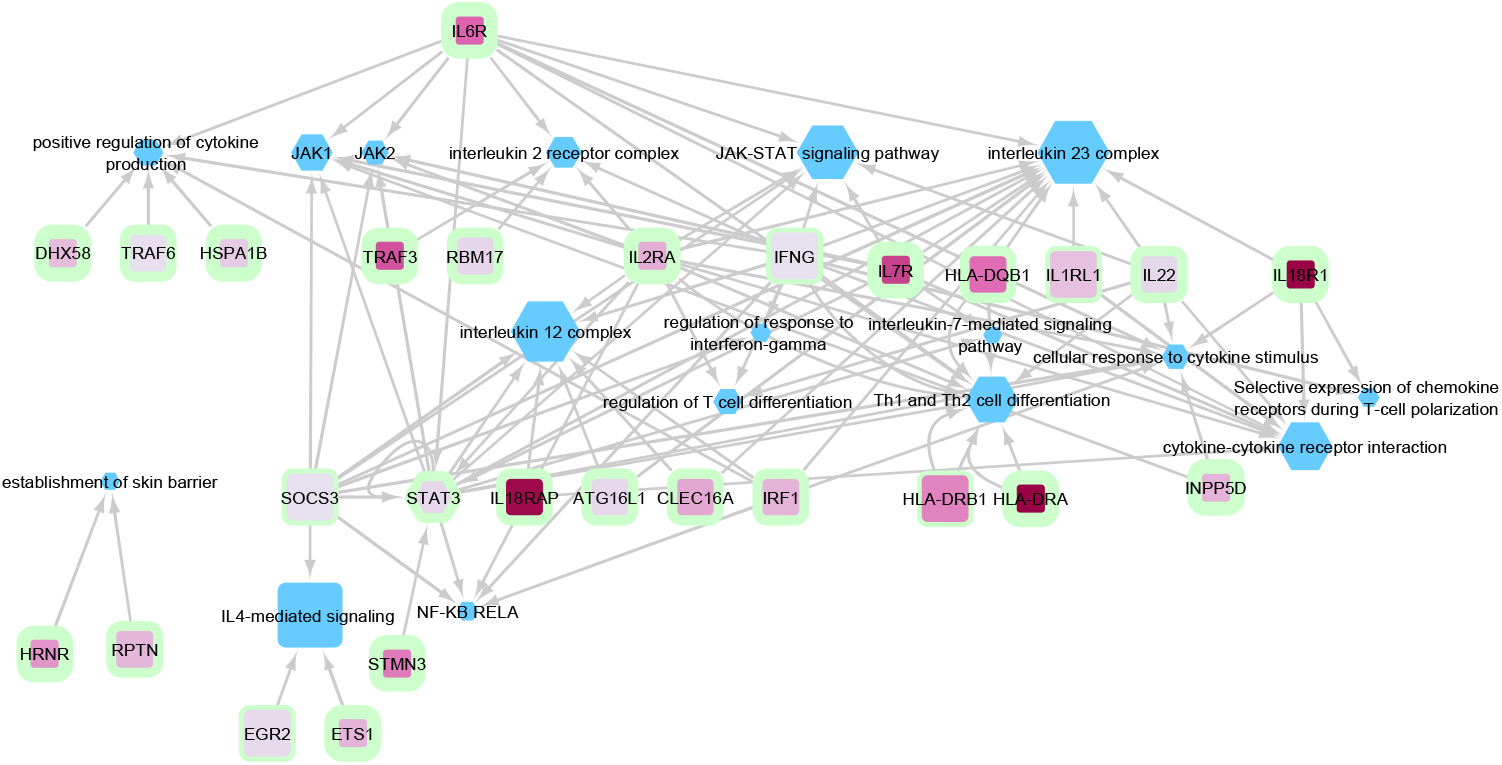
Network visualisation of the functional terms enriched among locus top 3 prioritised genes in ADGAPP. The ontology categories are depicted as blue hexagons, with their size linearly proportional to -log_10_ of adjusted enrichment *p*-value. AD genes are depicted as pink rectangles, with the intensity of the colour fill proportional to gene score and thickness of the green border marking the gene rank at the locus, with rank 1 the thickest.

Signalling involved in regulation of response to interferon γ (GO, p=0.039 ver. 0.043), JAK1-/JAK2-STAT3-interacting genes and JAK-STAT signalling pathway in general (KEGG, p=4 x 10^−5^ ver. 2 x 10^−4^),genes downstream of NF-κB-RelA transcription factor (TRRUST, p=0.036 ver. 0.012) also overlapped between the two gene sets, as did regulation of T cell differentiation (GO, p=0.011 ver. 0.007), selective expression of chemokine receptors during T-cell polarization (WikiPathways, p=0.036 ver. 0.004) and Th1, Th2 (KEGG, p=2 x 10^−4^ ver. 6 x 10^−4^), Th17 cell differentiation (KEGG, p=4 x 10^−7^ ver. 7 x 10^−4^). Beside the previously mentioned cytokine-related genes, we found other targets in different immune pathways: e.g. *STAT3, SOCS3, ETS1, TRAF3, TRAF6, IRF1*. We did not find enrichment of genes in any specific type of immunity – with all of Th1, Th2, Th17, Th22 represented and previously shown to play a role in certain subsets of AD patients, despite overall particular importance of Th2 and Th22[36,179,180]. We did not see much in the way of B cell-specific effects, despite selective expansion of certain B-cell subsets in AD patients[181].

Genes concerned with establishment of the skin barrier were marginally enriched for in ADGAPP (due to the prioritisation of cornified envelope genes, *HRNR* and *RPTN*), but less than the previously reported AD genes (GO, p=0.045 ver. 8 x 10^−8^).

The second way we validated our results was to test if our candidates interacted with each other and with the genes with established roles in AD pathogenesis. We used STRING[182] to visualise the highest-confidence interactions (Figure 5) among the top 3 prioritised genes at each locus from ADGAPP and other AD genes previously implicated (Additional File 9: Supplementary Table 3). Not surprisingly, the analysis revealed an extensive network that included 25 ADGAPP prioritised genes, centred on key immune regulators, such as *STAT3, STAT6, SOCS3, IRF1, TRAF6*. It included direct binding interactions between targets prioritised in the current AD GWAS and outside of it – between *INPP5D* and *FCER1G* as well as *FCER1A, IL7R*/*STAT3* and *TSLP, IL7R* and *TSLPR, IFNG* and *IFNGR1;* and *SOCS3* and *IFNGR1*. However, when it came to the genes directly taking part in establishing skin barrier, 2^nd^ ranked gene at the epidermal differentiation locus - *RPTN*, was the only one shown to interact with the late cornified envelope genes.

**Figure 5.**
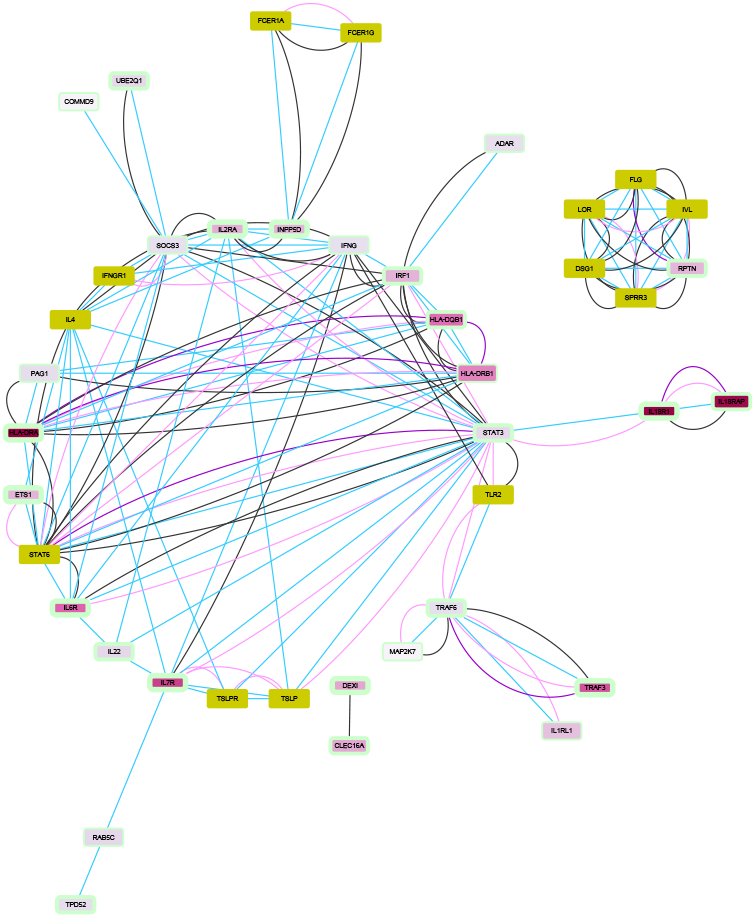
Highest-confidence interactions between locus top 3 prioritised genes in ADGAPP and other AD genes. AD genes prioritised outside of ADGAPP are depicted as olive rectangles. For GWAS AD gene legend, refer to Figure 4. Edges are coloured according to source of evidence for interaction: light blue - known interactions from curated databases, pink - experimentally determined known interactions, green - predicted interactions based on gene neighbourhood, red - predicted interactions based on gene fusions, dark blue - predicted interactions based on gene co-occurrence, black - co-expression, purple - protein homology

## Discussion

Previous annotations of AD GWAS loci have been limited in their ability to identify likely causal genes[11]. Here we provide a thorough and objective investigation of the 25 European AD loci, by integrating all relevant available data that can be used to provide evidence for identifying potentially causal genes, and combine this data in such a way as to produce a ranking for every gene at each locus. We intend that this thorough analysis and ranking will provide a prioritised list of AD candidate genes for further experimental work, and the detailed presentation of evidence for each gene will enable appropriate and tailored follow-up investigations to be designed.

For 10 loci the top ranked gene is not the gene closest to the index GWAS SNP. 8 loci have a single stand-out candidate causal gene (score >50% of the top 10 gene cumulative score) and 7 genes score particularly high (>700) and/or have a particular stand-out score (>75%). These are *IL6R* at 1q21.3, *IL18R1*/*IL18RAP* at 2q12.1, *PRR5L* at 11p13, *IL7R* at 5p13.2, *PP2R3C* at 14q13.2 and *TRAF3* at 14q32.32. Whilst in many cases our analysis strengthens the evidence for existing candidate causal genes at these loci, for 6 loci our score ranks alternative candidates as the most likely causal gene.

One of these 6 can be considered an interesting validation of our approach. *IL15RA* was previously considered the most plausible candidate gene at the 10p15.1 locus due to the limited eQTL evidence that was available at the time. Our ADGAPP approach however prioritised *IL2RA* over *IL15RA*. Since the publication of the GWAS in 2015 this locus has been followed up with CRISPR experiments, which reported that the T-allele at rs61839660 down-regulates IL2RA expression[144], suggesting that ADGAPP’s prioritisation at this locus is correct.

Other validations of our approach are provided by tests of enrichment of ontology terms and evidence of protein-protein interactions amongst the top ranked genes across all loci. Enrichment (amongst the top 3 prioritised genes at each locus) was found for the following ontology terms: skin barrier integrity, T helper cell polarisation, cytokine signalling and JAK-STAT signalling. JAK-STAT signalling has recently been confirmed as enriched for among genes prioritised for inflammatory skin diseases (including AD) with HiChIP-derived T cell enhancer connectome[183]. Furthermore, our enrichment analysis highlighted the role of JAK1, JAK2-interacting genes, and interestingly the two genes themselves were shown to contain an over-representation of rare coding variants in a new AD study[184], which underscores the functional importance of the pathway in AD.

Analogously, in investigating protein-protein interactions (using the STRING database) among our ADGAPP-prioritised candidate genes and other established candidates interactions between genes with immune regulation (but not skin barrier) functions were found amongst the established AD players: *TSLP* and its receptor, *TLR2, STAT6, IL4* and interferon γ receptor. Some roles in skin maintenance are supported by some of our candidate gene network modules. The *gp130-SOCS3-STAT3* axis promotes proliferation, migration and differentiation of keratinocytes[185] in skin-wound healing, activated amongst other by IL-6. SOCS3 negatively regulates STAT3 activation by simultaneously binding to phosphorylated JAK kinase and gp130 cytokine receptor[186].

STRING data is not entirely comprehensive and omits other functional relationships between prioritised genes. ETS1 activation results in downregulation of IL-6 pathway and in STAT3 phosphorylation[187]. SOCS3 antagonises IL1-dependent activation of NFkB pathway by inhibiting ubiquitination of TRAF6 in the TRAF-6/TAK1 complex[188]. LGP2’s negative regulation of type 1 interferon response is exerted through antagonizing TRAF ubiquitin ligase activity, including TRAF3 and TRAF6[189]. Finally, STAT3 is a regulator necessary for maintenance of STMN3 expression, and its phosphorylation as well as expression correlates with STMN3 expression[190]. STMN3 interacts with STAT3 via its C-terminal tubulin-associating region[191,192].

In general, the results of our GWAS prioritisation analysis remind us that interpretation of a GWAS locus is complicated due to varying regulation between cell types and widespread coregulation that makes identification of the true causal gene difficult. Indeed, recent GWAS explosion reveals that on top of each locus being able to contain multiple signals[193], each signal can influence multiple co-regulated genes[194].

Associations with molecular phenotypes follow the same pattern, with at least 9% of human eQTLs quantified to contain secondary signals[195] and multiple genes implicated for 50% human eQTLs[14]. It is well established that genes in the same locus show often correlated expression, especially for genes localised in the same TAD[196], and that correlation itself is heritable[197]. According to the multiple enhancer variant hypothesis, several variants in LD can influence multiple enhancers and cooperatively affect expression of target gene(s). Corradin *et al*. (2014) provide evidence for it in 6 autoimmune diseases, including RA, Crohn’s disease and SLE[198], while two adjacent thyroid-related genes are regulated by an enhancer haplotype at a thyroid cancer risk locus[199]. Therefore, it is not surprising that many of our loci showed multiple colocalisations for different genes and tissues, especially in gene-dense regions, with the caveat that not all may be causal. A recent analysis of the TWAS colocalisation method claims that around 75% of hits will be non-causal in the instance of correlated gene expression at the locus[145], and we hypothesize that may be the case at loci 11q13.1, 14q13.2, and 20q13.33, where expression of as many as 4-6 genes colocalises with AD GWAS signal in the TWAS results, alone. Still, due to a distinct possibility of detection of multiple target genes and variants at a locus, we do not focus only on top-rated hits in our gene and variant ranking. AD GWAS loci which we believe should be further experimentally investigated in that regard, include: 2q12.1 (*IL18R1, IL18RAP, IL1R1*), 5q31.1 (*KIF3A, PDLIM4, SLC22A4, IRF1*) and 20q13.33 (*STMN3, LIME1, ARFRP1*) – the first two especially due to containing at least two independent signals in the GWAS analysis.

Most of the genes which show eQTL colocalisation across many tissues indicate the same direction of effect, such as in the case of *PRR5L*, a top prioritised hit at the 11p13 locus, where the protective allele is associated with increased expression in the skin, whole blood and immune cell subsets. However, the results at three loci (2q12.1, 14q13.2, and 20q13.33) imply there may be tissue-dependent effects on expression. Protective alleles in the skin eQTLs showed positive effect and negative effect on expression in blood eQTLs in *STMN3* and *LIME1*, while reverse pattern was seen for *ARFRP1*, and similarly, change of sign between tissue types was also seen for *IL18RAP* and *PP2R3C*. This indicates that causal variants potentially reside in tissue type-specific regulatory regions and context-dependent effect of these genes could impact atopic dermatitis phenotype.

However, even when focusing on a single tissue of interest, we frequently observe lack of concordance in results among same tissues from different datasets, which was particularly clear in the eQTL colocalisation analysis, where we used eQTLs for the same tissues sourced from different studies: skin and LCL from both GTEx and TwinsUK, whole blood from GTEx and eQTLGen, CD4+ T cells from Blueprint and CEDAR. Hits such as *SLC22A5, PRR5L, IL2RA, TRAF3, FAM177A1* showed strong colocalisation in one dataset which was then missing in another. This could be explained by a host of factors relating to study design, experimental methods and data analysis choices. The first two include sample size and consequently statistical power, demographic composition of studied cohort, differences in harvested cell composition and condition, and technical details determining sensitivity of the assay. In the third category, choices regarding data clean-up such as quality control filters and data analysis parameters such as confounders and genome window size used for eQTL detection can also prevent SNP inclusion in QTL analyses and thus affect our ability to interrogate the data for a particular association. Nevertheless, consistent eQTL colocalisation replication across multiple datasets and tissues, whenever seen - as is the case e.g. for *IL6R* in whole blood instils us with more confidence that the effect seen is real.

Our analysis confirmed many loci where there is pleiotropy between atopy and inflammatory diseases. For three genes, the direction of effect matches: *DEXI* (T1D, MS), *SOCS3* (IBD), *ETS1* (SLE), but for many others the direction of effect is opposite. This is not surprising, as despite generally opposing types of immune activation in psoriasis versus eczema (Th17-skewed versus Th2-skewed), yet 81% of differentially expressed genes in AD are also DEGs in psoriasis[200]. We observe such contrasts for several immune regulators: *INPP5D* (IBD), *TRAF3* (IBD), *IL6R* (RA), *IL7R* (MS), *IL2RA* (T1D), *CLEC16A* (MS) which probably stems from different Th1/Th2/Th17 balance in adaptive immune system dysregulation in those diseases[201–204].

A strength of our variant prioritisation analysis is that we used three different Bayesian variant fine-mapping methods: JAM, Paintor and Finemap. We integrated the three methods (ran with different parameters) in light of previously shown incongruence in their results resulting from different model assumptions[205]. A notable caveat in our fine-mapping of variants is exclusion of indels and bigger structural variants. Therefore, variant prioritisation results do not aim to precisely identify the causal variants but finemap the broader region with the aim of prioritising SNPs which in turn may provide additional evidence for what genes in the region are likely to be involved. Our fine-mapping was also restricted by the power and imputation panel (1,000 Genomes) of the published EAGLE GWAS[11]. Future GWAS with bigger sample size and denser variant imputations will increase the power of future fine-mapping and, in turn, causal gene identification attempts.

Our ADGAPP pipeline for follow-up of GWAS signals, whilst focused on the integration of AD-relevant resources in this use case, can be easily adapted for other diseases or traits, following identification of the most relevant and powerful molecular datasets. For some traits (e.g. blood-related) there are very well-powered and informative accessible datasets (e.g. eQTLGen) while the more specific data may still be lacking, for instance different layers and cell types within the skin. Better predictions may be made in the future by integrating evidence from across more tissue types, especially at a single-cell resolution – such datasets are already being generated for related disorders, such as asthma[206]; considering *trans-* and isoform-level mechanism of action, and explicitly modelling network connectivity via protein-protein interactions and co-expression.

Whilst there are limitations in our approach, as outlined in the sections above, we nevertheless find ADGAPP useful to easily flag the genes where we find most evidence which can then be carefully evaluated. Furthermore, using our standardised scoring system to assess prioritisation, we can use absolute score to make direct comparison of strength of prioritisation across and within loci, % of cumulative score captured by top gene facilitates comparison of strength of evidence for genes within the locus. We use a combination of the two useful metrics to propose the most plausible candidate genes in the AD GWAS. Loci where we are more confident in prioritisation of single genes, lend themselves to direct experimental investigation, such as *TRAF3* at the 14q32.32 locus and *PRR5L* at the 11p13 locus. Additionally, investigating loci with clear candidate genes and association with multiple inflammatory diseases showing consistent direction of effect, such as 11p13 (*PRR5L* – MS, asthma), 11q24 (*ETS1* - psoriasis, celiac disease) and 16p13.13 (*DEXI* and *CLEC16A -* T1D, MS, alopecia areata, SLE, asthma) may reveal promising targets with potential drug repurposing future. Others with opposing directions of effects may reveal potential adverse side effects for consideration in therapeutic development (e.g. with anti-IL6 biologics for RA).

## Conclusions

Through our ADGAPP pipeline, we have developed an integrated approach which has allowed semi-quantification of the subjective process of prioritisation of genes (and variants) at AD GWAS loci. We have amassed 103 datasets (including molecular datasets from relevant tissues) and used these to generate prioritisation scores for each gene at the 25 established European AD GWAS loci. At eight loci we were able to prioritise single candidate genes with good evidence, many not being the closest gene to the GWAS index SNP. For 6 loci we identify genes not previously implicated as the top-ranked candidates. We present comprehensive results and discussion of the evidence at each locus to enable appropriate follow-up molecular biology investigations, including drug target discovery of these potential causal genes. The principles behind the ADGAPP pipeline can be adopted in annotating GWAS on other disease in the future.

**Additional File 1: Supplementary Text**. Description of ADGAPP scoring system.

**Additional File 2: Supplementary Figure 1**. Overview of contribution of different type of -omics evidence to gene and SNP ranking in all the AD GWAS loci. Each bar represents a number of individual studies in a given category contributing to the sum total of evidence.

**Additional File 3: Supplementary Figure 2**. Distribution of scores obtained in our model for candidate gene ranking at each locus (labelled with lead SNP).

**Additional File 4: Supplementary Figure 3**. Distribution of scores obtained in our model for causal SNP ranking at each locus (labelled with lead SNP).

**Additional File 5: Supplementary Table 1**. Colocalisation results from coloc and TWAS methods (either base TWAS or TWAS-based coloc) on AD GWAS and eQTLs in tissues from a number of datasets. We report coloc results for genes with posterior probability of a shared causal eQTL and GWAS variant (PPH4) > 0. 5 for any gene with at least one strong colocalisation result of PPH4 > 0.9, while for TWAS we show genes with genome-wide significant and independent colocalisation evidence in conditional and joint analysis. Gene rank at the given locus in our final GWAS gene prioritisation model is also given.

**Additional File 6: Supplementary Table 2**. Results meeting the genome-wide significance threshold with no significant heterogeneity in HEIDI analysis for SMR test of GTEx, Blueprint and eQTLGen eQTL instruments against AD GWAS. Gene rank at the given locus in our final GWAS gene prioritisation model is also given.

**Additional File 7: Supplementary Text 2**. ADGAPP prioritisation summary for AD loci.

**Additional File 8: Supplementary Figure 4**. Score by types of evidence in all the AD GWAS loci. Scores for top 3 ranked genes at each locus are shown partitioned by category of evidence – here including the top 10 categories contributing the highest proportion of total score at the top 10 ranked genes for all loci. Order of loci corresponds to the order in Table 1.

**Additional File 9. Supplementary Table 3**. Genes previously implicated in AD, not prioritised by ADGAPP.

**Additional File 10: Supplementary Table 4**. Comparison of Enrichr-based significant enrichment testing results for: top 3 genes combined from across all the AD GWAS loci tested (Genes.top_3) and other known AD genes (Genes.previously_known), against select ontologies.*List of abbreviations* AD: atopic dermatitis; ADGAPP: Atopic Dermatitis GWAS Annotation Prioritisation Pipeline; caQTL - chromatin accessibility quantitative trait locus; AS: ankylosing spondylitis; CD: Crohn’s disease; DGE: differential gene expression; eQTL: expression quantitative trait locus; GTEx: Genotype-Tissue Expression project; GWAS: genome-wide association study; hQTL: histone quantitative trait locus; LCL – lymphoblastoid cell line; LD: linkage disequilibrium; MHC: major histocompatibility complex; mQTL: DNA methylation quantitative trait locus; MS: multiple sclerosis; PP: posterior probability; pQTL: protein quantitative trait locus; RA: rheumatoid arthritis; SLE: Systemic lupus erythematosus; SNP: single nucleotide polymorphisms; sQTL: splicing quantitative trait locus; T1D: type I diabetes; UC: ulcerative colitis

## Supporting information

Additional Files Titles & legends

Consortium members

## Data Availability

All the code written to carry out the ADGAPP analysis is available on GitHub at: https://github.com/MRCIEU/eczema_gwas_fu
Code is archived under: https://doi.org/10.5281/zenodo.3775865
The datasets supporting the conclusions of this article are available in the Figshare repository.

https://doi.org/10.5281/zenodo.3775865

https://doi.org/10.6084/m9.figshare.12222731

https://doi.org/10.6084/m9.figshare.12130701

https://doi.org/10.6084/m9.figshare.12221006

https://doi.org/10.6084/m9.figshare.12221033

https://doi.org/10.6084/m9.figshare.12130824

https://doi.org/10.6084/m9.figshare.12130857

https://doi.org/10.6084/m9.figshare.12130863

https://doi.org/10.6084/m9.figshare.12130878

## Declarations

### Ethics approval and consent to participate

Not applicable

### Consent for publication

Not applicable

### Availability of data and materials

All the code written to carry out the ADGAPP analysis is available on GitHub at: https://github.com/MRCIEU/eczema_gwas_fu Code is archived under: https://doi.org/10.5281/zenodo.3775865

The datasets supporting the conclusions of this article are available in the Figshare repository.

**Dataset S1**. Reference table detailing individual datasets used in ADGAPP. https://doi.org/10.6084/m9.figshare.12222731

**Dataset S2**. The final evidence dataset used in ranking genes and variants: https://doi.org/10.6084/m9.figshare.12130701

**Dataset S3**. Locus Zoom-style gene and variant score plots for each locus: Gene: https://doi.org/10.6084/m9.figshare.12221006

Variant: https://doi.org/10.6084/m9.figshare.12221033

**Dataset S4**. Full gene and variant loci rankings:

Gene: https://doi.org/10.6084/m9.figshare.12130824 Variant: https://doi.org/10.6084/m9.figshare.12130857

**Dataset S5**. Data evidence for top 10-ranked gene and variants at each locus: Gene: https://doi.org/10.6084/m9.figshare.12130863

Variant: https://doi.org/10.6084/m9.figshare.12130878

### Competing interests

TRG receives funding from GlaxoSmithKline and Biogen for unrelated research.

### Funding

This work was supported by a Springboard award (SBF003\1094), awarded to LP, supported by the Academy of Sciences (AMS), the Wellcome Trust, the Government Department of Business, Energy and Industrial Strategy (BEIS) and the British Heart Foundation (BHF). TRG acknowledges support from the UK Medical Research Council (MC_UU_00011/4). TGR is a UKRI Innovation research fellow (MR/S003886/1). TwinsUK is funded by the Wellcome Trust, Medical Research Council, European Union, Chronic Disease Research Foundation (CDRF), Zoe Global Ltd and the National Institute for Health Research (NIHR)-funded BioResource, Clinical Research Facility and Biomedical Research Centre based at Guy’s and St Thomas’ NHS Foundation Trust in partnership with King’s College London.

### Authors’ contributions

LP initiated the study, LP, MKS and TRG contributed to the design of the study. MKS and TGR performed the analysis of the data. MKS wrote the manuscript with input from LP and TRG. All authors read and approved the manuscript before submission.

## Acknowledgements

We acknowledge the many open access datasets that have made this work possible. We also acknowledge eQTLGen for providing eQTL results before their publication and TwinsUK (http://www.twinsuk.ac.uk/data-access/publications/) for providing access to data for eQTL analysis. Fine-mapping analysis used the UK Biobank genetic data resource as an LD reference [Application #10074].

## Notes

### Author Declarations

No ethics approval was required.

