## Additional File 1 Supplementary Text for "Triangulating molecular evidence to prioritise candidate causal genes at established atopic dermatitis loci"

**Standardised results table**

Results from individual analyses and lookups were summarised into tables (available in Dataset S2) with the following columns, with required values marked by *:

**table id***	Particular analysis/ experiment in the study

**study id***	Unique study which data was sourced from. See Supp Table 3 for references

**index SNP***	Index SNP representing a genetic locus harbouring AD GWAS hit

**current SNP**	rsid ID of a SNP within the GWAS genetic locus

**gene name**	Gene associated with current SNP or prioritized by other non SNP-based methods, such as DGE (differential gene expression).

**FDR/p-value/posterior probability/score/beta**	A value indicating confidence in true association and/or magnitude of the effect. Note that results used in the analysis were preselected on being significant in the original analysis in the first place, with the exception of scores, which provided a continuous measure of variant deleteriousness.

**significance threshold** Value threshold for the result to be included in contributing to evidence score

**effect allele**	Effect allele at a given SNP, when available

**tissue**	Target tissue investigated in a given experiment

**sample size**	Sample size of a given experiment

**study type***	Type of experiment (e.g. ChiP-Seq) or analysis (e.g. eQTL)

**cis/trans**	Interaction in cis- or trans- (for QTLs)

**evidence weight***	Subjective prior belief in evidence strength, from 1 (highest) to 3	(lowest) – see main Methods

**number of SNP significant values***	Total number of unique variants in a given locus found among significant hits in the given analysis/experiment

**number of gene significant values***	Total number of unique genes in a given locus found among significant hits in the given analysis/experiment

**n experiments***	Number of analyses/ experiments in the study

**Calculation of basic score (per gene or variant in a given experiment/analysis):**

$p$ is measure of result strength (in the order of preference: False Discovery Rate (FDR), p-value, posterior probability (PP), score, beta; $p_{max}$ is the top lowest (for FDR or p-value) or top absolute highest (PP, score, beta) value present in the dataset

$b= 1+\frac{p}{\sqrt{{log}_{10}(p_{max})}}$ when $p$ refers to FDR or p-value

$b=1+\frac{p}{p_{max}}$ when $p$ refers to PP, score or beta value

**Calculation of total score (per gene or variant across all experiments/analyses):**

*gene ranking:*

$$\sum_{all b} \frac{b*v}{\sqrt{n}*g* \sqrt{s}}* \sqrt{\frac{\left( t + i \right)}{2}}$$

*variant ranking:*

$$\sum_{all b} \frac{b*v}{\sqrt{n}*s}* \sqrt{\frac{\left( t + i \right)}{2}}$$

*v* – evidence weight adjustment: *v* = 1 for evidence weight of 3 (lowest credibility), *v* = 2 for evidence weight of 2, *v* = 20 for evidence weight of 1 (highest credibility)

$n$– number of experiments in a given study

$g$ – number of genes in a given interval in a given experiment/analysis

$s$ – number of SNPs in a given interval in a given experiment/analysis

$t$ – number of unique study types showing evidence for a given variant/gene

$i$ – number of unique study IDs showing evidence for a given variant/gene
