## Supplementary figures and images for "Triangulating molecular evidence to prioritise candidate causal genes at established atopic dermatitis loci"

### Additional File 2 Supplementary Figure 1

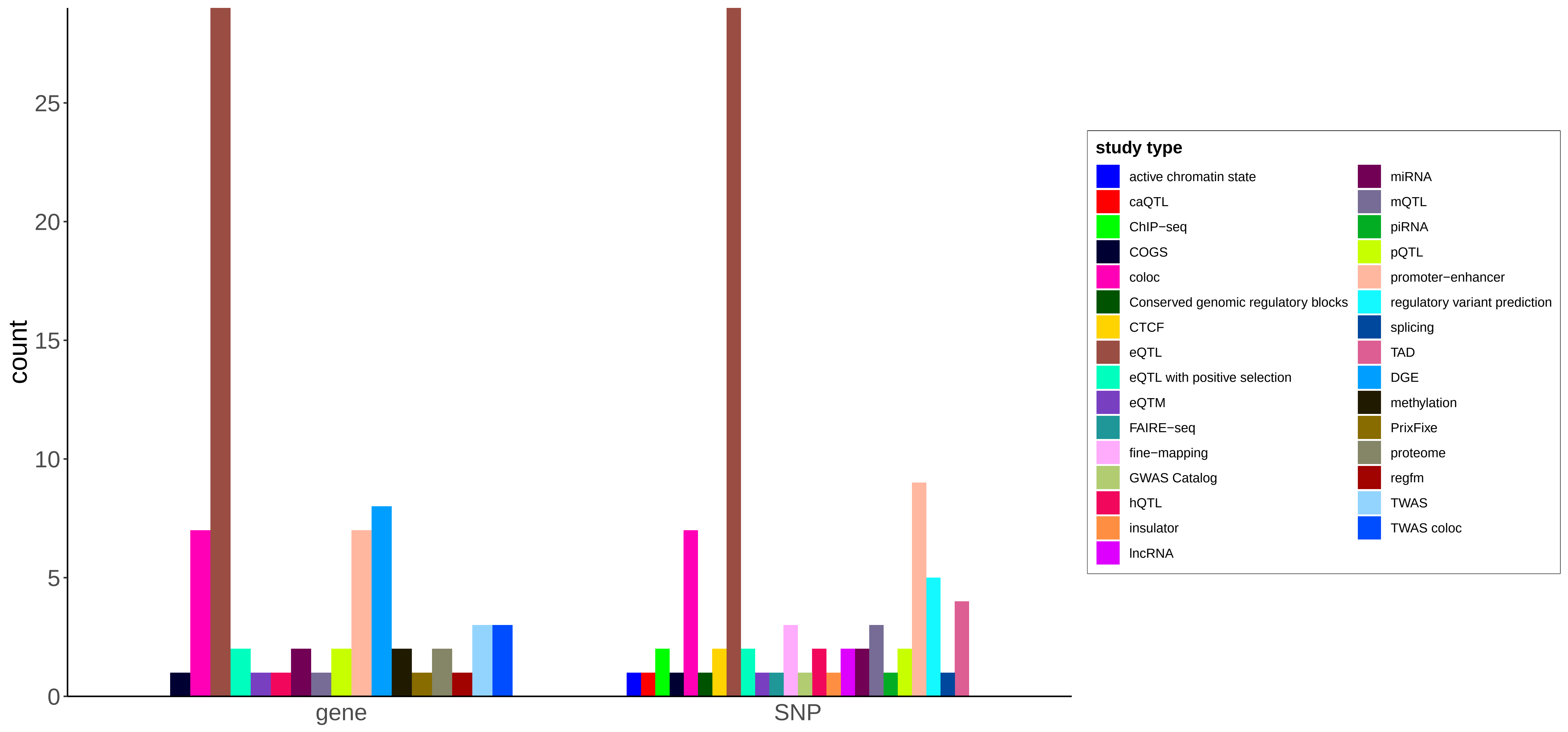
