## Additional File 3 Supplementary Figure 2 for "Triangulating molecular evidence to prioritise candidate causal genes at established atopic dermatitis loci"

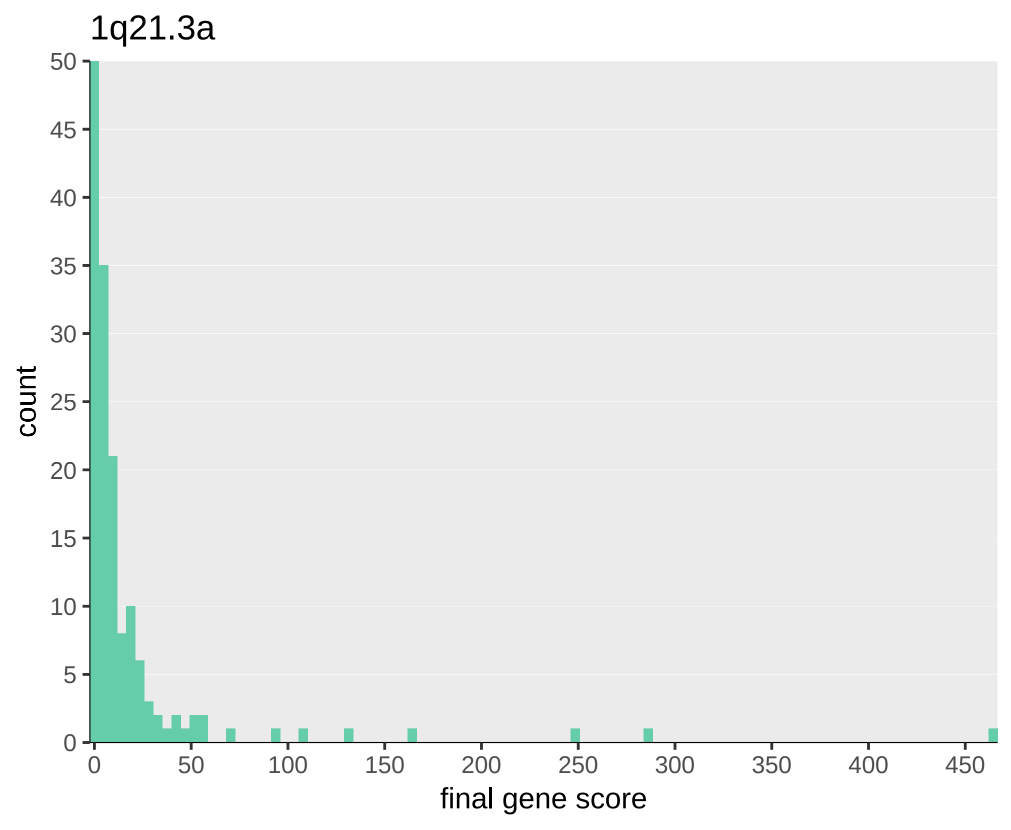
1q21.3a
count
final gene score
0
5
10
15
20
25
30
35
40
45
50
0
50
100
150
200
250
300
350
400
450

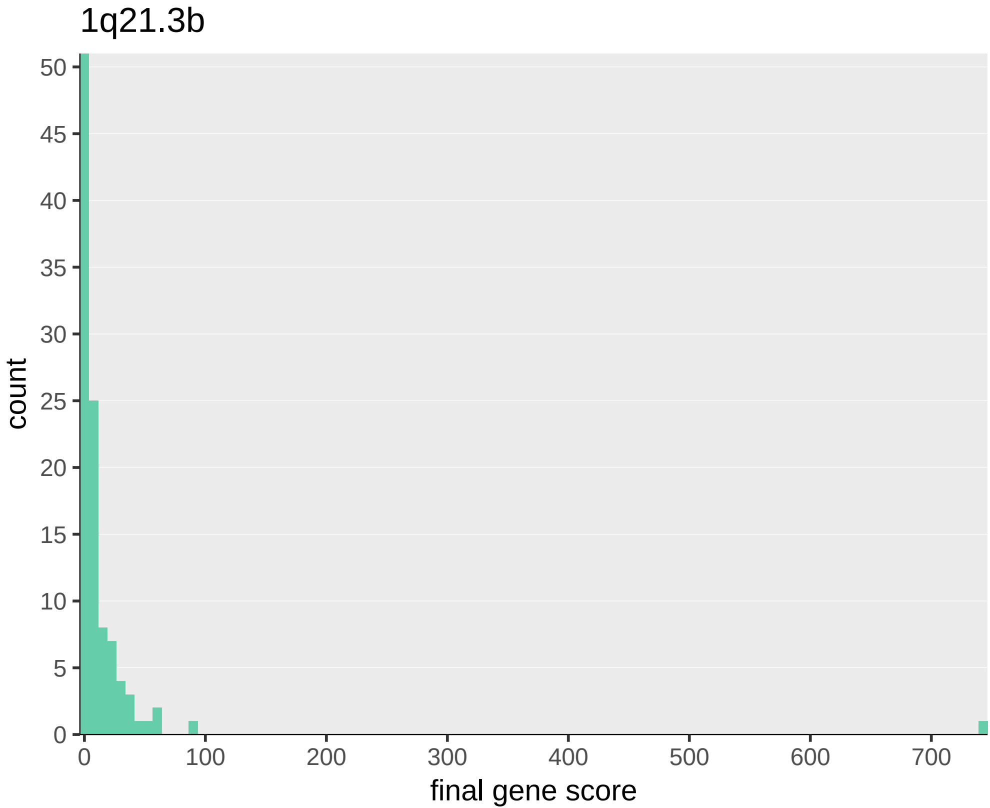
1q21.3b
count
final gene score
0
5
10
15
20
25
30
35
40
45
50
0
100
200
300
400
500
600
700

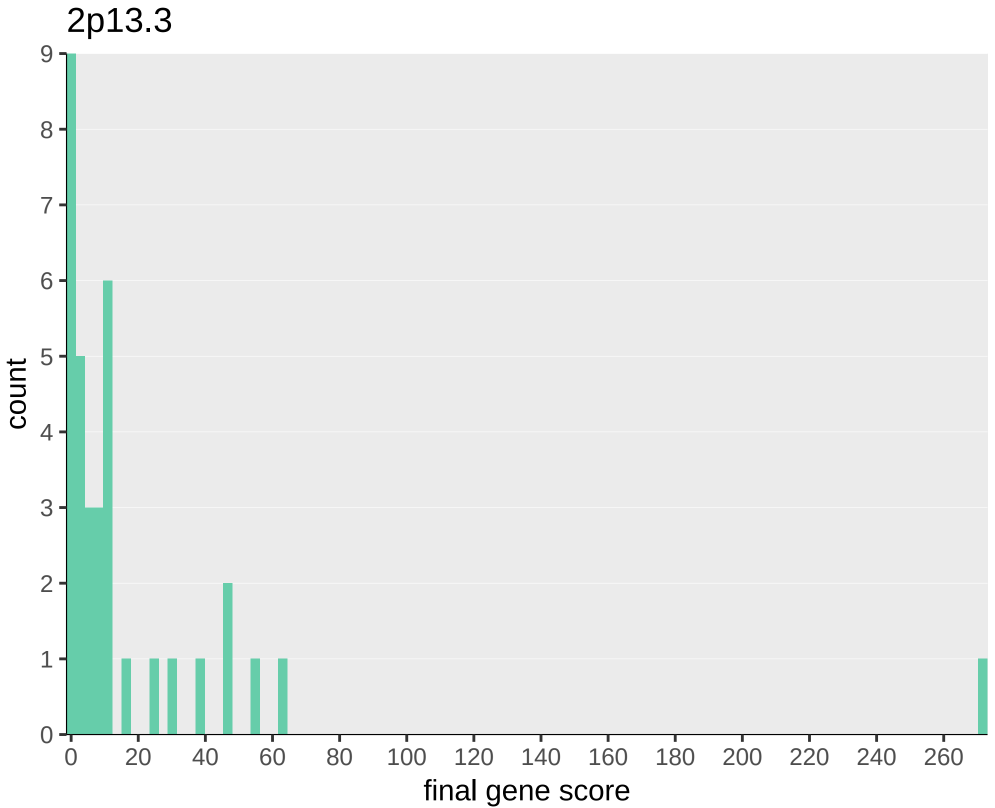
2p13.3
count
9
8
7
6
5
4
3
2
1
0
0
20
40
60
80
100
120
140
160
180
200
220
240
260
final gene score

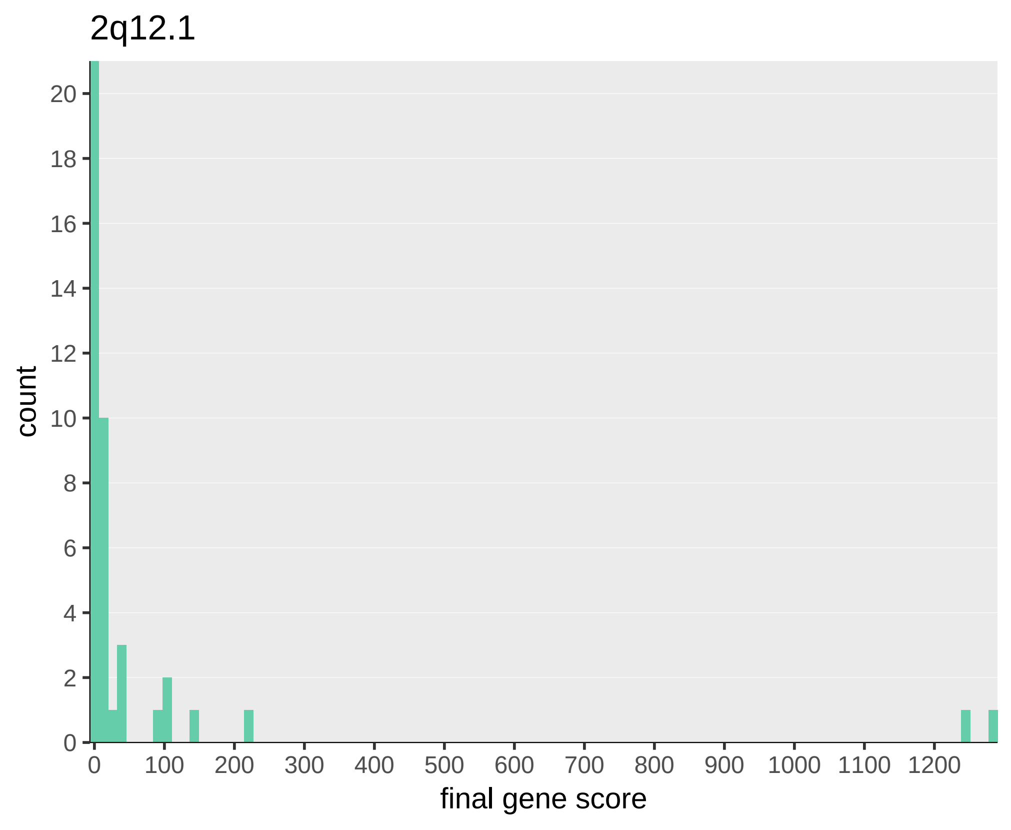
2q12.1
count
20
18
16
14
12
10
8
6
4
2
0
0
100
200
300
400
500
600
700
800
900
1000
1100
1200
final gene score

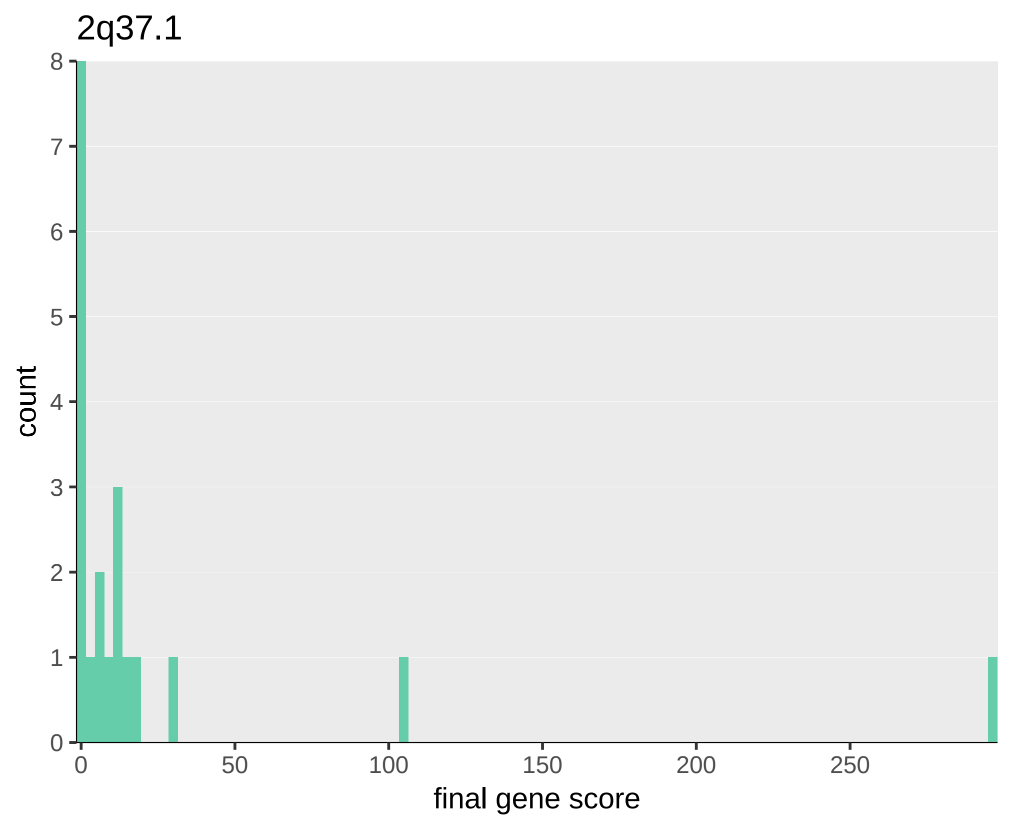
2q37.1
count
final gene score
0
1
2
3
4
5
6
7
8
0
50
100
150
200
250

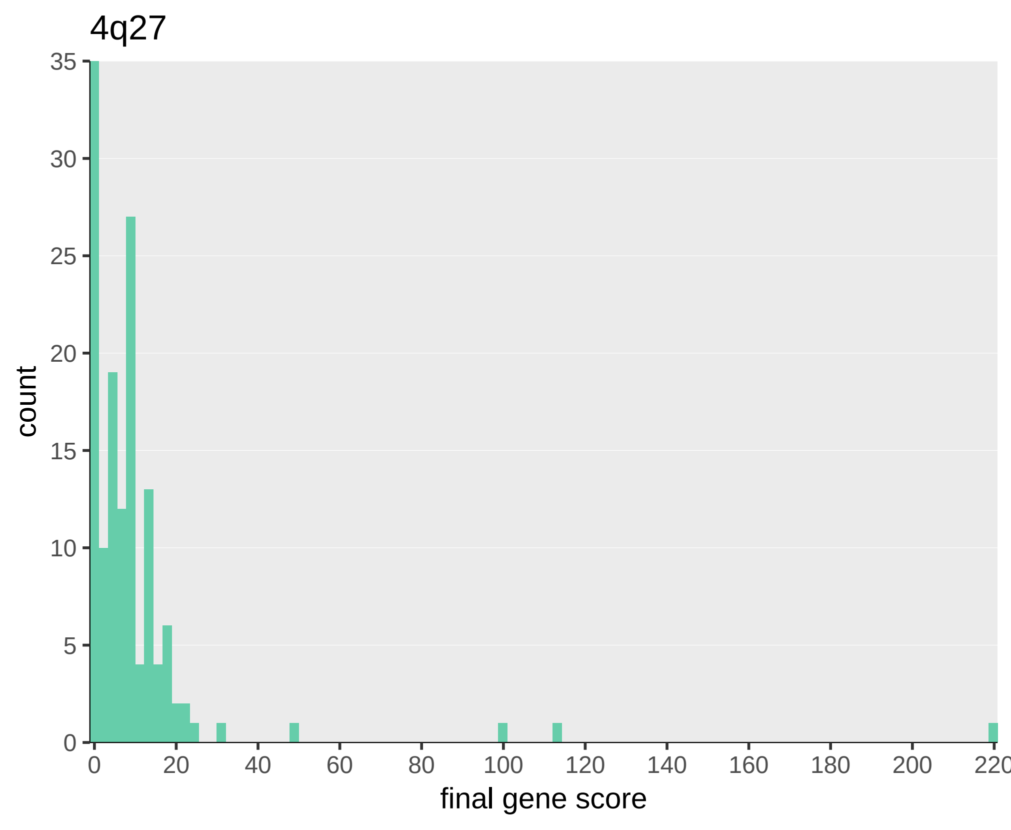
4q27
count
35
30
25
20
15
10
5
0
0
20
40
60
80
100
120
140
160
180
200
220
final gene score

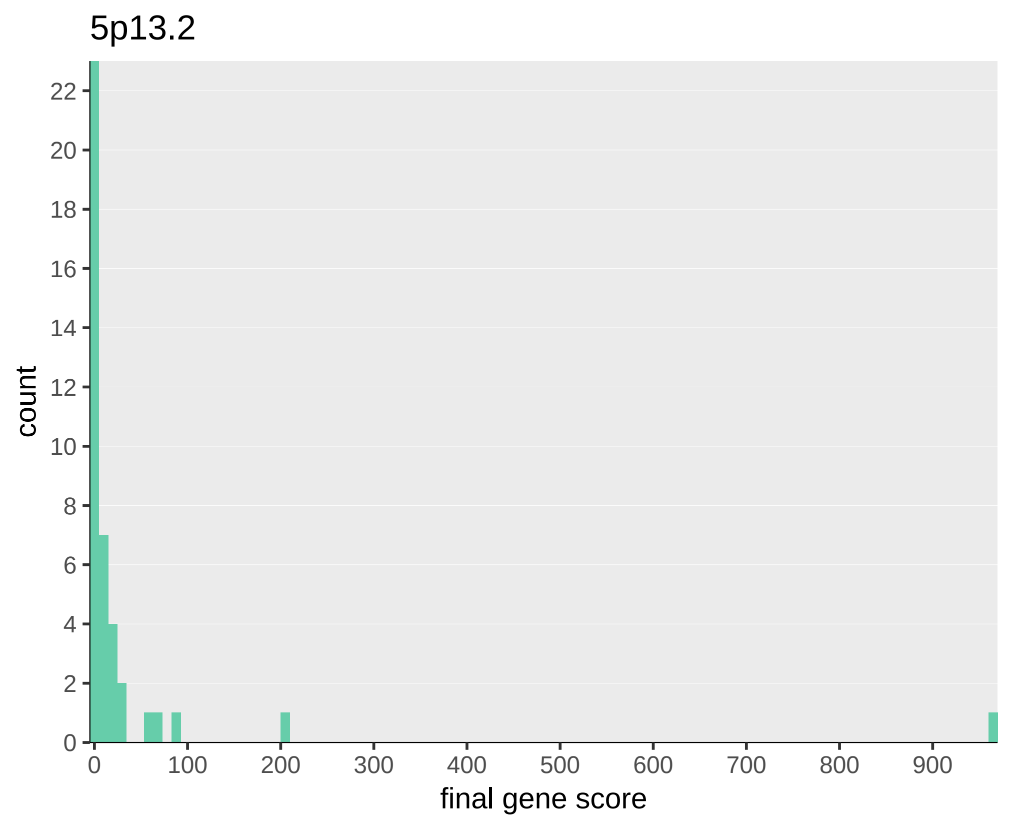
5p13.2
count
final gene score
0
2
4
6
8
10
12
14
16
18
20
22
0
100
200
300
400
500
600
700
800
900

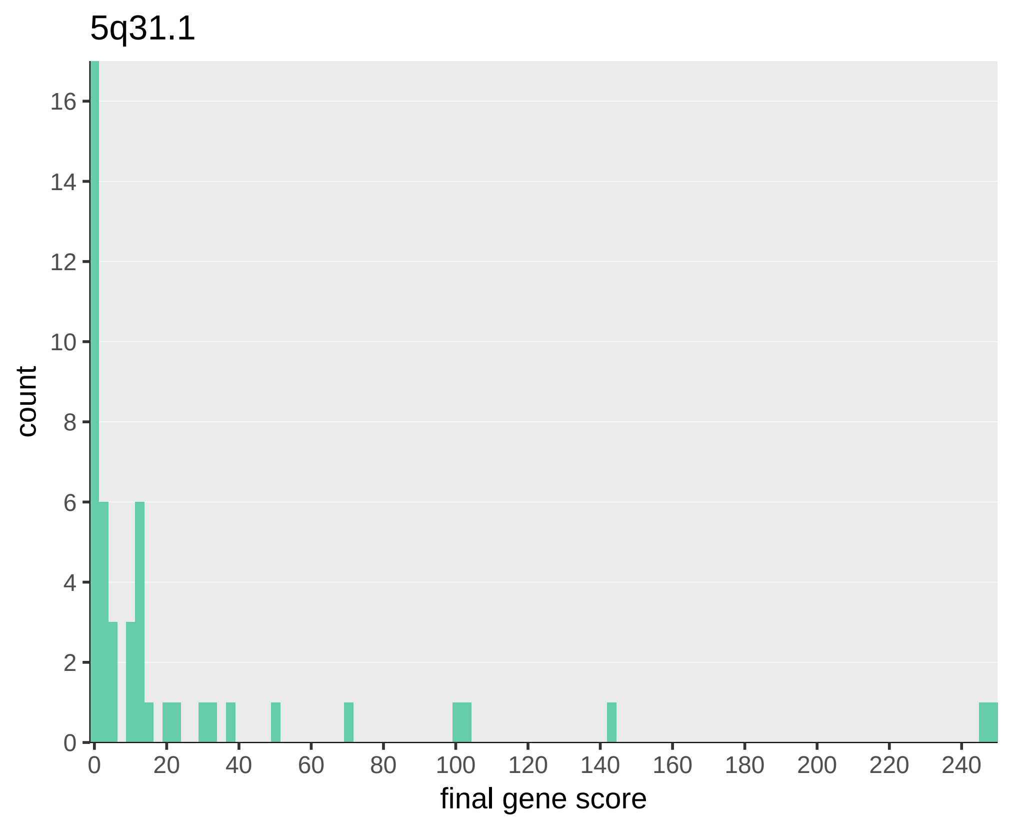
5q31.1
count
16
14
12
10
8
6
4
2
0
0
20
40
60
80
100
120
140
160
180
200
220
240
final gene score

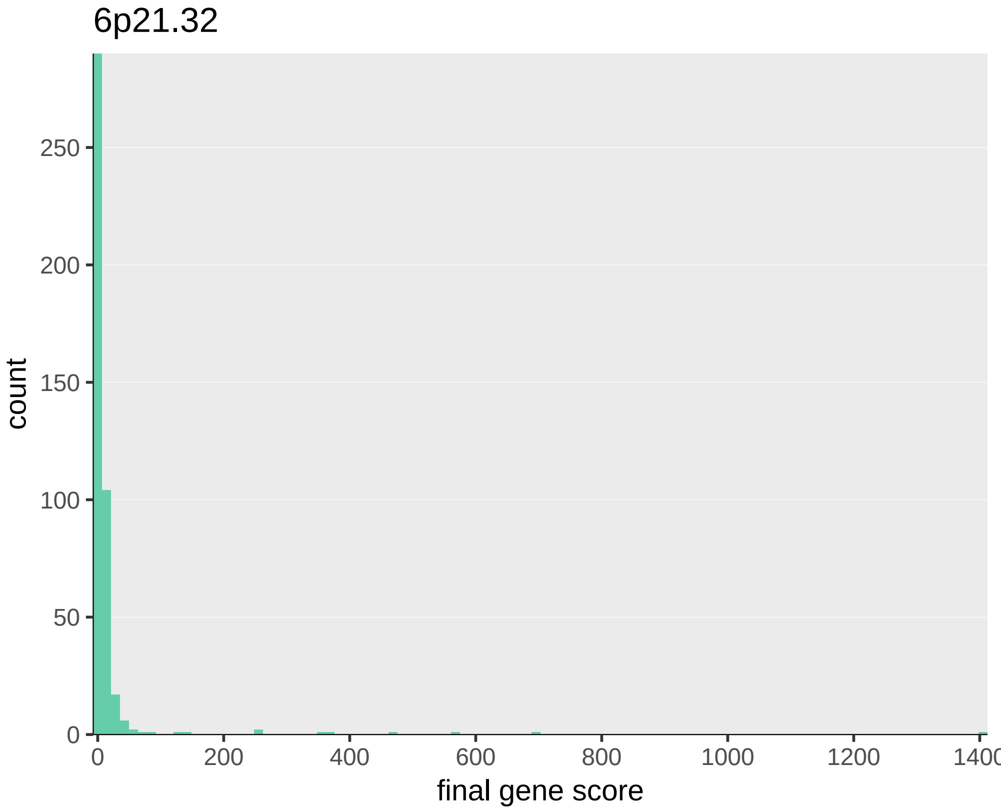
6p21.32
count
250
200
150
100
50
0
0
200
400
600
800
1000
1200
1400
final gene score

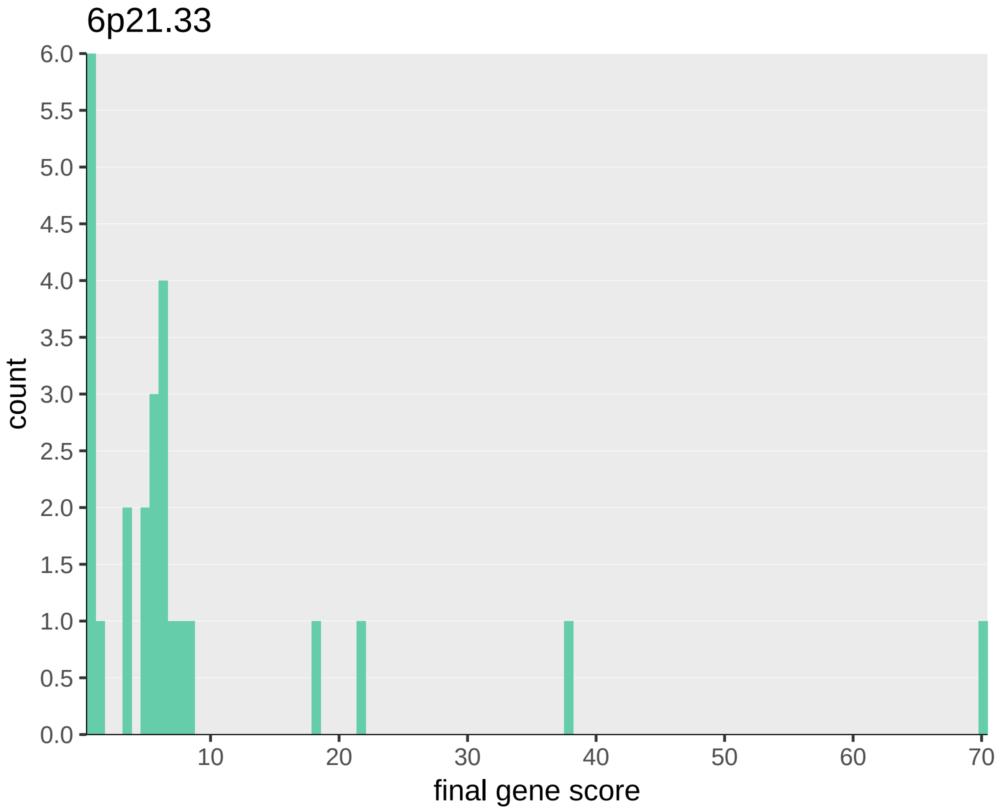

6p21.33
count
final gene score
6.0
5.5
5.0
4.5
4.0
3.5
3.0
2.5
2.0
1.5
1.0
0.5
0.0
10
20
30
40
50
60
70

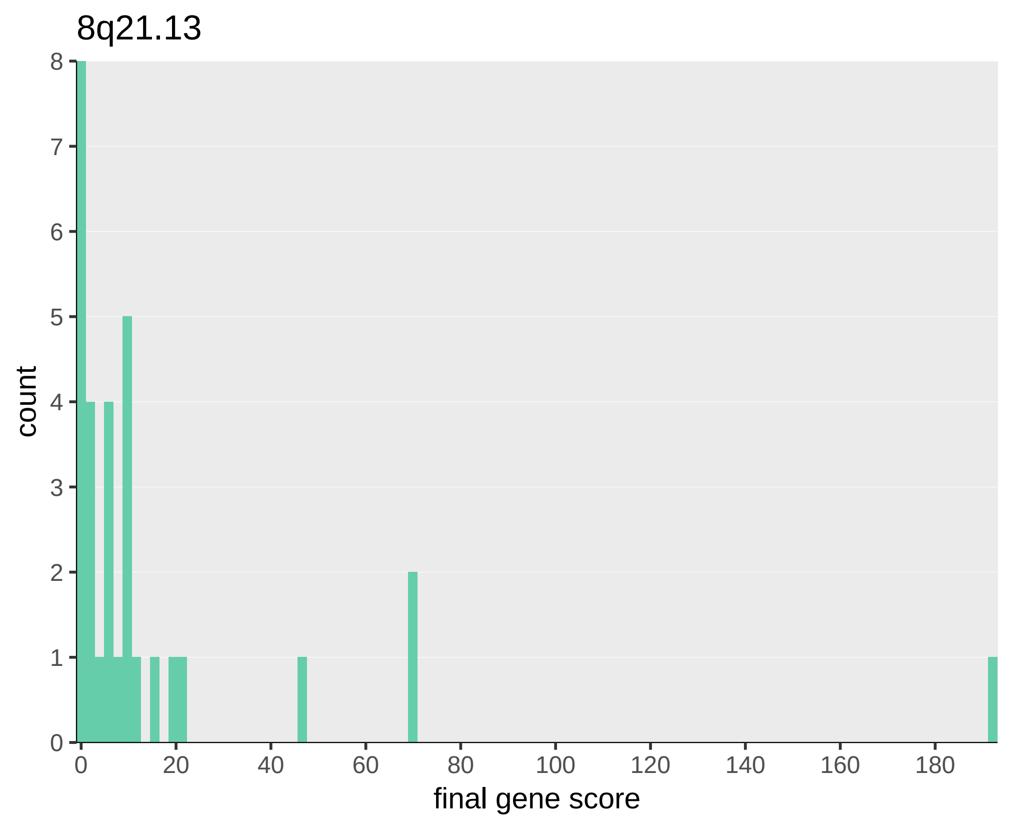
8q21.13
count
0
1
2
3
4
5
6
7
8
0
20
40
60
80
100
120
140
160
180
final gene score

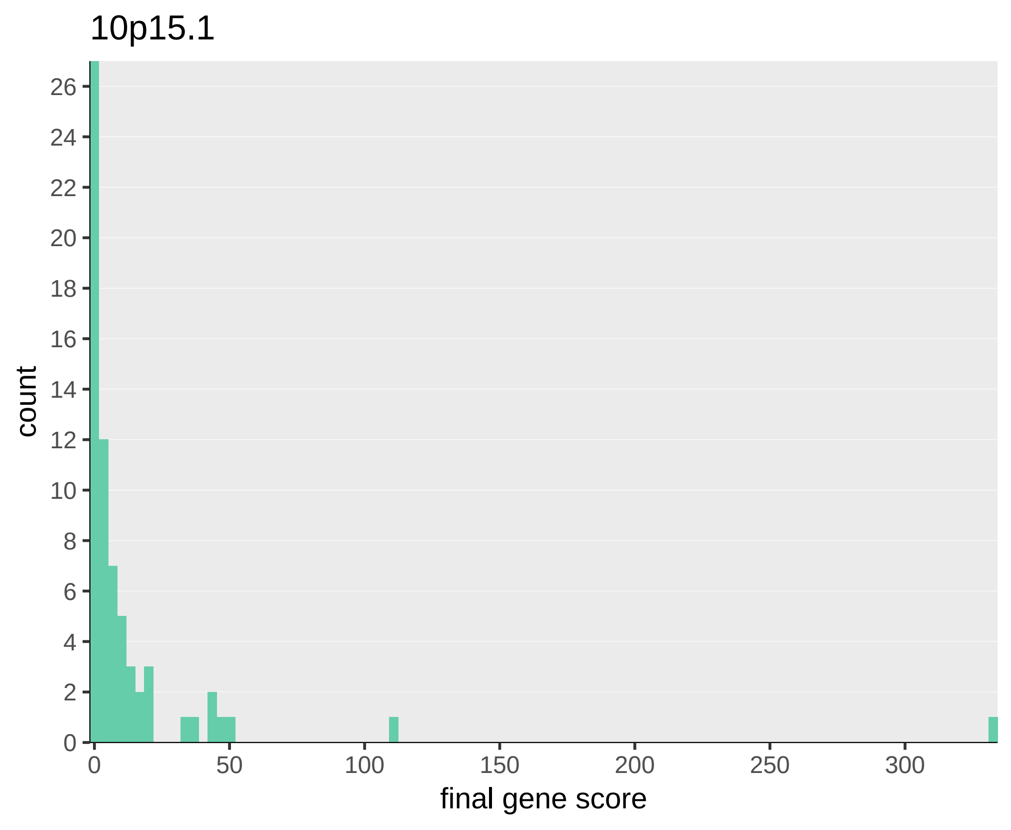
10p15.1
count
0
2
4
6
8
10
12
14
16
18
20
22
24
26
0
50
100
150
200
250
300
final gene score

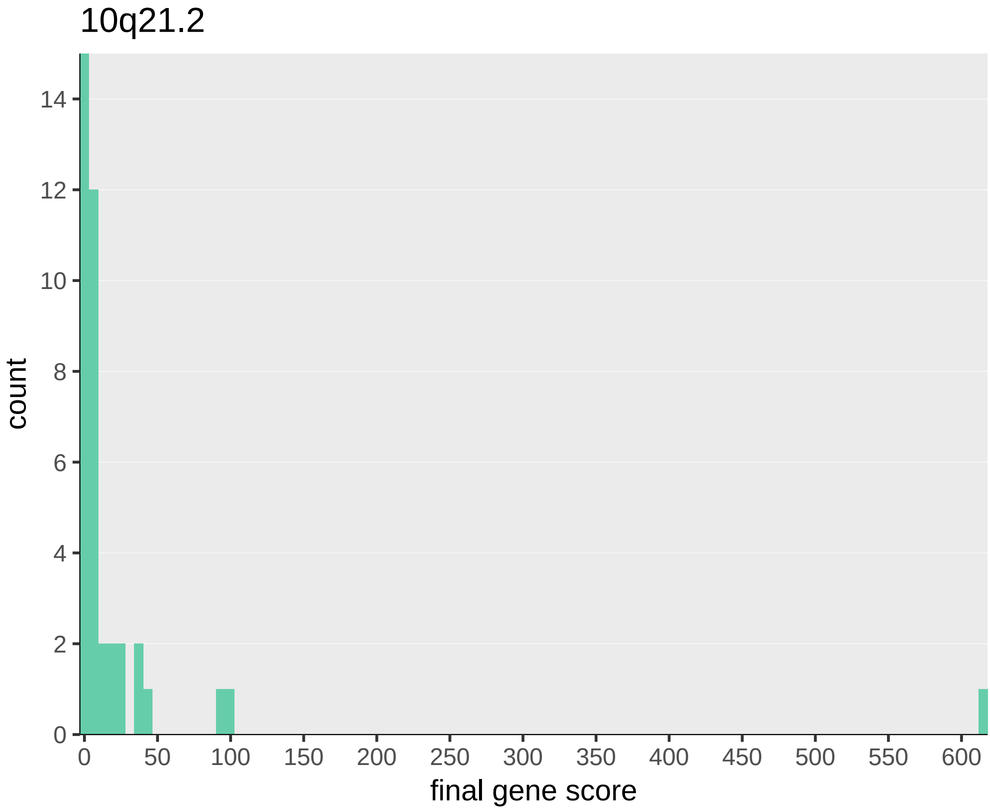
10q21.2
count
final gene score
0
2
4
6
8
10
12
14
0
50
100
150
200
250
300
350
400
450
500
550
600

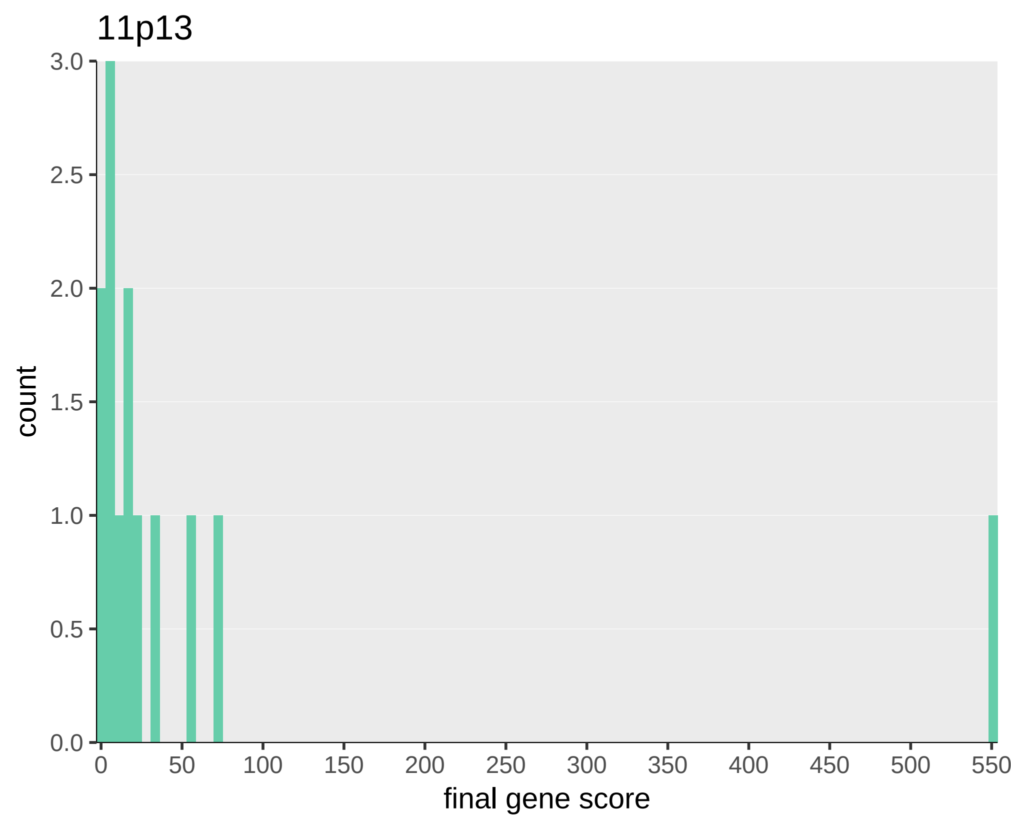
11p13
count
3.0
2.5
2.0
1.5
1.0
0.5
0.0
0
50
100
150
200
250
300
350
400
450
500
550
final gene score

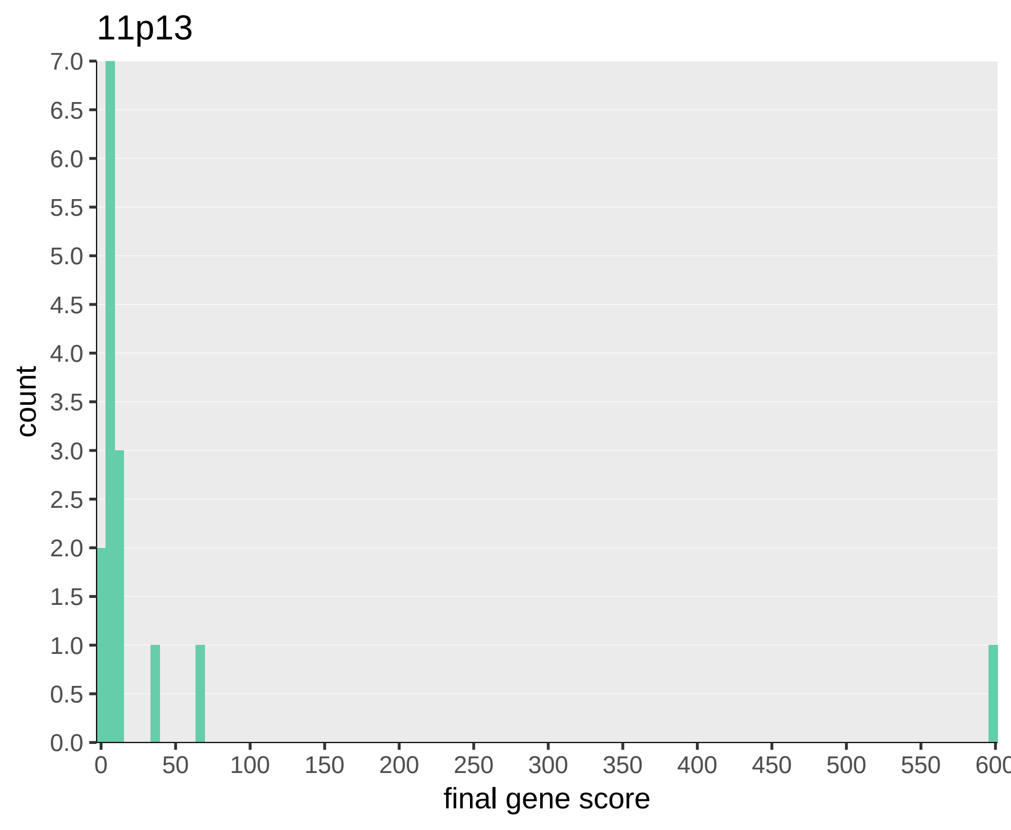
11p13
count
7.0
6.5
6.0
5.5
5.0
4.5
4.0
3.5
3.0
2.5
2.0
1.5
1.0
0.5
0.0
0
50
100
150
200
250
300
350
400
450
500
550
600
final gene score

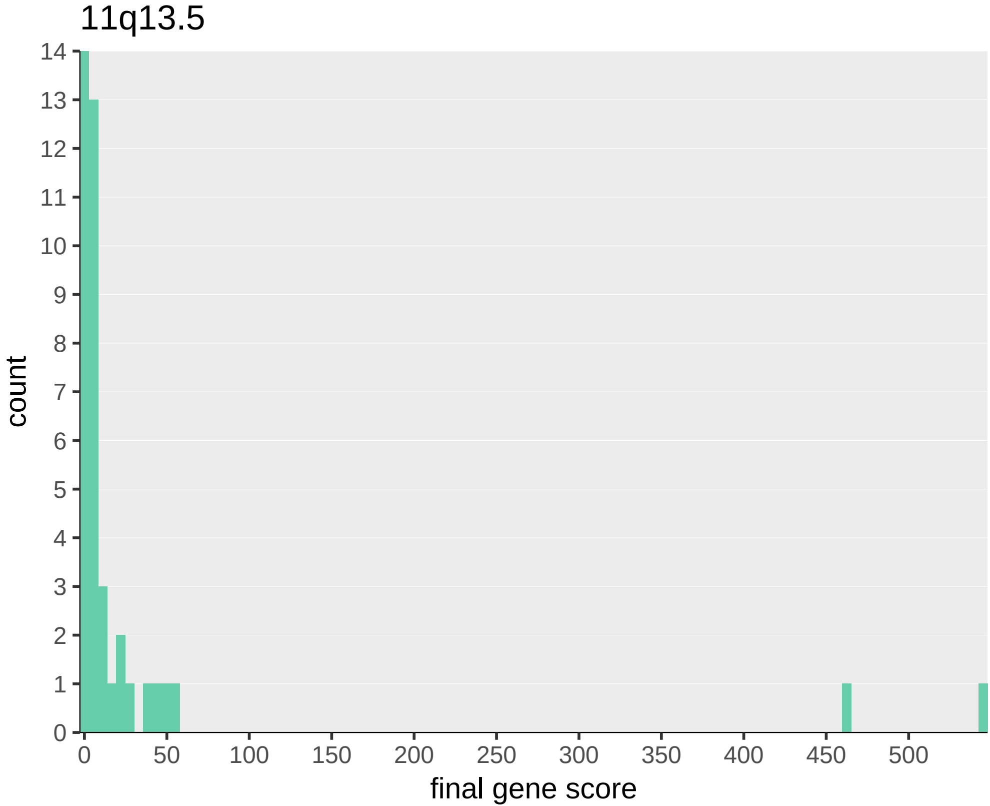
11q13.5
count
final gene score
0
50
100
150
200
250
300
350
400
450
500

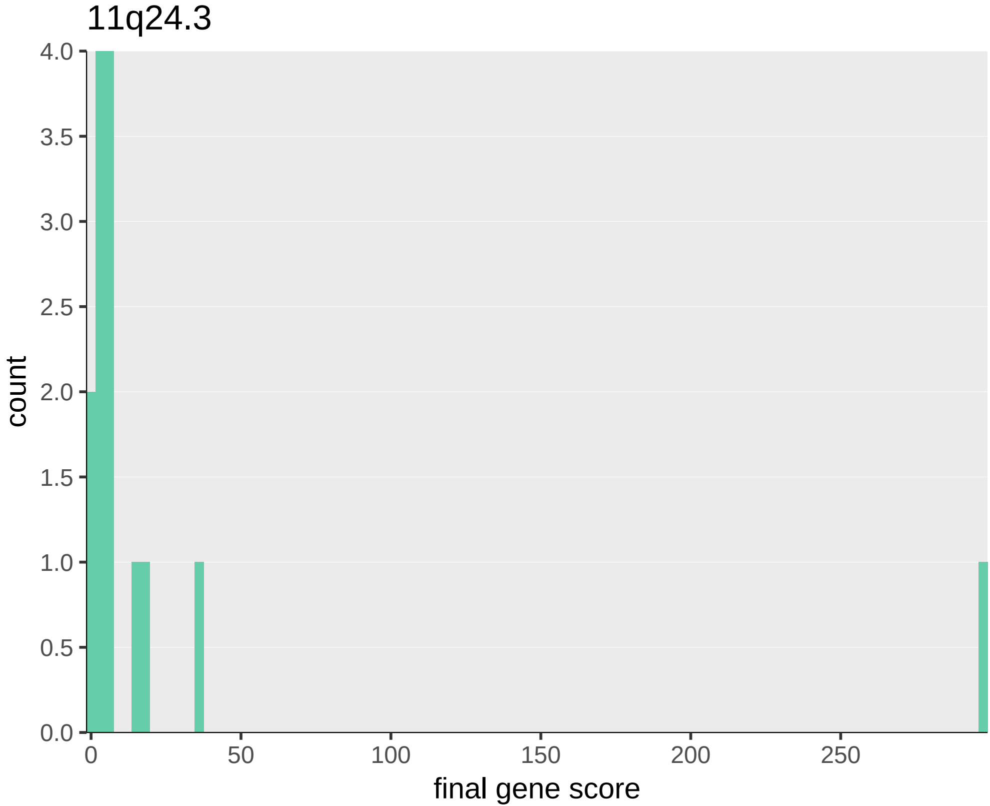
11q24.3
count
final gene score
4.0
3.5
3.0
2.5
2.0
1.5
1.0
0.5
0.0
0
50
100
150
200
250

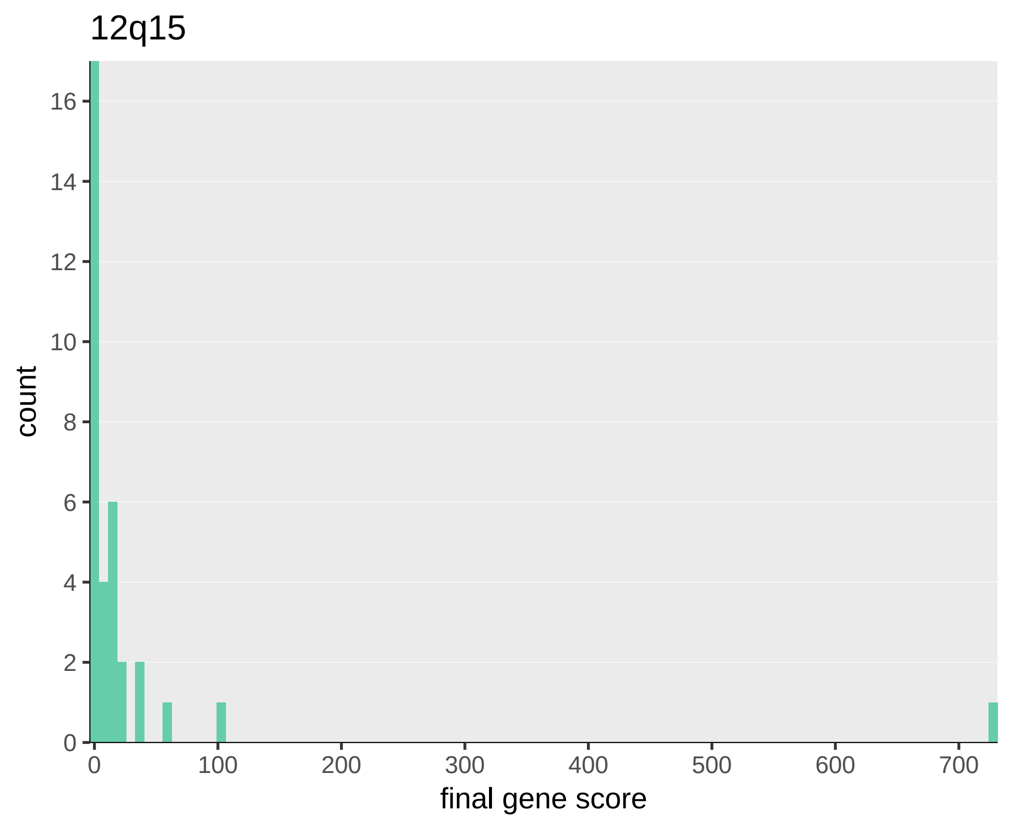
12q15
count
final gene score
0
2
4
6
8
10
12
14
16
0
100
200
300
400
500
600
700

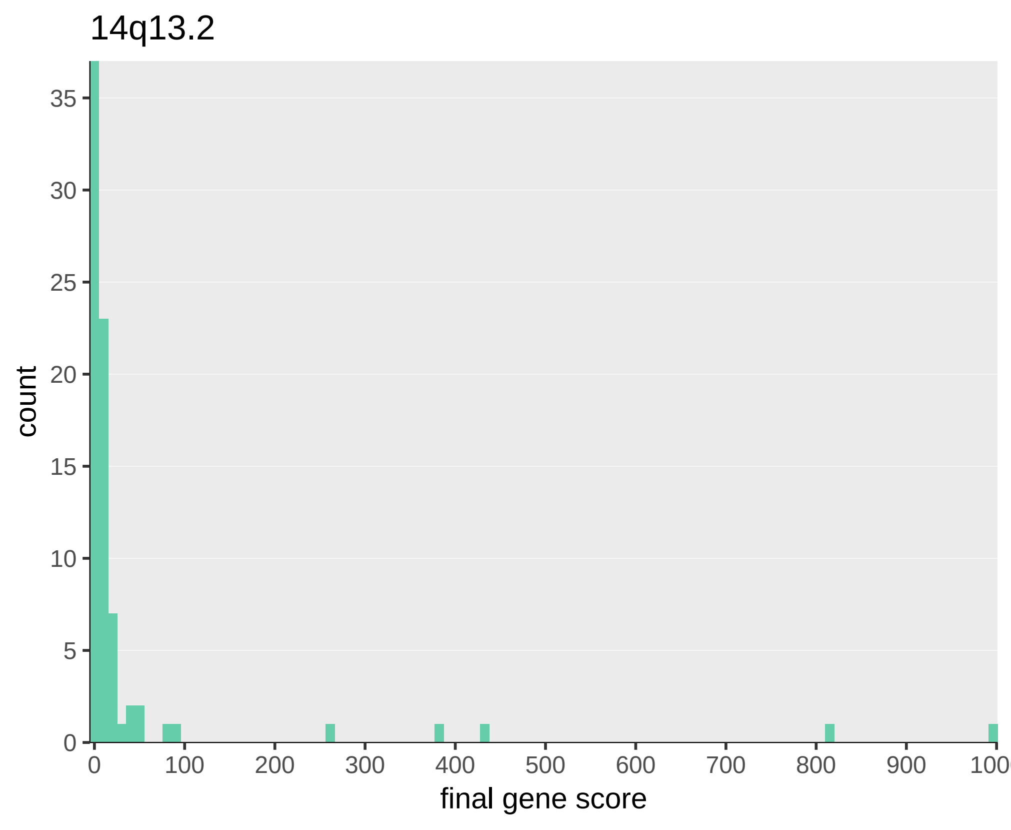
14q13.2
count
35
30
25
20
15
10
5
0
0
100
200
300
400
500
600
700
800
900
1000
final gene score

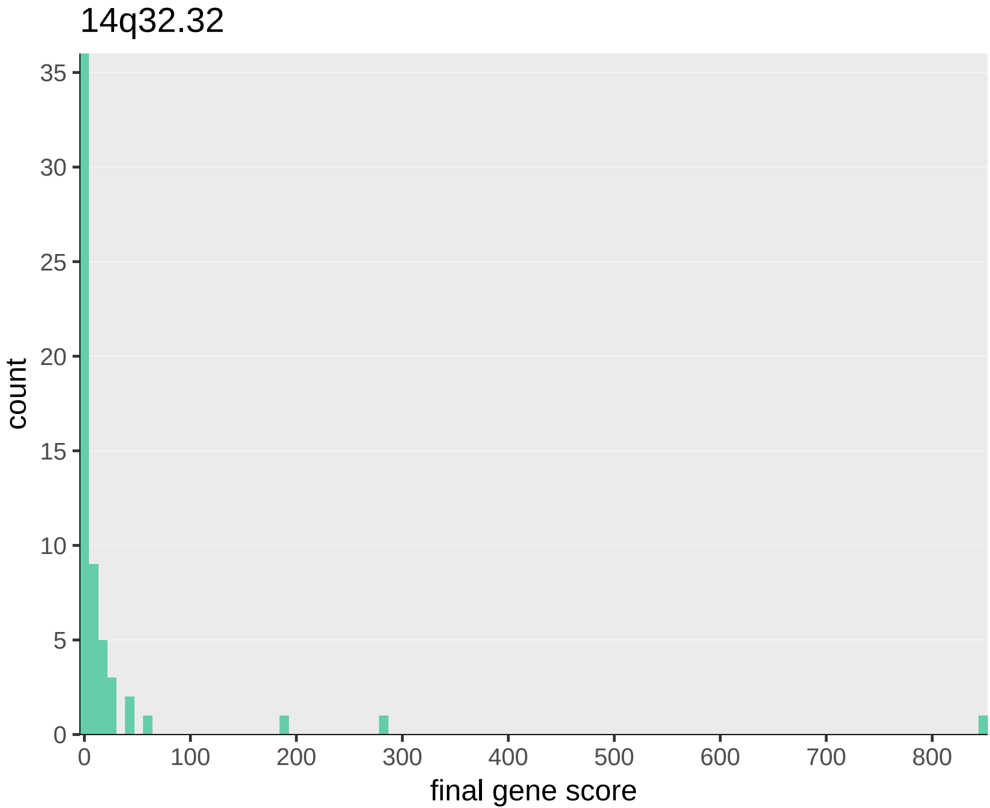
14q32.32
count
35
30
25
20
15
10
5
0
0
100
200
300
400
500
600
700
800
final gene score

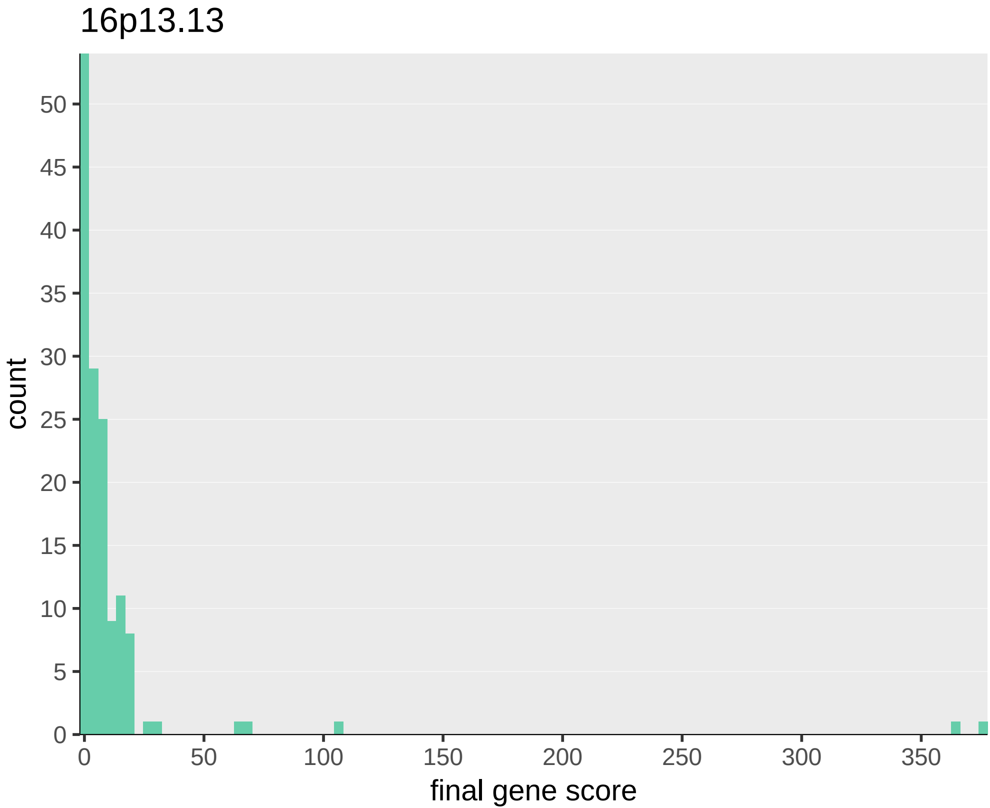
16p13.13
count
final gene score
0
5
10
15
20
25
30
35
40
45
50
0
50
100
150
200
250
300
350

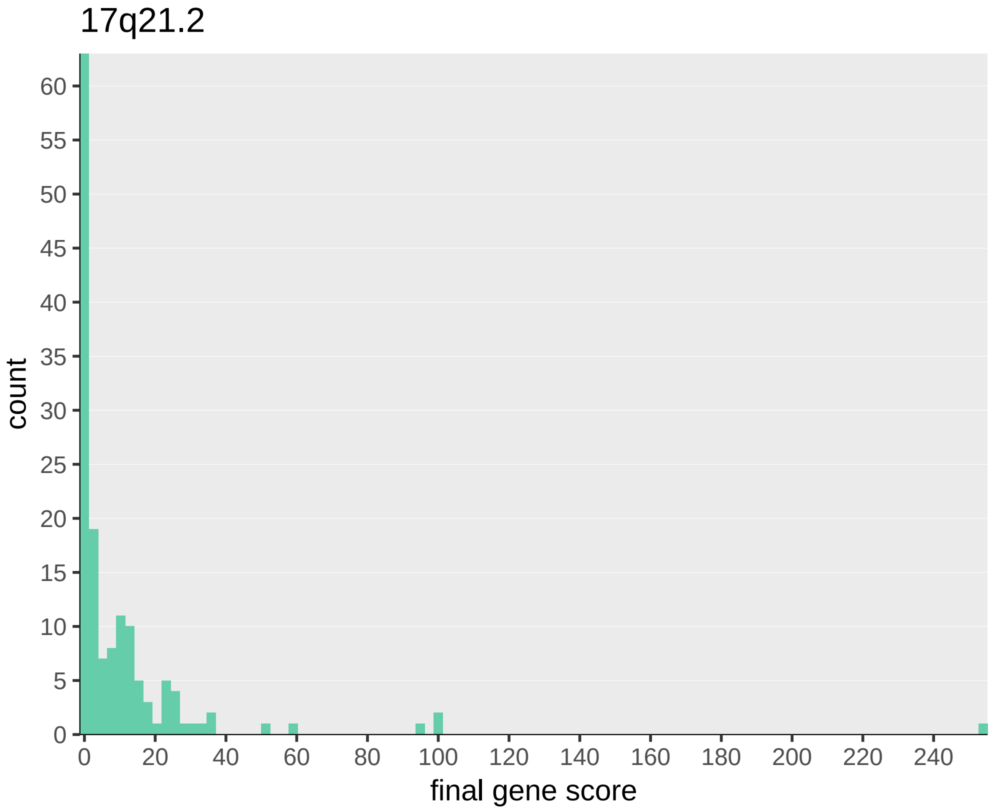
17q21.2
count
60
55
50
45
40
35
30
25
20
15
10
5
0
0
20
40
60
80
100
120
140
160
180
200
220
240
final gene score

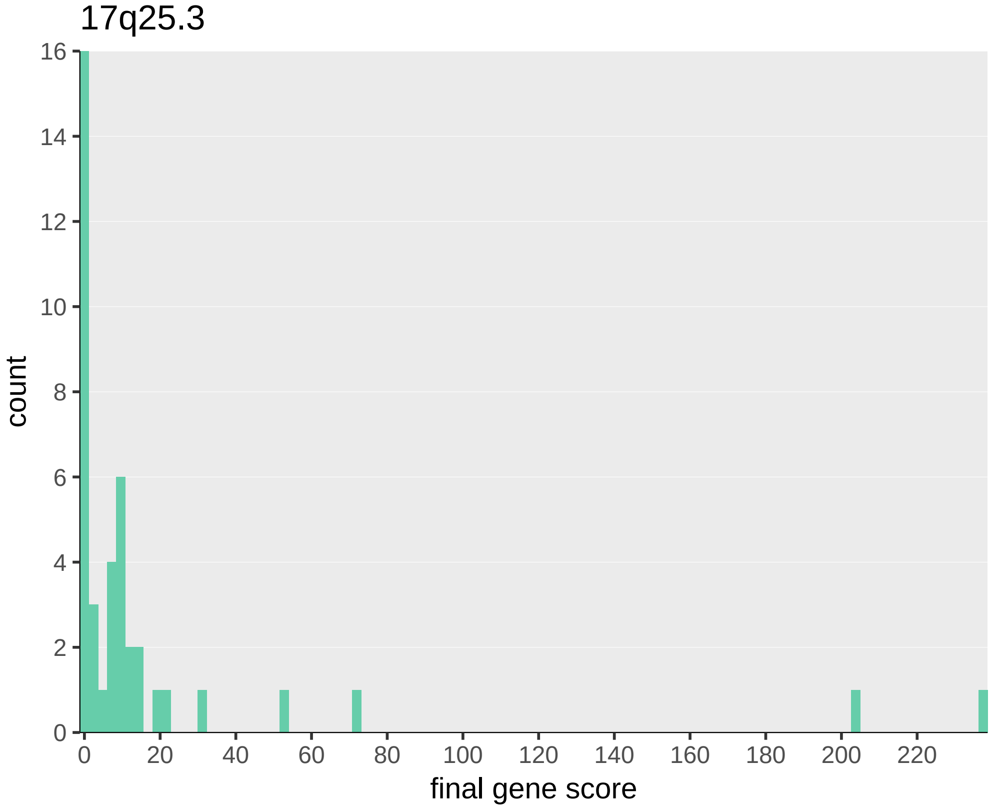
17q25.3
count
final gene score
0
2
4
6
8
10
12
14
16
0
20
40
60
80
100
120
140
160
180
200
220

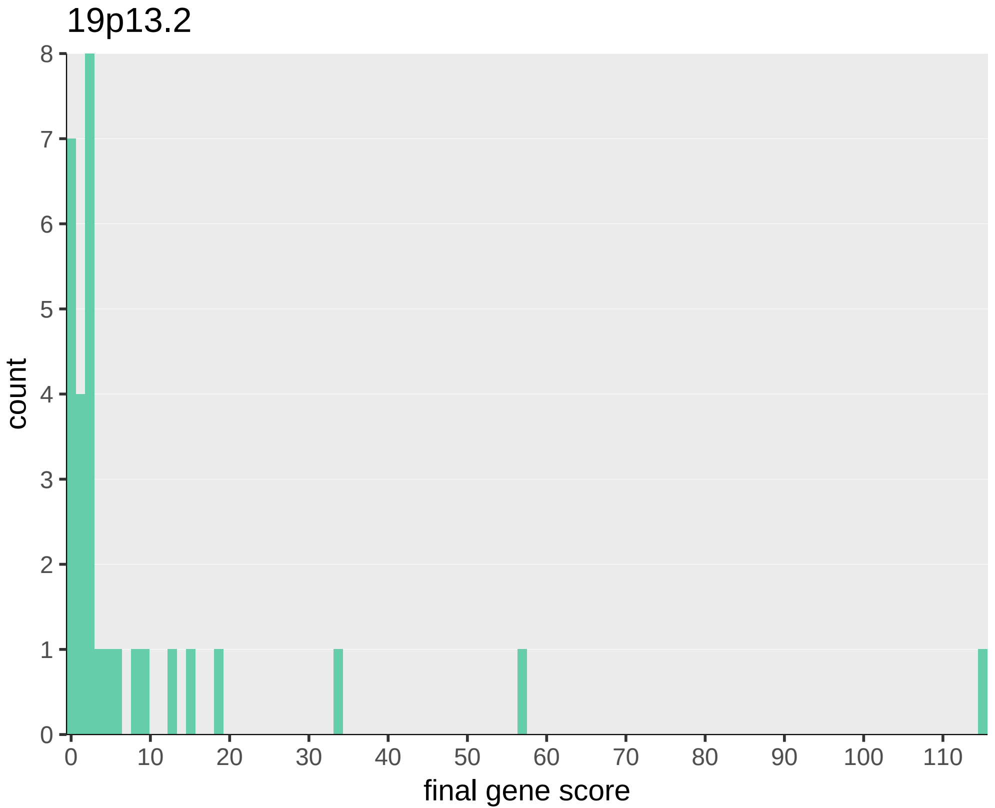
19p13.2
count
final gene score
0
1
2
3
4
5
6
7
8
0
10
20
30
40
50
60
70
80
90
100
110

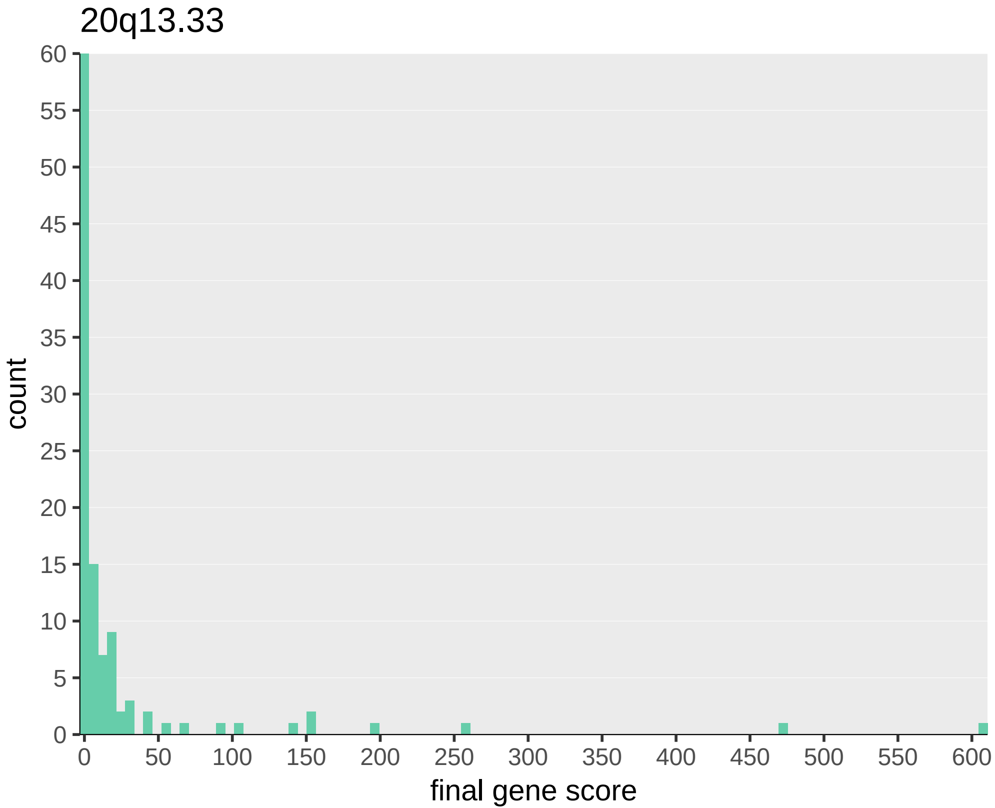
20q13.33
count
60
55
50
45
40
35
30
25
20
15
10
5
0
0
50
100
150
200
250
300
350
400
450
500
550
600
final gene score
