## Additional File 4 Supplementary Figure 3 for "Triangulating molecular evidence to prioritise candidate causal genes at established atopic dermatitis loci"

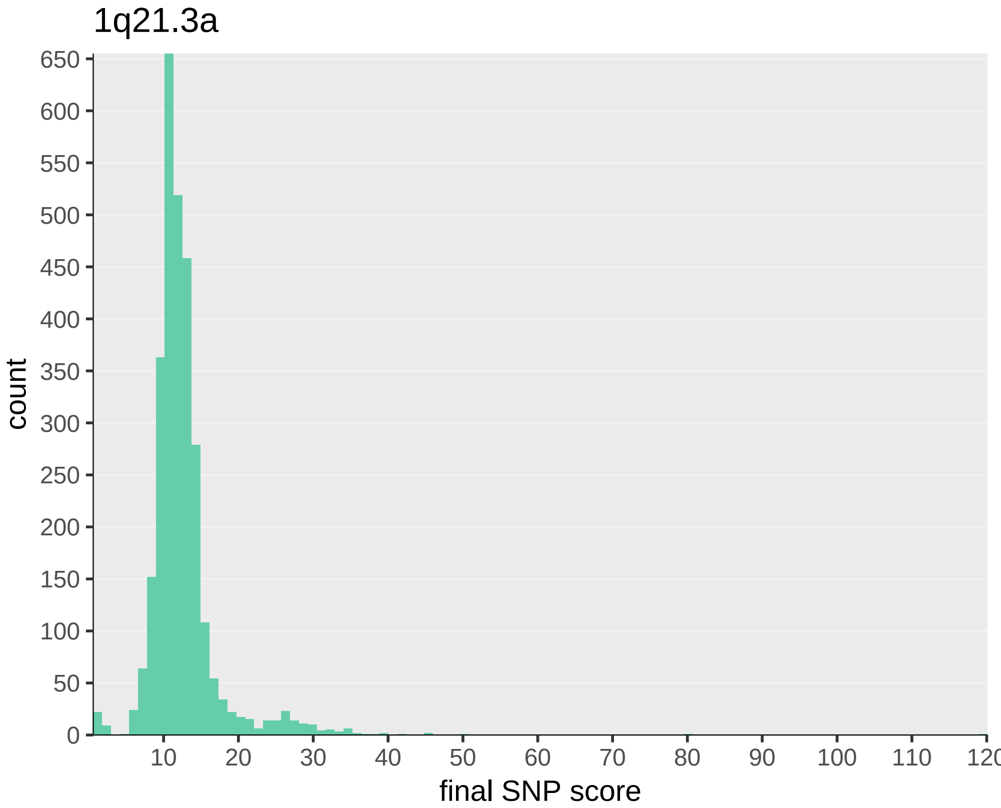

1q21.3a
count
650
600
550
500
450
400
350
300
250
200
150
100
50
0
10
20
30
40
50
60
70
80
90
100
110
120
final SNP score

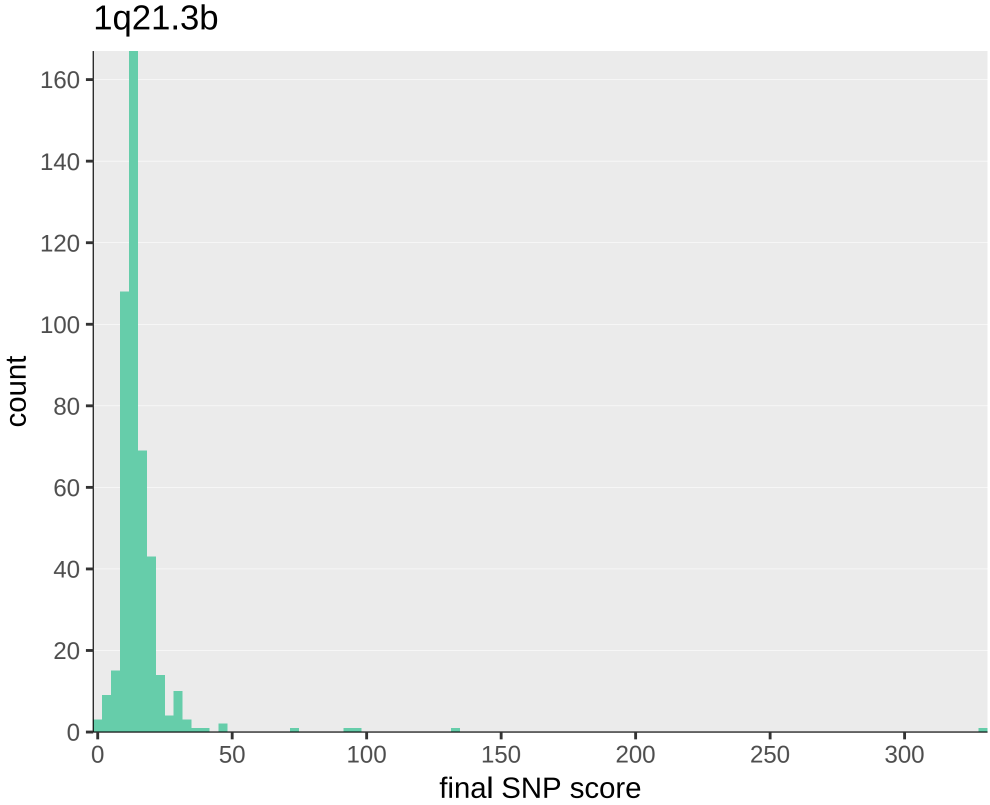
1q21.3b
count
160
140
120
100
80
60
40
20
0
0
50
100
150
200
250
300
final SNP score

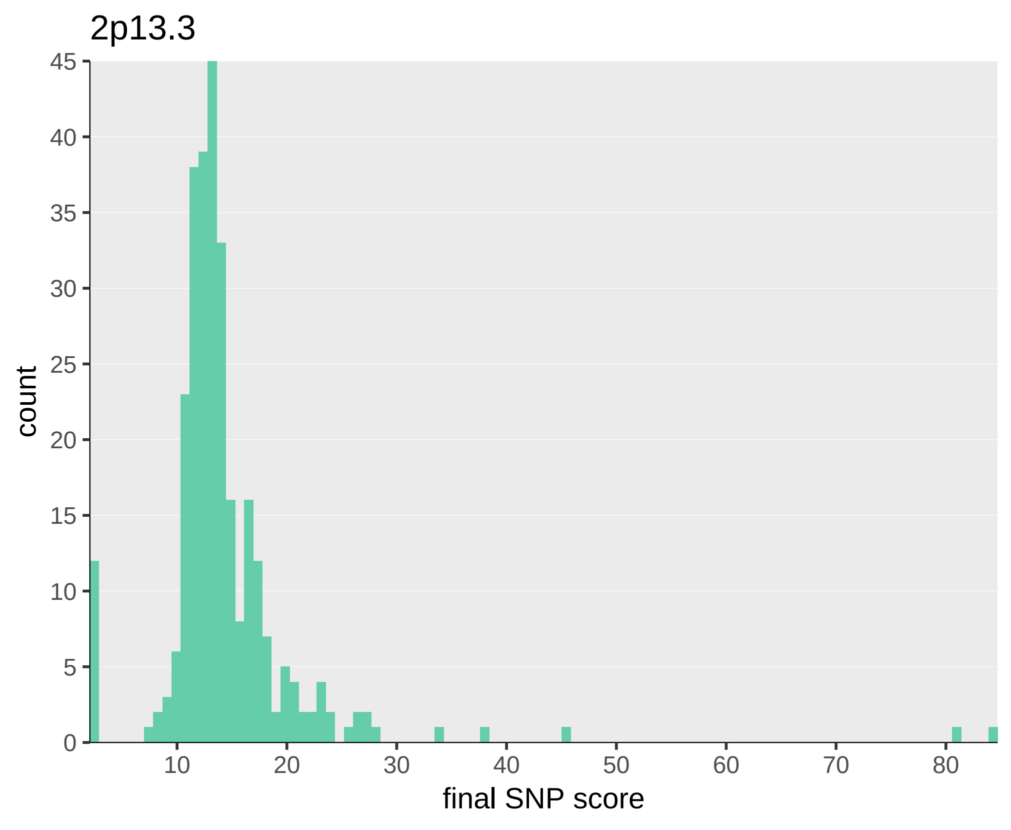

2p13.3
count
final SNP score
0
5
10
15
20
25
30
35
40
45
10
20
30
40
50
60
70
80

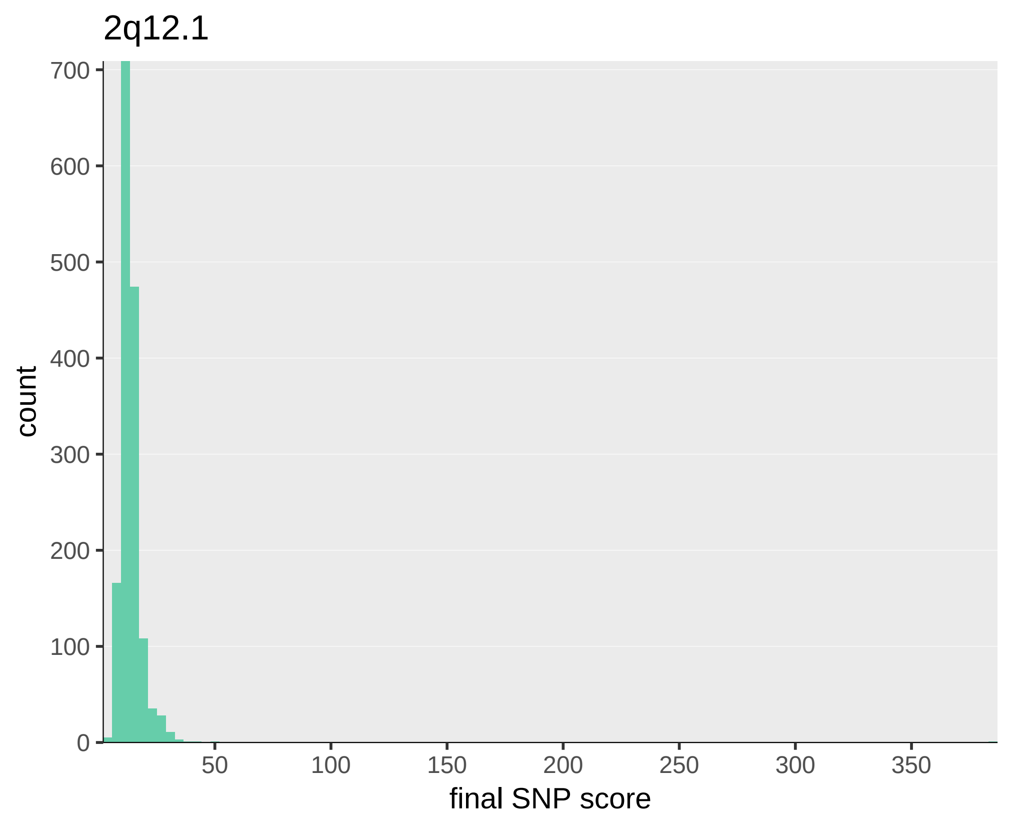
2q12.1
700
600
500
400
300
200
100
0
count
50
100
150
200
250
300
350
final SNP score

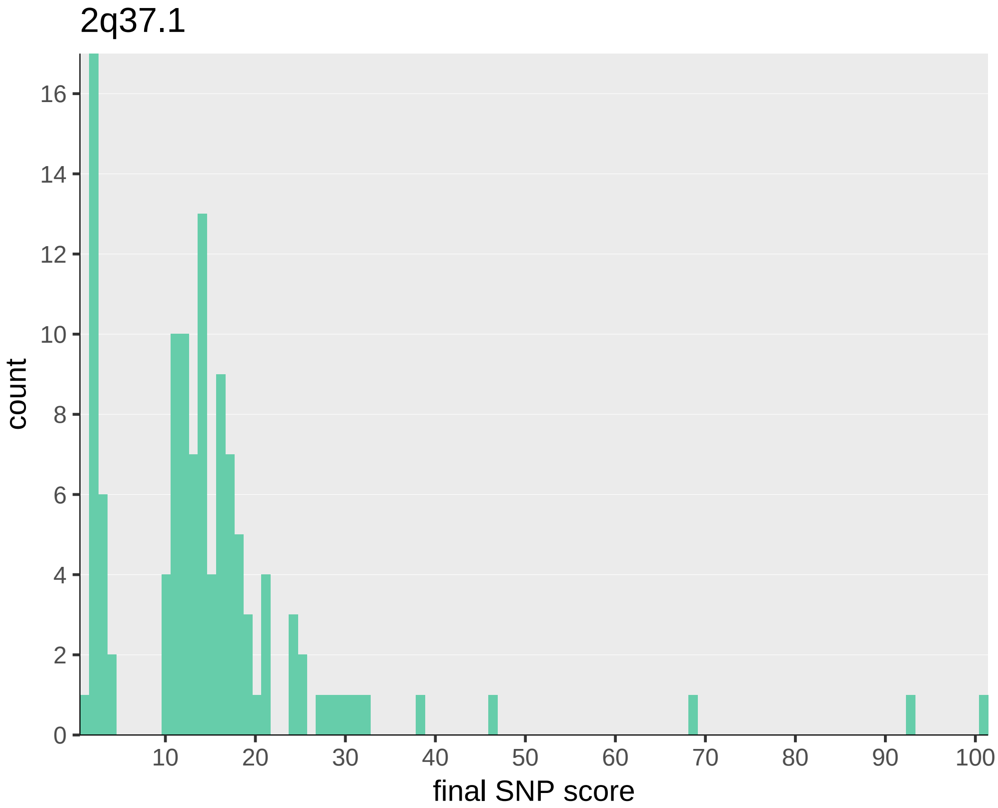

2q37.1
count
0
2
4
6
8
10
12
14
16
10
20
30
40
50
60
70
80
90
100
final SNP score

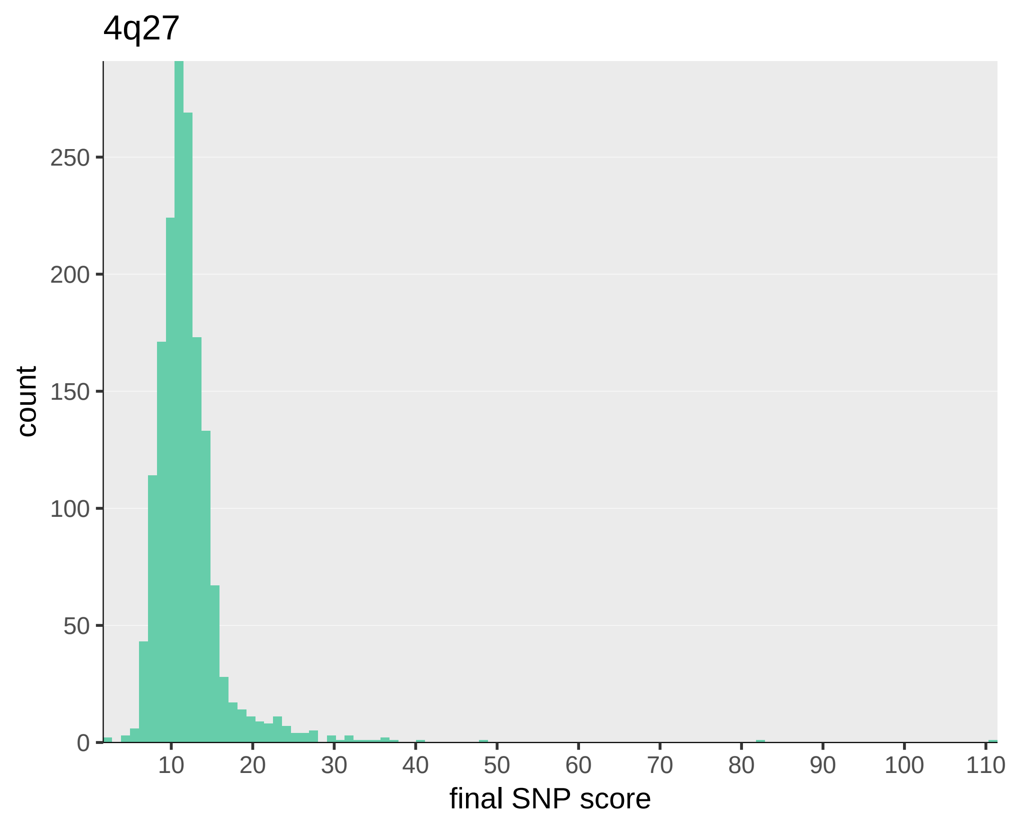
4q27
count
250
200
150
100
50
0
10
20
30
40
50
60
70
80
90
100
110
final SNP score

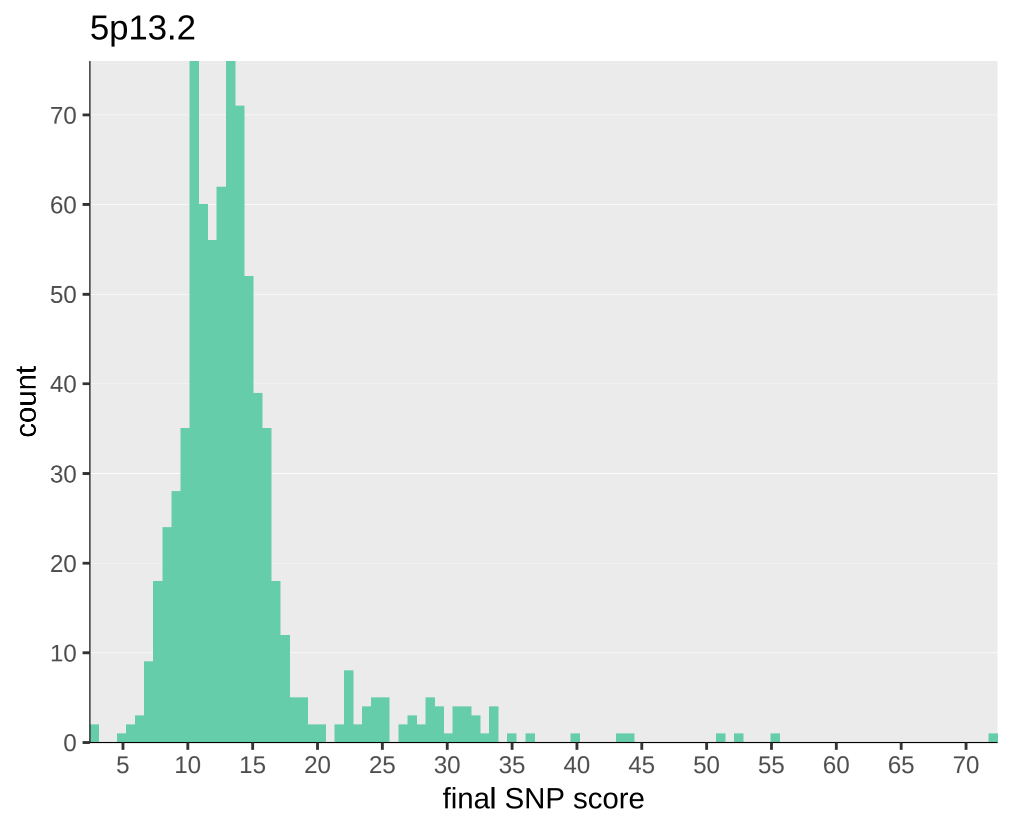
5p13.2
count
0
10
20
30
40
50
60
70
5
10
15
20
25
30
35
40
45
50
55
60
65
70
final SNP score

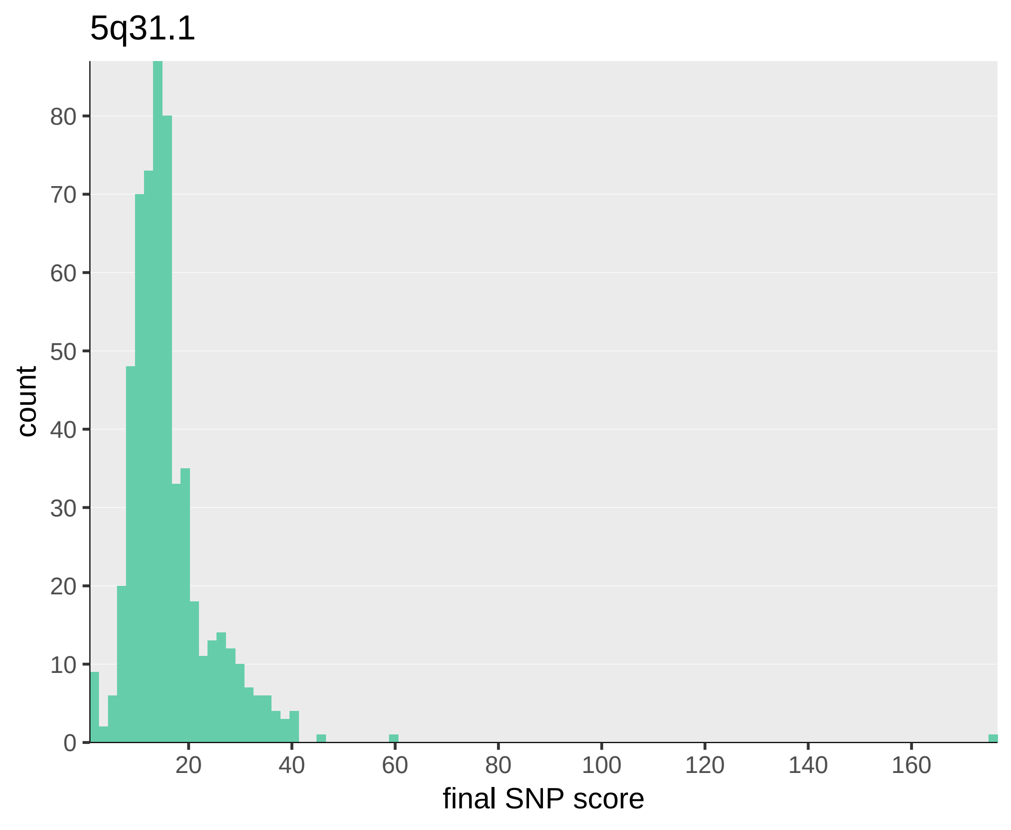
5q31.1
count
80
70
60
50
40
30
20
10
0
20
40
60
80
100
120
140
160
final SNP score

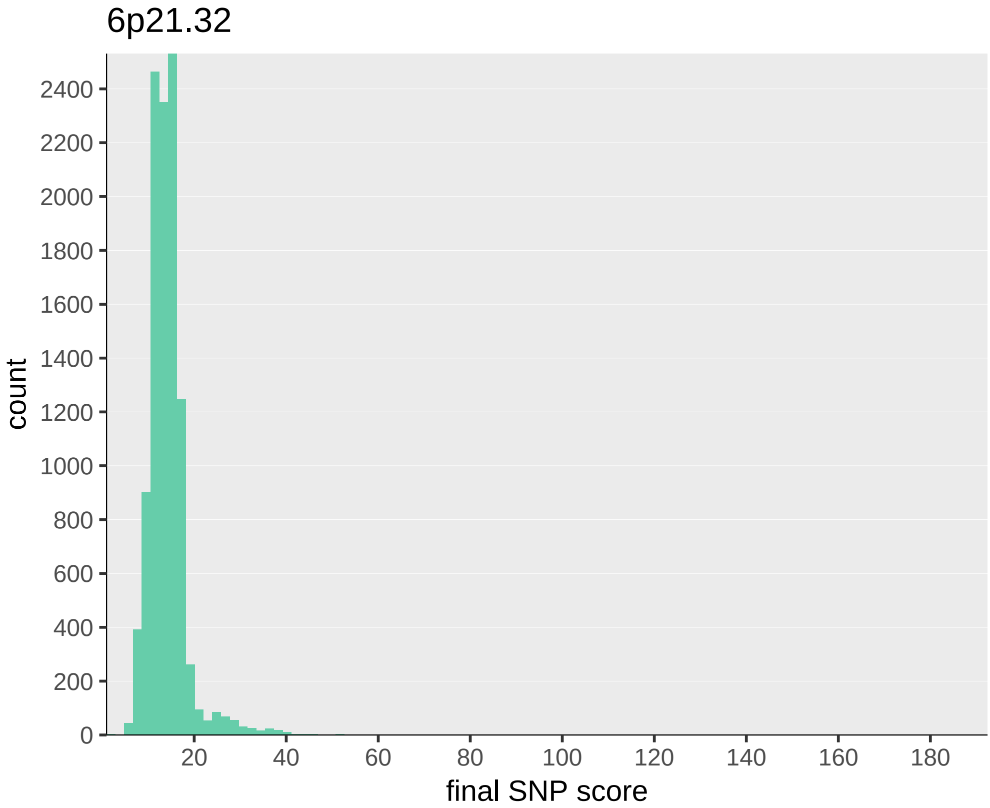
6p21.32
count
2400
2200
2000
1800
1600
1400
1200
1000
800
600
400
200
0
20
40
60
80
100
120
140
160
180
final SNP score

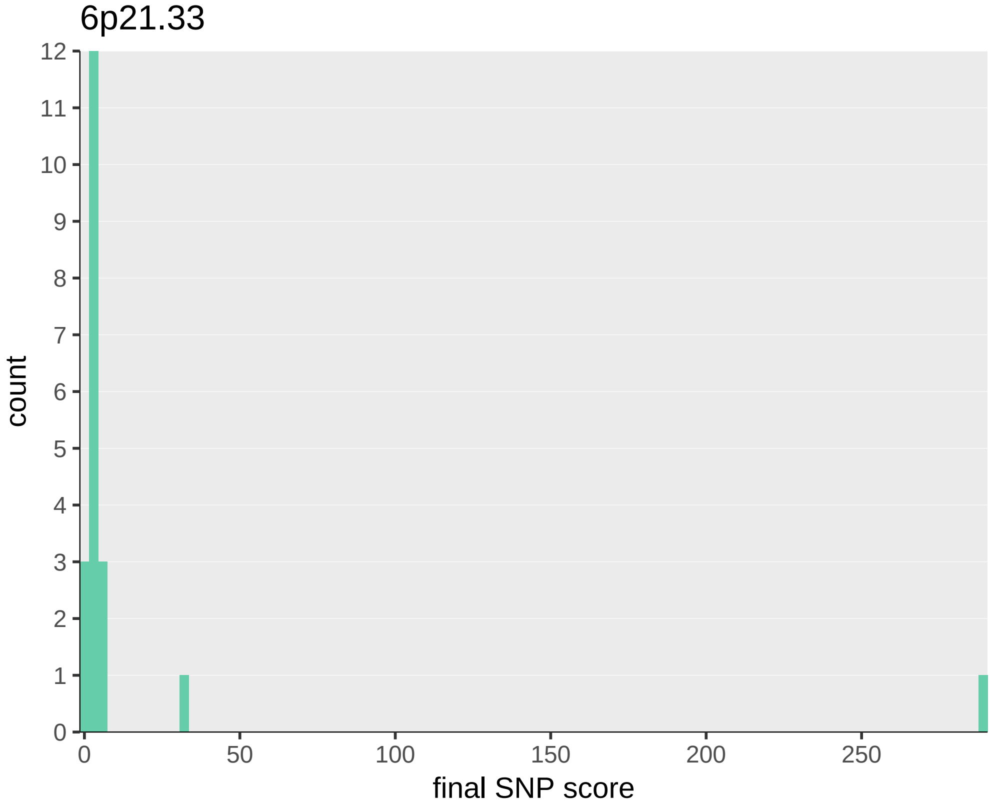

6p21.33
count
final SNP score
0
1
2
3
4
5
6
7
8
9
10
11
12
0
50
100
150
200
250

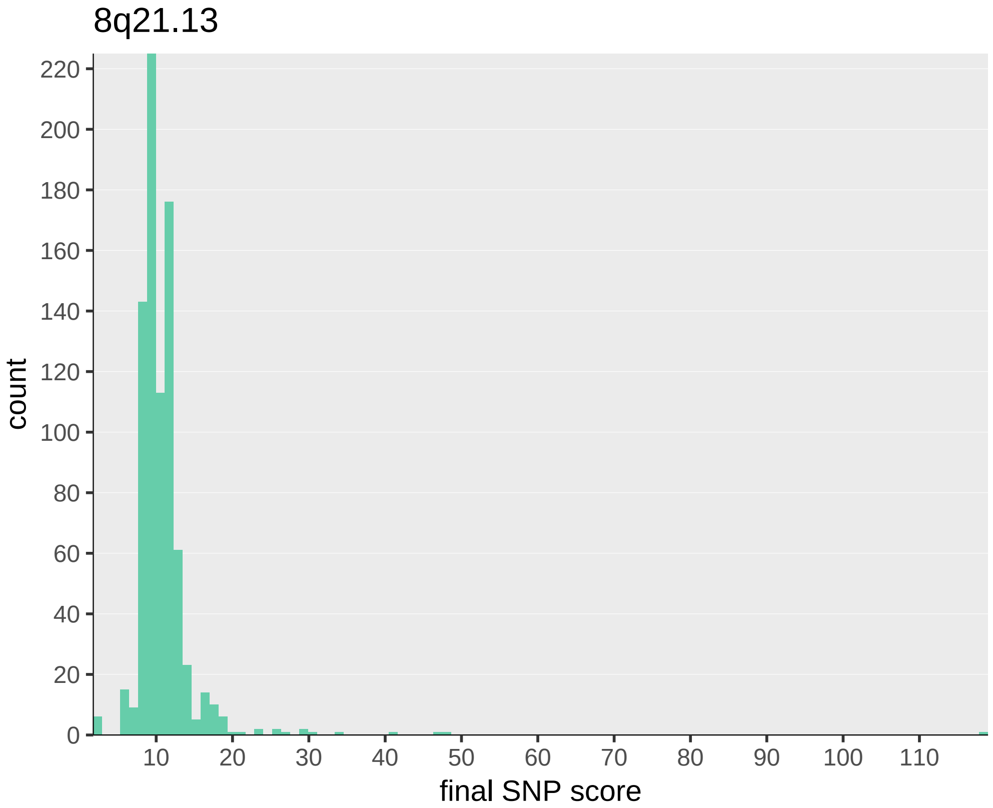

8q21.13
count
220
200
180
160
140
120
100
80
60
40
20
0
10
20
30
40
50
60
70
80
90
100
110
final SNP score

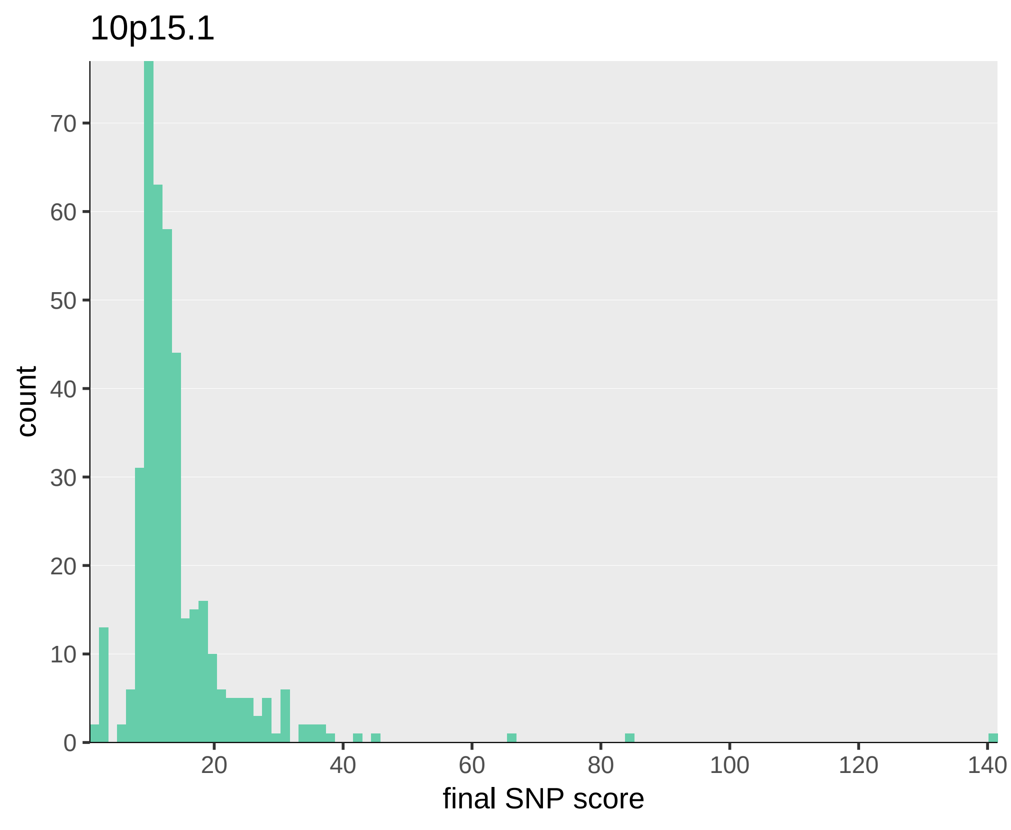

10p15.1
count
0
10
20
30
40
50
60
70
20
40
60
80
100
120
140
final SNP score

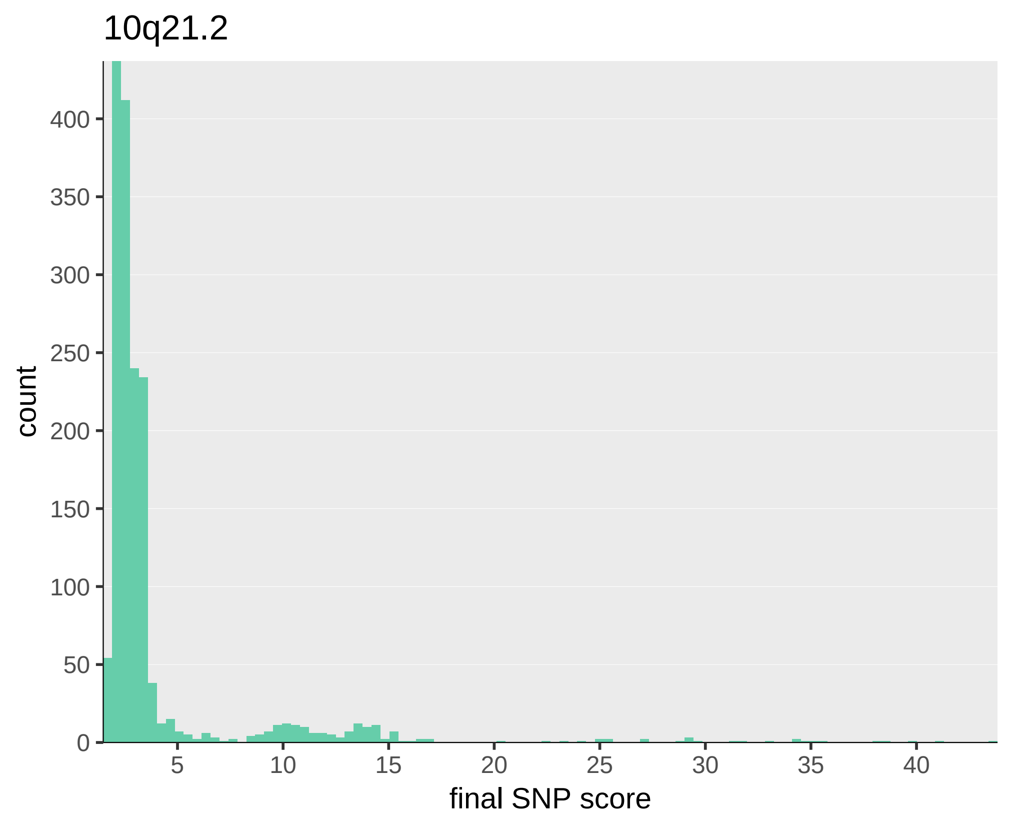
10q21.2
count
400
350
300
250
200
150
100
50
0
5
10
15
20
25
30
35
40
final SNP score

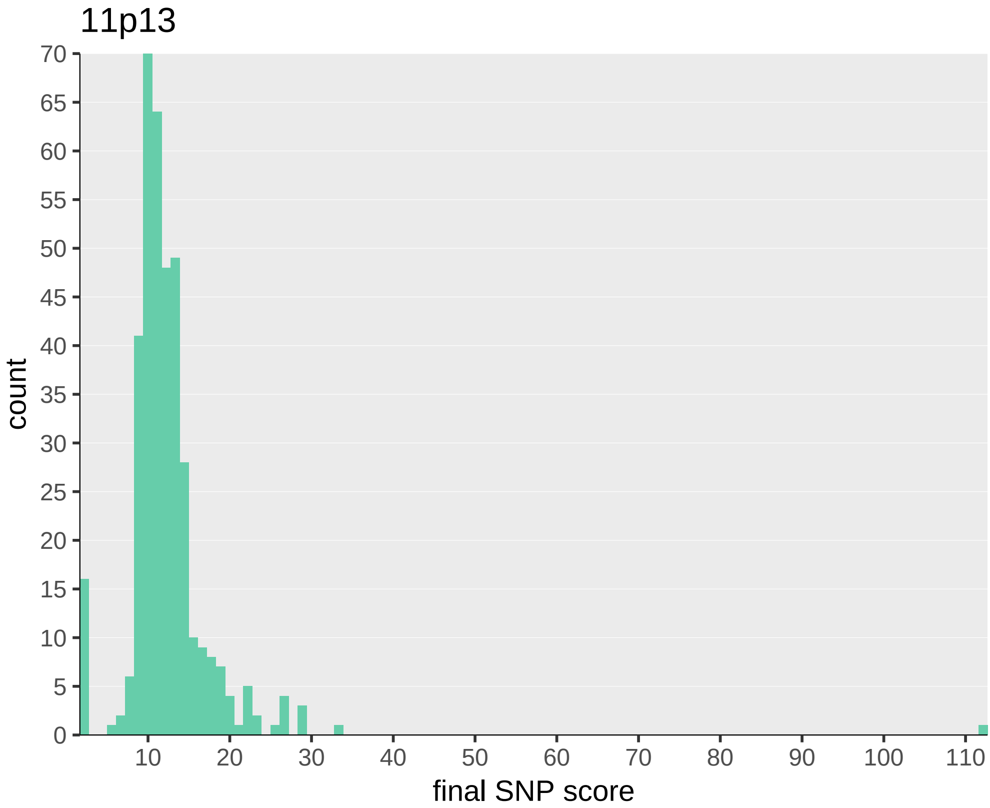

11p13
count
70
65
60
55
50
45
40
35
30
25
20
15
10
5
0
10
20
30
40
50
60
70
80
90
100
110
final SNP score

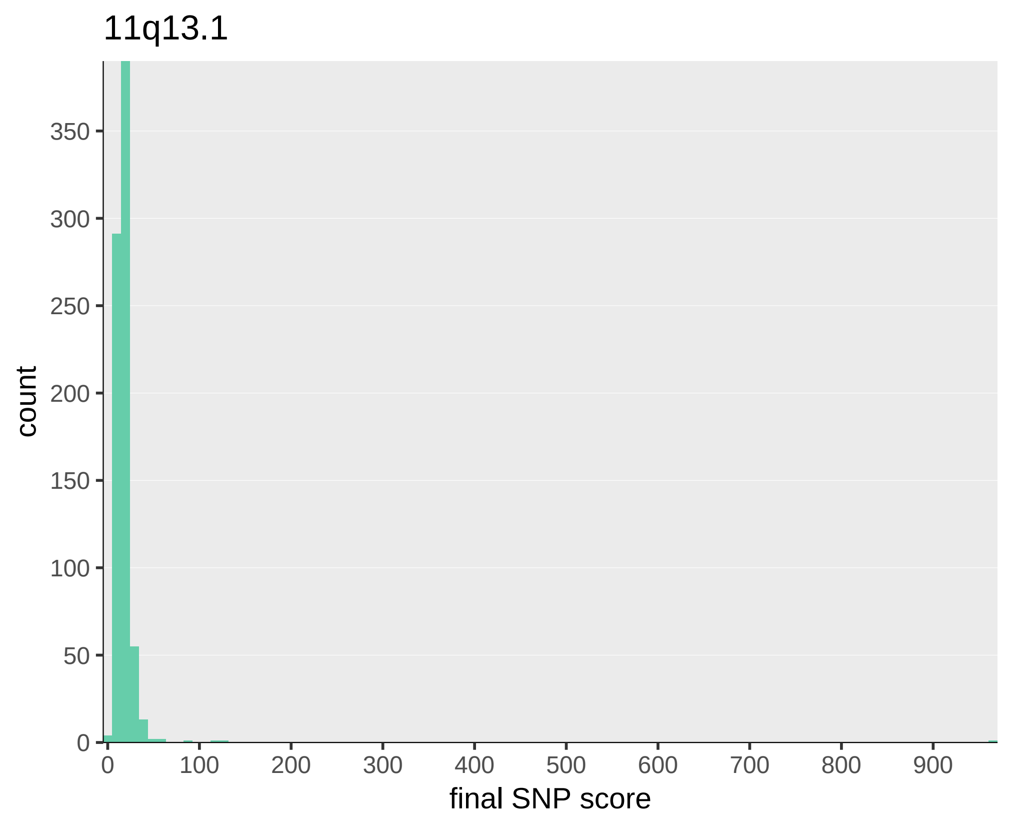
11q13.1
count
350
300
250
200
150
100
50
0
0
100
200
300
400
500
600
700
800
900
final SNP score

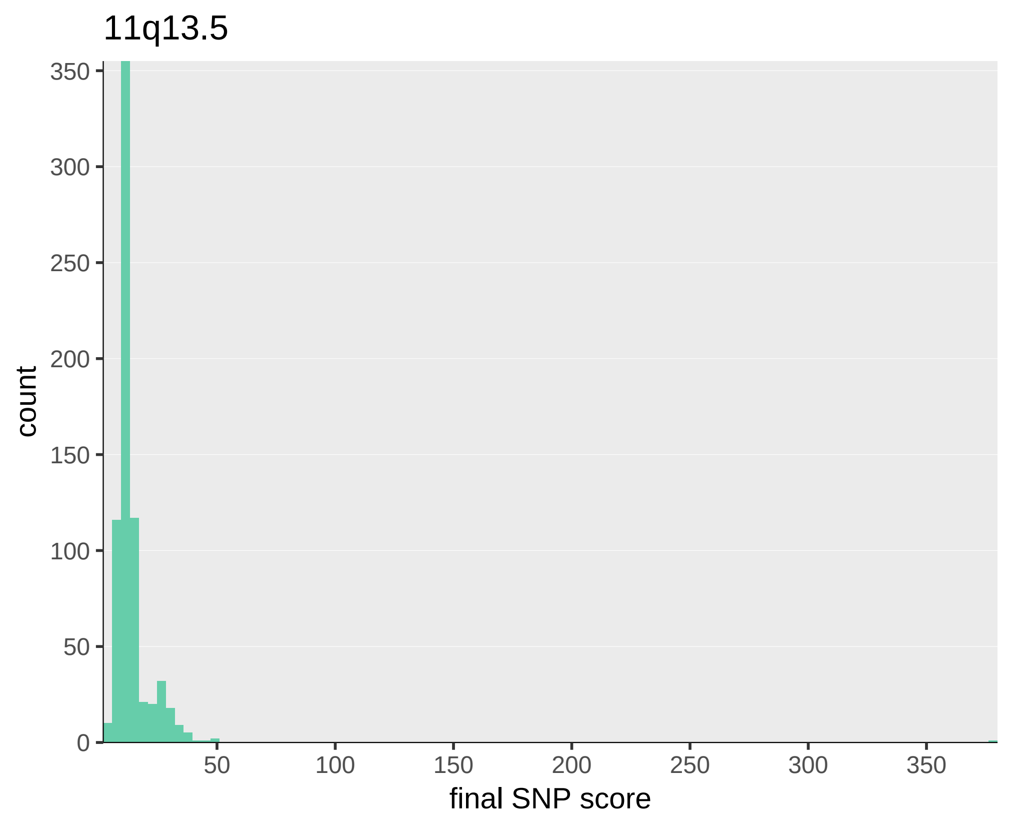
11q13.5
count
350
300
250
200
150
100
50
0
50
100
150
200
250
300
350
final SNP score

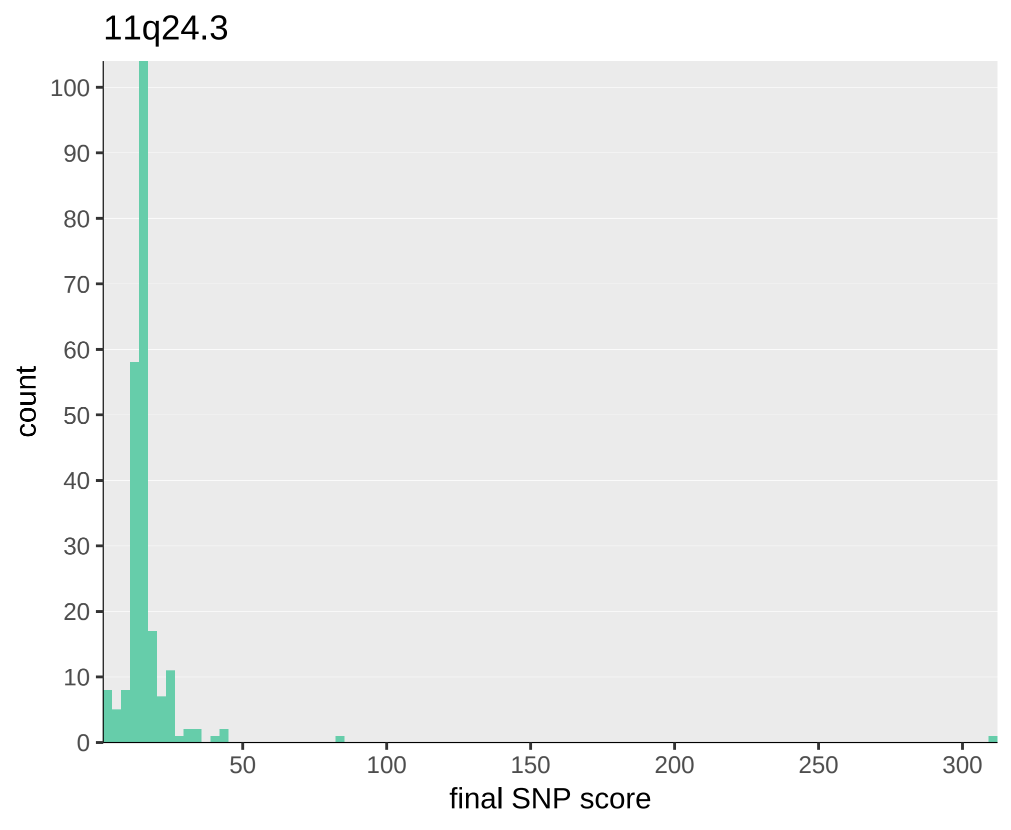

11q24.3
count
100
90
80
70
60
50
40
30
20
10
0
50
100
150
200
250
300
final SNP score

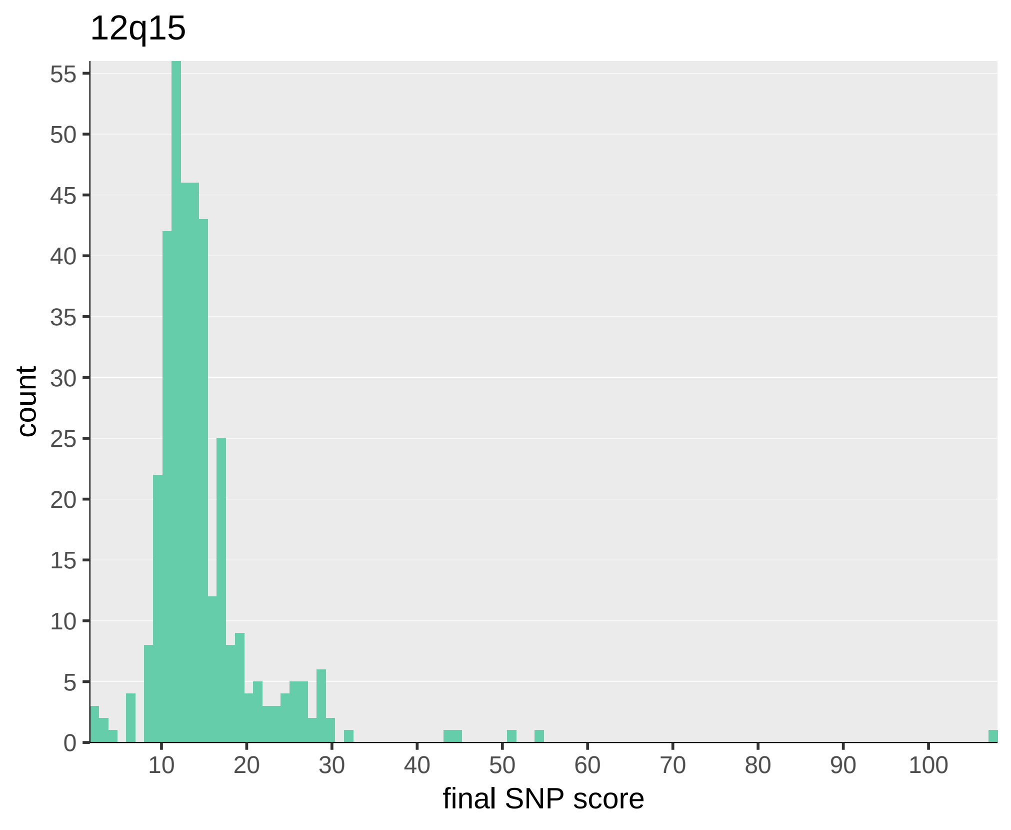

12q15
count
0
5
10
15
20
25
30
35
40
45
50
55
10
20
30
40
50
60
70
80
90
100
final SNP score

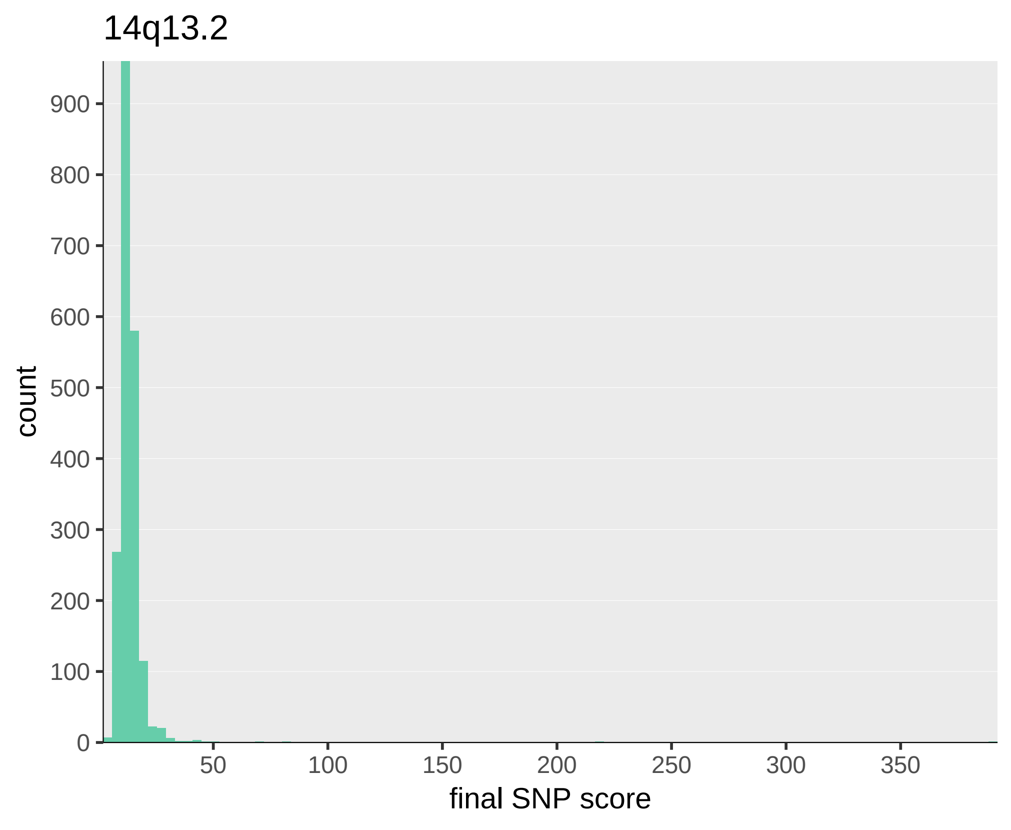

14q13.2
count
900
800
700
600
500
400
300
200
100
0
50
100
150
200
250
300
350
final SNP score

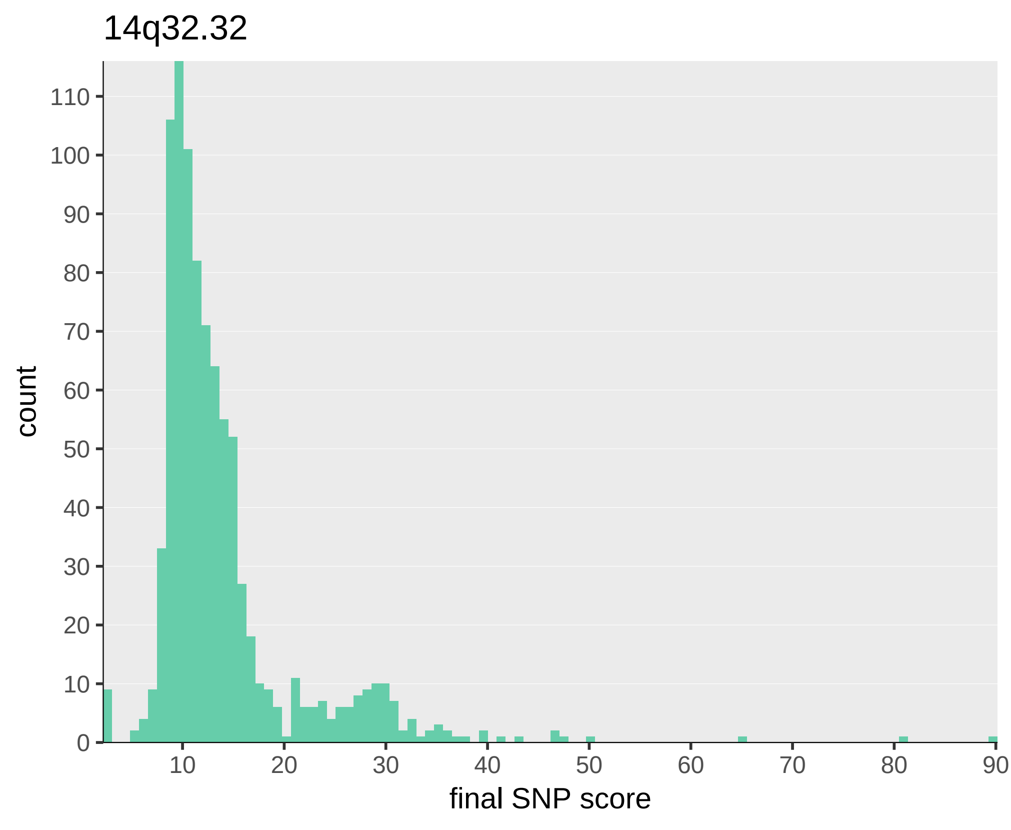
14q32.32
count
final SNP score
0
10
20
30
40
50
60
70
80
90
100
110
10
20
30
40
50
60
70
80
90

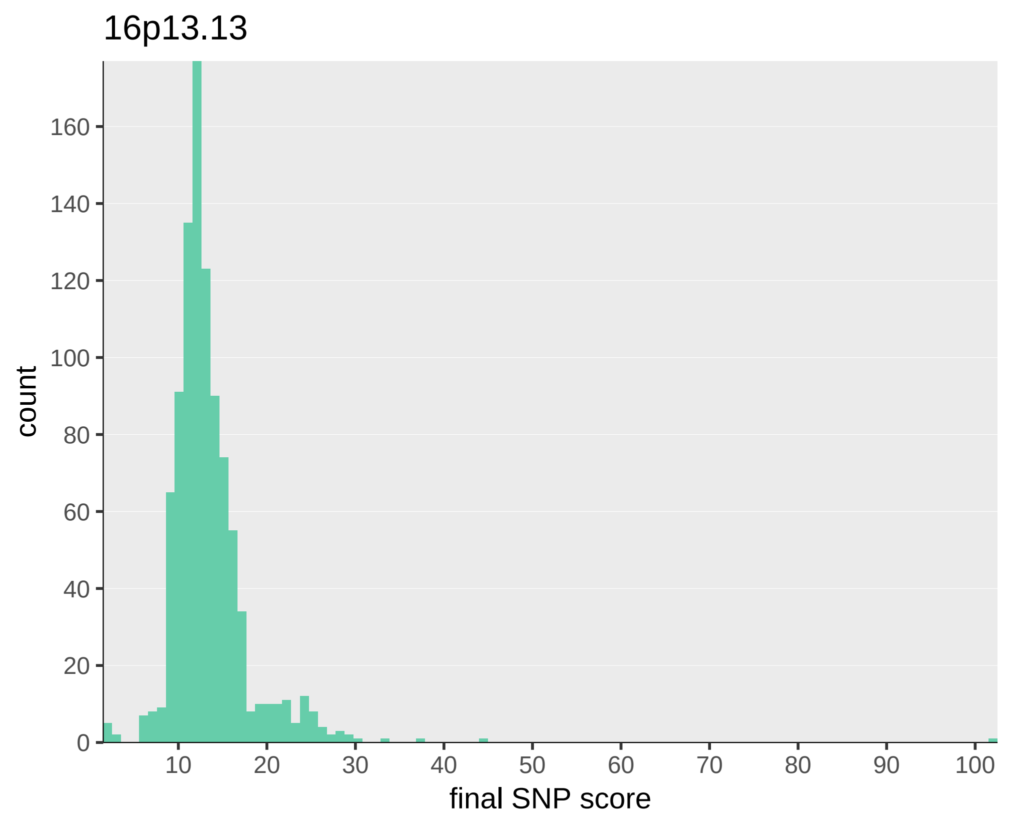
16p13.13
count
160
140
120
100
80
60
40
20
0
10
20
30
40
50
60
70
80
90
100
final SNP score

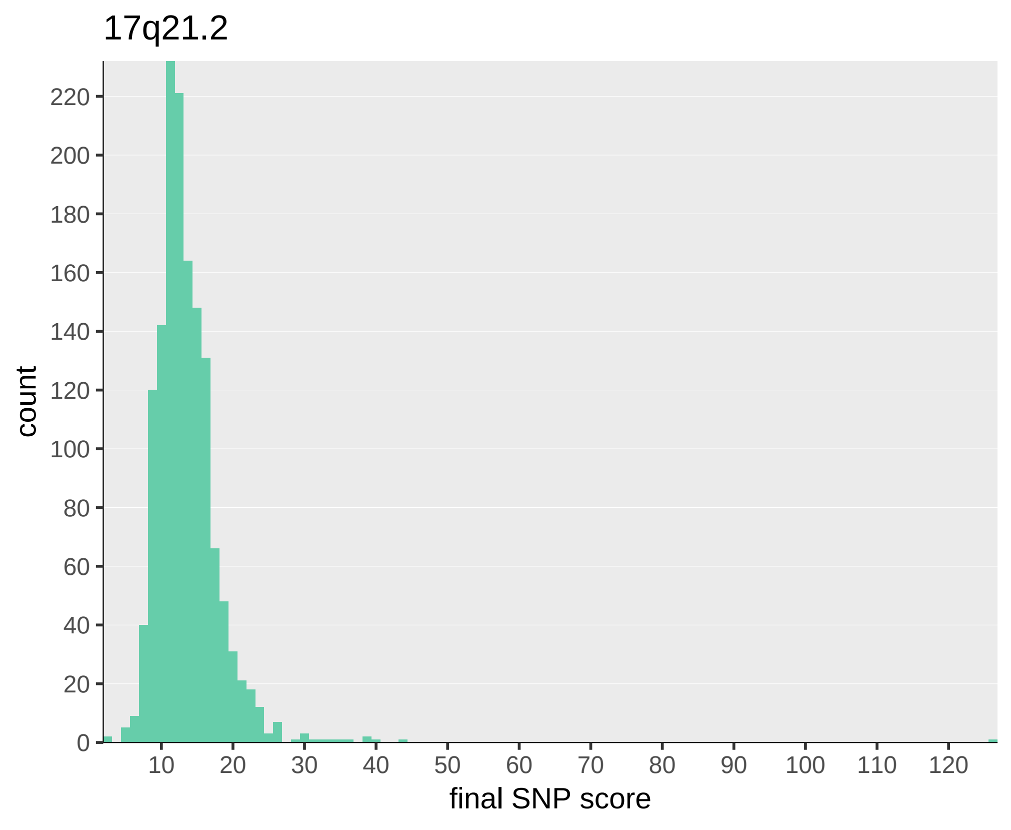
17q21.2
count
220
200
180
160
140
120
100
80
60
40
20
0
10
20
30
40
50
60
70
80
90
100
110
120
final SNP score

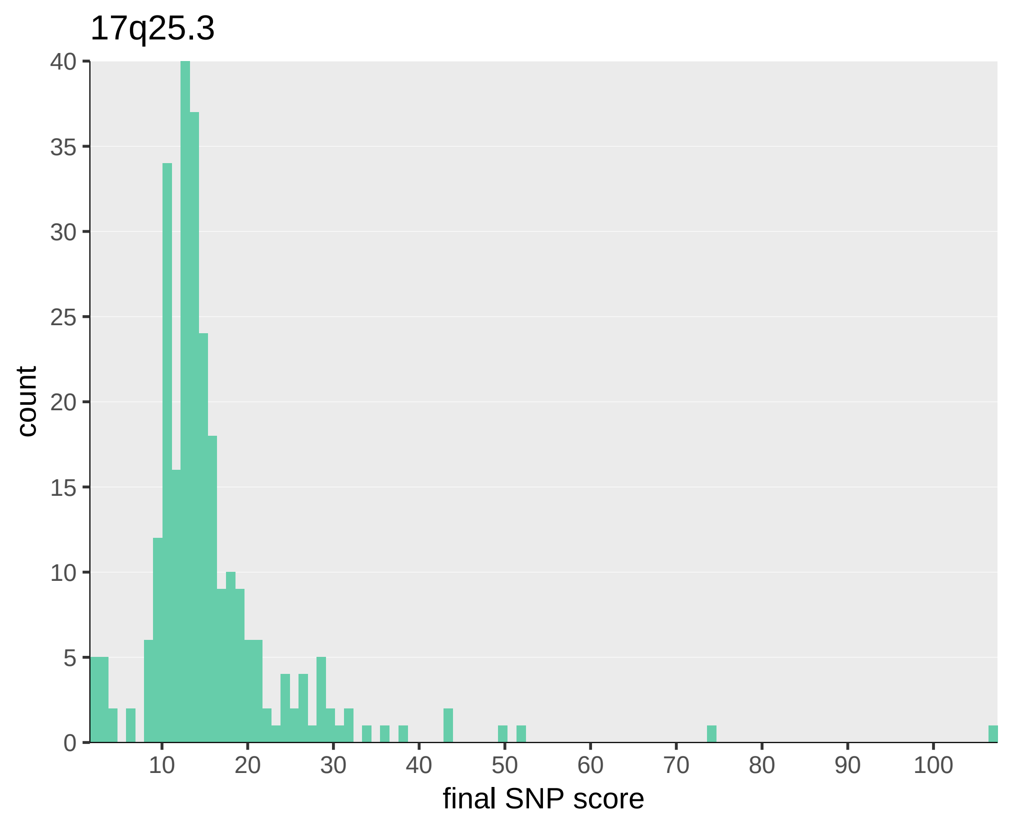

17q25.3
count
final SNP score
0
5
10
15
20
25
30
35
40
10
20
30
40
50
60
70
80
90
100

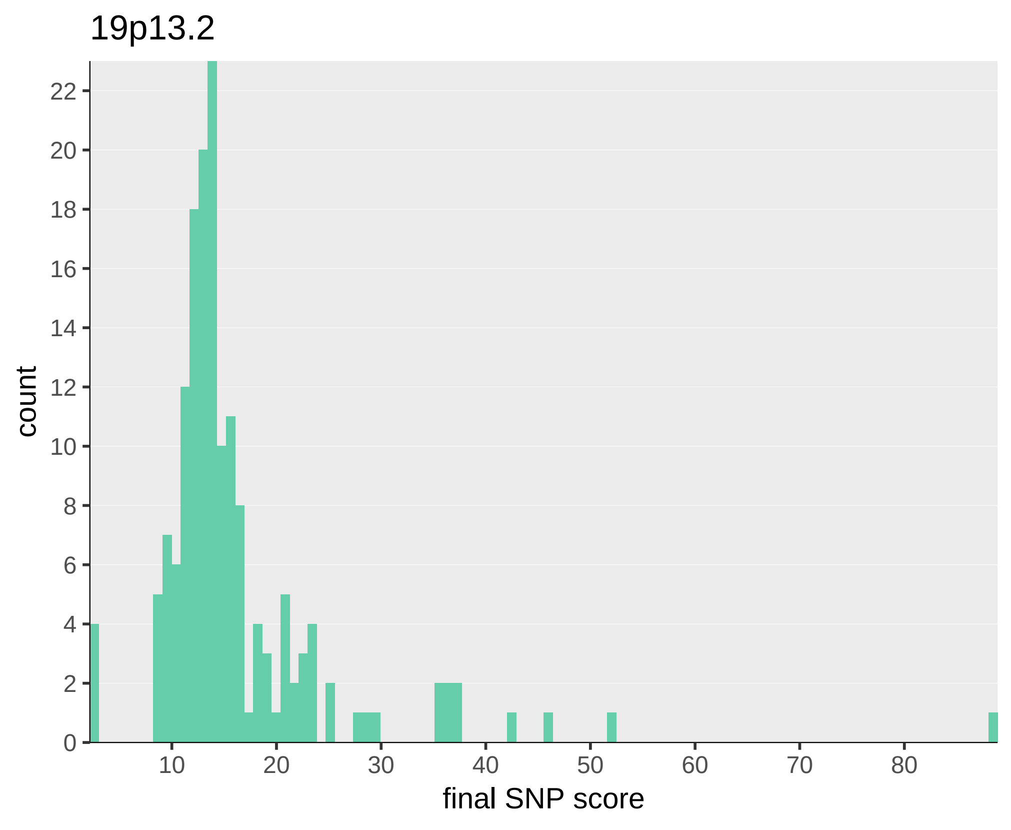
19p13.2
count
22
20
18
16
14
12
10
8
6
4
2
0
10
20
30
40
50
60
70
80
final SNP score

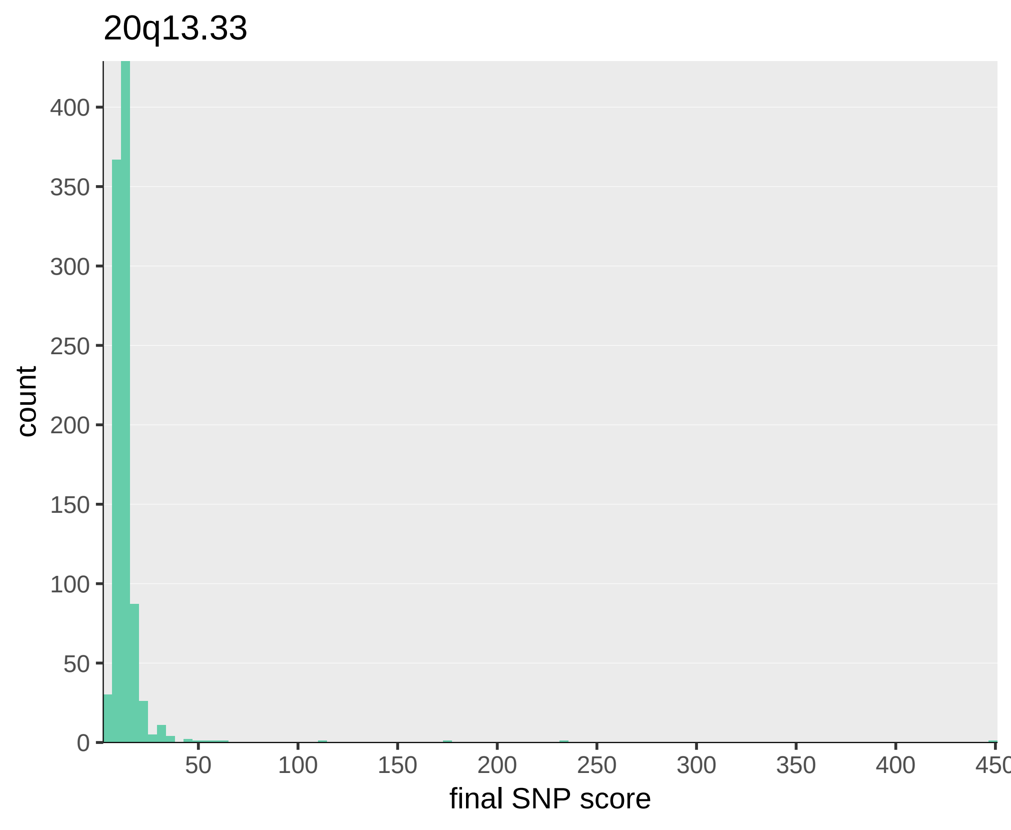

20q13.33
count
final SNP score
0
50
100
150
200
250
300
350
400
50
100
150
200
250
300
350
400
450
