## Additional File 7 Supplementary Text 2 for "Triangulating molecular evidence to prioritise candidate causal genes at established atopic dermatitis loci"

**Supplementary Text 2. ADGAPP prioritisation summary for AD loci.**

**Locus 1q21.3**

At least two signals are known to be present in this epidermal differentiation complex region, rich in ~45 genes with roles in late cornified envelope (LCE) formation[1]. One signal, represented by GWAS index SNP rs61812715 (or rs12144049 in Schaarschmidt *et al.* (2015)[2]), likely represents the known filaggrin (*FLG*) mutations, as the signal disappears after conditioning on the four most common substitutions[3]. We hereby refer to this signal as locus 1q21.3 - a. The second independent signal is represented by rs12730935[3]. We refer to this signal as locus 1q21.3 - b and feature it also in the Results section in the main body of the paper.

*Locus 1q21.3 - a*

We considered 151 genes within 3 Mbp of index SNP rs61812715 in 1q21.3 - a to be potential candidate genes (Dataset S4). Whilst the causal gene at this locus has been shown to be FLG[3], our analysis was fairly inconclusive (with several genes with similar moderate scores) and the results gave three alternative filaggrin-like genes as the top prioritised loci. Top was hornerin (*HRNR*, score 464), which accounted for 28% of the cumulative score of the top 10 genes (Table 1), followed by repetin (*RPTN*, score=285) and cornulin (*CRNN*, score=249), which respectively account for 17% and 15% of the top 10 score. *FLG* was ranked 6 (score=108). Our gene colocalisation analysis (Additional File 5: Supplementary Table 1) was not conclusive. It showed some evidence for *FLG* (AD effect signal colocalised with reduced expression in skin, TWAS *p* = 1x10^-11^), but also for other genes, *CRNN*, *FLG-AS1*, *IVL*, *LCE1E*, *SPRR1B* and *SPRR2D*. The strongest evidence being for CRNN - a filaggrin-like protein (effect signal colocalised with increased expression, TWAS *p* = 1x10*^-20^*). Our inconclusive findings in this region are likely due to the complex and highly duplicate nature of the differentiation complex, with many homologous genes, probably under coordinated regulation. Although conditional analysis in the EAGLE GWAS suggested that this signal is explained by the known functional FLG mutations[3], a previous study has suggested that there may be an independent signal in markers surrounding the HRNR gene, and so this should be considered in future work on this locus[4].

*Locus 1q21.3 - b*

At this locus we tested 104 genes within the 3 Mb interval (Figure 3 & Dataset S4). Interleukin 6 receptor (*IL6R)* had the highest prioritisation score – 743 (62% of top 10 cumulative score; Table 1 & Figure 3A), with the next best gene being *UBE2Q1*- score of 93 (only additional 8% of top 10 cumulative score). The high prioritisation of *IL6R* by ADGAPP is mostly driven by colocalisation (Figure 2) in whole blood in eQTLGen and GTex (coloc posterior probability of colocalisation (PPH4) = 63%-88% and TWAS *p*-value = 5 x 10^-6^), in lipopolysaccharide-/muramyl dipeptide- primed monocytes in Kim-Hellmuth *et al.* (PPH4 = 75%-77%) and sun-unexposed skin in GTEx (PPH4 = 64%) (Additional File 5: Supplementary Table 1). Possible placement of many locus interval SNPs within an enhancer of the gene is highlighted by HiC interaction data in human embryonic stem cells[79], whole blood[81], CD34+ hematopoietic cells and lymphoblastoid cell lines[146] and epidermal stem cells along with keratinocytes[5]. Previously, *IL6R* was also implicated as the potential causal gene at this locus in the EAGLE GWAS[3] as one of the variants in the MANTRA-based credible set of 23 associated variants, rs2228145, represents a missense (Asp358Ala) mutation in the gene[6]. This variant is ranked 3^rd^ in ADGAPP and is associated with blood serum levels of IL6R protein as well as transcript[7]. However, the actual action of rs2228145 is to shift IL6R protein from membrane-bound state to soluble cell fraction, or in other words, from the anti-inflammatory classical signalling pathway to pro-inflammatory trans-signalling[8]. Mendelian randomization analyses indicated increased soluble IL6R levels as causal for higher AD (and asthma) risk and decreased soluble IL6R levels indicative of increased rheumatoid arthritis (RA) risk[9]. This is in agreement with the Asp358Ala mutation being protective towards RA and increasing risk in AD[10]. However, in our colocalisation analyses the AD risk allele is linked to overall reduction in *IL6R* transcript levels, likely due to the complex balance of IL6 classical and trans-signalling. A range of anti-IL6 biologics are in clinical use and development for inflammatory diseases, including tocilizumab, which inhibits both soluble and membrane-bound IL6R, and is approved for treatment of RA and juvenile idiopathic arthritis[11]. However, AD adverse events have been reported for in tocilizumab trials[12], consistent with the opposing effects of this GWAS locus on RA and AD.

**Locus 2p13.3**

Altogether, 35 potential candidate genes were tested at this locus within 3 Mbp centred around index SNP rs112111458 (Dataset S4). Among the top 3 ranked genes, *CD207* clearly stood out with the score of 272 (45% of the top 10 total, Table 1) compared to 2^nd^ and 3^rd^ ranked *CLEC4F* and *VAX2* (scores of 62 (10%) and 56 (9%), respectively). Our analysis showed strong colocalisation with the protective allele leading to reduction of gene expression in the skin in TwinsUK RNA-Seq data, Sun-exposed and unexposed skin in GTEx, as well as transformed fibroblasts according to TWAS analysis (Additional File 5: Supplementary Table 1). No colocalisation was detected for the other genes in the locus, which makes *CD207* a particularly promising candidate.

The original GWAS analysis in Paternoster et al. (2015) proposed that *CD207* is the likely candidate gene behind the locus due to colocalisation of its microarray-derived eQTLs in TwinsUK (MuTHER) skin samples, CD207’s specific expression in the skin Langerhans cells and the cells’ abnormal phenotype presenting in the mouse knockout model. *CD207’s* protein product langerin marks Langerhans cells resident in the stratified squamous epithelium of the skin and mucosal tissues[13]

**Locus 2q12.1**

This locus consists of two independent signals represented by rs6419573 and rs3917265 lead SNPs[3]. Altogether, we considered 46 genes located within the 3 Mbp interval of each SNP (Figure 3B & Dataset S4). As mentioned earlier, the highest scores were obtained by *Interleukin-18 receptor 1* - *IL18R1* (score=1384, Table 1) and *Interleukin-18 receptor accessory protein 1* - *IL18RAP* (score=1341) which together account for 77% of the top 10 cumulative score at the locus, and so are likely to represent the causal genes behind the two signals. *IL18R1* and *IL18RAP*’s roles are supported by abundant expression colocalisation evidence in the whole blood in GTEx (*IL18R1*: PPH4 = 51%-80% and p-value = 7 x 10^-6^; *IL18RAP*: PPH4 = 96%-99% and p-value = 4 x 10^-12^) and immune cell types in CEDAR, TwinsUK and Kim-Hellmuth *et al.* (*IL18R1*: PPH4 = 55%-86% and p-value = 2 x 10^-5^, *IL18RAP*: PPH4 = 55%-97% and p-values = 5 x 10^-5^ to 6 x 10^-8^), as well as skin in TwinsUK for *IL18R1* only (p-value = 1 x 10^-4^*;* Figure 2 & Additional File 5: Supplementary Table 1). The direction of effect indicates increased expression of both genes for AD risk alleles, with one exception in GTEx whole blood for *IL18RAP* (Additional File 5: Supplementary Table 1). Interestingly, opposing expression effects of the same allele in a variant has previously been reported in monocytes and whole blood for *IL18RAP*[14]. SNPs in LD with the lead SNP at the three top-ranked genes overlapped whole blood mQTL SNPs in the GoDMC study[15], likely influencing the methylation of promoters of those genes. Differential gene expression (DGE) consistently shows significant upregulation of *IL18RAP*[16] and *IL18R1*[17,18] in the skin of AD patients (Dataset S5). Both genes were put forward as candidate genes in the EAGLE GWAS annotation, as the credible set variants span both genes and individual variants linked to *IL18R1* eQTLs in TwinsUK skin microarray data, but previously there was little evidence for colocalisation.

In general, cytokine IL-18, in the formation of whose receptor complex both IL18RAP and IL18R1 participate, has been shown to elicit significant immune gene expression change in keratinocytes from AD lesions[18]. *IL18R1* and *IL18RAP* are both involved in T-cell, especially T helper cell signalling and activation of the NFkB-pathway. IL18R1 and IL18RAP mediate IL-18-dependent signal transduction, which is of special importance in Th1 response. Thus, it would not be surprising if one variant affected the expression of the whole gene cluster, and that has indeed been shown to be the case with SNP rs917997 associated with IBD and coeliac disease[19]; and ranked 8^th^ among the two signals. A variant situated 15 kbp away from rs917997 - rs990171, again associated with celiac disease and lymphocyte counts[20], is ranked as second-best among the two signals in the locus (Dataset S4). There is strong pQTL support for association of rs990171 with IL18R and IL1RL1 resulting in increased protein availability[21] (Dataset S5). In addition, rs990171 overlaps allele-specific eQTLs for IL18RAP and acetylation hQTLs in neutrophils[22] appearing during Th1 polarization[23].

**Locus 2q37.1**

This locus was only discovered in the original GWAS after MAGENTA gene set enrichment analysis and was later revealed as a shared locus for asthma, allergy and eczema[24]. Twenty candidate genes within 3 Mbp window of lead SNP rs1057258 were analysed in ADGAPP (Dataset S4). The pipeline strongly prioritised the proximal gene of inositol polyphosphate-5-phosphatase D (*INPP5D*) with the score of 296 (57% of total top 10 score, Table 1), with the second and third best genes - *ATG16L1*, *RN7SL32P* - scoring only 106 (20%) and 29 (6%) respectively.

Colocalisation evidence for *INPP5D* is tenuous, with posterior probability of shared causal GWAS and eQTL variant (PPH4) in monocytes in Blueprint estimated at 52% (Additional File 5: Supplementary Table 1) with the protective allele conferring reduced expression. The gene is upregulated in *FLG* mutation-harbouring eczema patients compared to negative controls[17], while downregulated in IBD, correlating there with reduced T CD4+ cell numbers[25]. PrixFixe network prioritisation algorithm selected *INPP5D* as the top candidate for the locus (Dataset S5). In addition, multiple exon-level eQTLs overlap SNPs at the locus, in agreement with existence of gene isoforms with internal promoters and displaying altered functionalities[26]. *INPP5D* was also proposed as the candidate gene at the locus in the original EAGLE GWAS[3] due to the location of the lead SNP, involvement of the gene in cytokine signalling and knockout mouse phenotype.

rs1057258, the lead and top ranked SNP at the locus (score=101, Dataset S4) was also identified as lead SNP in the joint atopy GWAS by Ferreira et al. (2017) and associated with Alzheimer’s disease progression[27]. The variant is located in the 3’ UTR region of the *INPP5D* gene and may contribute to its regulation. Another top *INPP5D* SNP with potential causal role is 2^nd^ ranked (score=93) intronic rs6740918 and 3^rd^ ranked (score=69) predicted missense SNP rs9247 (His>Asn/Tyr). rs9247 has been picked up as the top SNP for the locus in a new integrative GWAS method – FINDOR, which incorporates variant functional enrichment information into association detection[28]. The three variants: rs6740918, rs1057258 and rs9247 show histone modifications indicative of transcription in a variety of blood cell types[29] and are strong *INPP5D* eQTLs in naïve and induced monocytes[30,31], and additionally neutrophils for the two last SNPs[22] (Dataset S5).

INPP5D, also known as SHIP-1, is a general inhibitor of activation of the signalling cascades leading to immune cell survival and division, including lymphocyte activation[32,33]. The gene's signalling is also implicated in regulating neutrophil chemotaxis, macrophage activation, CD16-induced cytotoxicity in NK cells, among others.

**Locus 4q27**

The locus contains two independent signals, but we did not detect any differences in top candidate genes. Out of 132 genes contained within the 3 Mbp interval around index SNPs (Dataset S4), in each case we prioritise *KIAA1109* (score=220, 35% top 10 score), followed by *BBS12* (score=112), *TRPC3* (score=100), contributing 18% and 16% of the top 10 score, respectively.

Individual SNPs in the locus associate with KIAA1109 expression in the whole blood[34] and T cells[35], but no shared causal signal between GWAS and eQTL emerged in colocalisation analyses[36] (Dataset S5). While *KIAA1109*, in whose intron the lead variant in the GWAS study is localised, was ranked as first, only *BBS12* (Bardet-Biedl syndrome 12) gene had any colocalisation evidence - in lymphoblastoid cell lines (LCL) in TwinsUK, with the risk allele conferring reduced gene expression (PPH4 = 91%, Additional File 5: Supplementary Table 1). Despite the gene being closest to an enhancer active in epidermal stem cells and keratinocytes[5], no skin eQTLs were observed for *BBS12*.

*KIAA1109*’s function has not been well characterised but missense mutations in the gene cause abnormal brain development and arthogryposis (curving of joints) in humans[37]. *BBS12* encodes a chaperone, part of the chaperonin-containing T-complex (TRiC) assisting with protein folding and through that role mediating BBSome complex assembly, whose role is export of small vesicles to the cilia[38]

**Locus 5p13.2**

We considered 41 potential candidate genes situated within the 3 Mbp window centred on index SNP for this locus (Figure 3C & Dataset S4). Most of the cumulative score was assigned to *interleukin-7 receptor subunit alpha* - *IL7R* (score=965, which contributes to 65% of top 10 cumulative score, Table 1), followed by *SPEF2* (score=203 and a further 14%). While our GWAS results do not directly colocalise with any eQTLs for the gene (in datasets where this could be tested), there are multiple eQTL associations for SNPs in LD with the index SNP: in whole blood[34,36], CD4+ T cells[22,30,39], macrophages[40] and monocytes[31] and pQTLs in whole blood[7,21] (Figure 2, Dataset S5) as well as promoter-enhancer interactions in human embryonic stem cells[41], CD34+ hematopoietic cells[42], naïve T regulatory cells and T helper 17 cells[43] (Figure 2, Dataset S5). *IL7R* is among the genes found to be upregulated in the skin in eczema patients in a meta-analysis[16]. Our results confirm initial prioritisation of *IL7R* in the GWAS[3], which was supported by 18 credible set variants spanning the gene, including one non-synonymous (Thr>Ile) variant, rs6897932. In ADGAPP, rs6897932 was ranked as the 8^th^ most likely causal variant. The variant affects splicing of the *IL7R* transcript, with the minor C allele, the risk allele for multiple sclerosis (MS), favouring secreted over surface isoform of the protein. Elevated levels of secreted isoform exacerbate symptoms of MS in animal model[44,45]. In common with opposite effects seen in AD compared to autoimmune diseases, the minor allele is protective in eczema[10]. *IL7R* is part of the thymic stromal lymphopoietin (TSLP) receptor/IL-7/IL7R axis required for correct lymphocyte maturation, especially of Th2 lymphocytes of interest in AD, with overexpression associated with acute lymphoblastic leukaemia[46], whereas recessive mutations in the gene resulting in reduction of gene expression is seen in severe combined immunodeficiency (SCID) patients[47].

**Locus 5q31.1**

Our analysis lends support to two independent signals being present at this locus, which has been a GWAS hit in multiple immune-mediated diseases[48] (psoriatic arthritis, juvenile idiopathic arthritis, IBD and asthma) and similarly hosts two independent signals in psoriatic arthritis. The first one, represented by the primary lead SNP rs12188917 and labelled “5q31.1 - a” here, is located in the intron of *TH2LCRR*, whereas the secondary signal (labelled 5q31.1 - b) lead SNP rs4705962 is an intronic variant of *KIF3A*. Original analysis presented in Paternoster et al. (2015)[3] suggests *RAD50* and *IL5* as the top likely targets due to overlap with eQTLs in both and cytokine function of the latter, which is at odds with our ADGAPP ranking.

*Locus 5q31.1 - a*

In total, 65 genes were found within the 3 Mbp window around index SNP rs12188917 (Dataset S4). Best ranked gene was *SLC22A5* with the score of 461 (Table 1), however that only constituted 35% of top 10 total score at the locus. Second-ranked gene *IRF1* scored 303 (23% ) and 3^rd^ ranked gene *RAD50* scored 122 (9%).

The gene ranked highest at the locus, *SLC22A5* (solute carrier family 22 member 5) displayed the strongest colocalisation to be in Sun exposed-skin in GTEx (PPH4 = 94%; Additional File 5: Supplementary Table 1), but less supported colocalisation was also observed in CD8+ T cells (CEDAR, 75%), monocytes (Blueprint, 77%) and whole blood (GTEx, 58.6%), with the risk allele resulting in reduction in gene expression, which agrees with the reduction of expression seen in eczema patients[49] (Dataset S5). *SLC22A5* was also the strongest eQTL candidate in CD8+ T cells and CD4+ T cells for the locus in psoriatic arthritis[48] and in CD4+ T cells, CD15+ T cells in IBD. For 2^nd^ best gene *IRF1* (interferon regulatory factor 1), colocalisation evidence in the skin and activated monocytes is only moderate (PPH4 = 63%-64%, with increased expression in the skin associated with the AD risk allele – no direction reported for monocytes), and individual variant lookups in the interval around index SNP reveal IRF1 eQTLs only in the immune cells[30,31,34,40] (Dataset S5). Lastly, *RAD50* (RAD50 double strand break repair protein) is the only gene among the top 3 with colocalisation evidence both from coloc and TWAS, and specific to the skin (GTEx Sun-exposed and unexposed, PPH4 = 58%-81%, *p*-value = 1 x 10^-10^-7 x 10^-10^; Additional File 5: Supplementary Table 1). The eczema risk allele leads to the reduction in the expression of this gene.

SLC22A5 represents a novel candidate gene at this locus (not considered in the previous EAGLE GWAS locus annotation). It is an organic cation transporter, specialised in carnitine uptake. Especially of interest in the case of AD, inflammation increases epithelial SLC22A5 expression, presumably as a downstream effect of action of pro-inflammatory cytokines such as IFN-γ[50]. *SLC22A5’s* neighbouring paralog *SLC22A4*, also with carnitine transport function, is likely co-regulated with very similar blood cell type eQTL profile across the investigated studies (Dataset S5) and colocalisation detected also in whole blood (eQTLGen, PPH4 = 99%) and CEDAR monocytes (*p*-value of 1 x 10^-5^; Additional File 5: Supplementary Table 1), and likewise with risk allele leading to decreased expression. The gene’s polymorphisms have similarly been implicated in RA and IBD [51,52], but extensive LD in the region prevented further fine-mapping so-far.

IRF1 is worth mentioning due to its role as a master regulator of innate and acquired immune response, including activation and repression of interferon-inducible genes. This transcription factor binds to *IL4* promoter, thus promoting Th1 immunity, which is dampened in most of AD patients who displayreduced IRF1 levels[53]. Furthermore, mouse knockout mutants do not mount Th1 response on infection by Leishmania, but instead undergo Th2 differentiation, in line with Th2 response dominance in AD patients.

**Locus 5q31.1 - b**

At the 5q31.1 - b locus we tested 48 genes within the 3 Mbp interval of the index SNP (Dataset S4). Ranking at this locus showed comparable score for the two top genes: *KIF3A* (score=249, Table 1) and *SLC22A5* (score=247) – i.e. the top ranked gene at the other signal in the 5q31.1 locus; each contributing 23% of the total top 10 score. A further 13% was contributed by the third ranked gene *PDLIM4* (score=142). We observed particularly strong *KIF3A* colocalisation evidence in GTEx in the skin (sun-exposed and unexposed PPH4 = 96% and 95%; Additional File 5: Supplementary Table 1) and fibroblasts (*p*-value = 2 x 10^-11^), while *PDLIM4* colocalised well in LCL using both coloc (PPH4 = 91%) and TWAS (*p*-value = 2 x 10^-5^). The protective allele for AD was associated with increase in *KIF3A* and reduction in *PDLIM4* expression.

As the KIF3A protein provides the motor subunit to kinesin-2, it plays an important role in cilia production and motility, and its differential expression in nasal epithelial cells suggests roles in allergen clearance in asthma[54]. This hypothesis was supported by asthma-like phenotype of *kif3a* mouse knockouts showing Th2-mediated pulmonary inflammation[55]. Analogously to knockouts in the airway epithelial cells and consistent with direction of effect for risk allele seen in expression colocalisation, mouse knockouts of the gene in the skin suffer from epidermal barrier dysfunction involving increased epidermal thickness and abnormal expression of filaggrin and claudin-1[56]. They are also more prone to acquire AD following cutaneous allergen exposure. In humans, an intronic variant (rs12186803) in *KIF3A* has been shown to interact with allergens to increase risk for asthma in high-risk paediatric groups with and without AD [57]. Finally, *PDLIM4* (PDZ and LIM domain 4) is a gene promoting actin bundling and re-organisation of the actin cytoskeleton[58].

**Locus 6p21.32 and** **6p21.33**

The two loci positioned in the major histocompatibility region (MHC) represent difficult prediction targets due to notoriously intractable LD patterns in the region, which is why variant prioritisation results are not presented. That fact along with high gene density - 106 and 117 genes in the 3 Mbp intervals around index SNP in the 6p21.32 and 6p21.33 loci, respectively (Dataset S4) - makes it also less likely that our top 3 prioritised genes will contain the true genes associated with eczema. Looking at colocalisation results alone (Additional File 5: Supplementary Table 1), eQTLs for 6 and 9 genes at 6p21.32 and 6p21.33, accordingly, are found to show colocalisation with the eczema GWAS.

At the 6p21.32 locus, we prioritise various human leukocyte antigen (HLA) genes in the top 8 ranked genes (Dataset S4); they all encode HLA class II alpha or beta chain paralogues. The top hit *HLA-DRA* has a score of 1405 which corresponds to 30% of the top 10 score, the next two best ranked genes each contribute less than half of that score: HLA-DQB1 (689 - 14%) and HLA-DRB1 (566 - 13%). Interestingly, the third ranked HLA-DRB1 has been previously hypothesised as the causal gene at the locus due to the strong association of the classical HLA allele HLA-DRB1*0701 [59].

The top 3 hits at the 6p21.33 locus are poorly distinguished, with scores ranging from 173 for the first ranked *HSPA1B,* through 165 for the second ranked *HCG27* to 152 for *CSNK2B.* Heat shock protein family A (Hsp70) member 1B (*HSPA1B*) colocalizes in the following immune cell types in the CEDAR dataset: CD19+ B cells (PPH4 = 93%), CD14+ monocytes (PPH4 = 99%), CD15+ T cells (PPH4 = 85%) and CD4+ T cells (PPH4 = 75%) with the protective allele correlated with reduction in gene expression. *HSPA1B* is a negative regulator of endoplasmic reticulum stress-induced apoptosis[60]. The third hit at the 6p21.33 locus, *CSNK2B* (casein kinase 2 beta) also strongly colocalises in the CEDAR cohort: in platelets, CD14+ monocytes (PPH4 = 99.7%), CD8+ T cells (PPH4 = 99.4%), CD19+ B cells (PPH4= 99.8%) with the AD protective allele associated with reduced expression. This ER- and Golgi-targeted protein kinase has previously been associated with schizophrenia[61] and SLE in GWAS studies[20] (Dataset S5). While the index SNP for the 6p21.33 locus is located in the intron of *MICB* (MHC class I polypeptide-related sequence B) gene, we found it to be linked to the locus only through strong colocalisation in the granulocytes in the CEDAR dataset (PPH4 = 98%), where the protective AD allele corresponds to increased expression. The original Paternoster et al. (2015) prioritised MICB because of location of credible interval SNPs in the gene and differential expression in eczema patients[3].

**Locus 8q21.13**

No gene is highly prioritised at this locus out of the 31 tested within the 3 Mbp window of index SNP (Dataset S4), with the top spot occupied by *ZBTB10* with a low score of 192 (41% of total top 10 score, Table 1), followed by *TPD52* and *PAG1* (scores of 70 and 69, respectively, each contributing 15% of total top 10 score). *ZBTB10* (zinc finger and BTB domain-containing protein 10) features a suggestive and sole colocalisation hit at the locus, in TwinsUK LCL (PPH4 = 82%; Additional File 5: Supplementary Table 1) with the risk allele associated with reduced expression. Additionally, ZBTB10 is prioritised by regfm in CD8+ T cells, and the PrixFixe network enrichment method (Dataset S5). The second gene worth considering at the locus is third-ranked *PAG1* (phosphoprotein membrane anchor with glycosphingolipid microdomains 1). The gene is upregulated in *FLG* mutation-harbouring eczema patients’ skin[17], is prioritised in the regfm pipeline in the thymus, and it neighbours enhancers active in epidermal stem cells and keratinocytes[5] (Dataset S5).

Paternoster et al. (2015)[3] also tentatively prioritise *ZBTB10* as the top candidate gene due to its function as a repressor of Sp1, a transcription factor regulating many immune-related genes[62]. However, ZBTB10 transcripts were not available in the eQTL analysis of that study and so no eQTL evidence was previously presented. More recently, it has been shown that ZBTB10 is involved in alternative telomere lengthening mechanism characteristic of tumour cells[63]. We believe PAG1 is also a promising candidate as the gene’s overexpression has previously been linked to allergic disease. Vicente *et al.* (2015)[64] demonstrated that rs7009110, a lead variant at the GWAS locus predisposing to allergy as well as lead variant in the previous AD GWAS[65], and ranked 9^th^ at the current locus, is associated with *PAG1* expression in LCL in the GEUVADIS dataset. Regulatory regions were shown to act as *PAG1* transcriptional enhancers in an allele-specific fashion, with variant rs2370615’s C risk allele (ranked 59^th^ in ADGAPP) determined as a substitution likely increasing *PAG1* transcriptional activation, and at the same time disrupting binding of Foxo3a, a negative regulator of lymphocyte activation in the NF-kB pathway. However, conflicting evidence, reviewed in Vicente et al*.* (2015), exists regarding PAG1’s pro-inflammatory or anti-inflammatory role in T, B and mast cell activation.

**Locus 10p15.1**

We prioritise the *IL2RA* gene, neighbouring the lead SNP with moderate confidence at this locus. Out of 64 tested genes in the 3 Mbp neighbourhood (Dataset S4), *IL2RA* scored 333 which comprises 45% of the top 10 cumulative score (Table 1). Second-best ranked gene, *RBM17*, had 3x lower score (111 - 15%) and third best *PFKFB3* score of only 51 (7%). The locus contains GWAS-eQTL colocalisations with the top two prioritised genes in separate tissues. *IL2RA* (interleukin 2 receptor subunit alpha) shows strong evidence for colocalisation (PPH4 = 98%) in eQTLGen’s whole blood samples (Additional File 5: Supplementary Table 1), with the risk allele associated with increased expression. On the other hand, *RBM17* (RNA binding motif protein 17) colocalises using TWAS (*p*-value = 4.2x10^-6^) in the sun-exposed skin in GTEx, with the risk allele associated with decreased expression.

The original annotation in Paternoster et al. (2015)[3] supported *IL15RA* over *IL2RA* as the candidate gene for this locus due to weak eQTL overlap evidence (also present in ADGAPP) in whole blood[30,34,36,66], monocytes[31] and CD4+ T cells[67]. There was no evidence of any overlap with IL2RA eQTL evidence previously presented. *IL15RA* only scores 46 (6% of the top 10 cumulative score at this locus) in our prioritisation analysis (Dataset S4).

Regulation of IL2RA expression is complex, with at least 6 positive regulatory regions (PRRs) identified containing enhancers bound by ~8 transcription factors[68,69]. The top variant in ADGAPP ranking, rs61839660 (score=141, Dataset S4), localises to one such conserved intronic enhancer[29,41,70], contains a strong IL2RA eQTL[36] predicted to be the causal variant linking AD GWAS and IL2RA expression[71] in the blood, as well as likely to have a functional effect according to fathmm-XF[72] (Dataset S5). This variant has also previously been convincingly fine-mapped as the causal variant behind the associations with T1D at this locus[70]. It is of note that that the minor allele of rs61839660 is associated with protection from T1D, but risk of chronic inflammatory diseases, such as Crohn’s[73], SLE, allergic disease and AD[20] The causality of rs61839660 has lately been supported with a new multinomial fine-mapping approach that borrows information from GWAS on 6 different inflammatory diseases^58^.A recent landmark CRISPR activation study from Simeonov et al. (2017) present experimental evidence that rs61839660 regulates IL2RA through altering the function of an intronic enhancer, providing further evidence that this is the causal SNP behind the autoimmune signals at this locus. *Ho*wever, while the AD risk allele was shown to increase IL2RA expression in whole blood[71] (as confirmed in ADGAPP) and CD4+ T memory cells[70], the allele appears to have a negative effect on gene’s expression in T cells directly in response to antigen stimulus, delaying activation of IL2RA in the CRISPR study[73]. Deletion of the entire enhancer harbouring the SNP and partially blocking IL-2 with antibody resulted in polarisation of T cells towards proinflammatory Th17 cells, which the authors surmise would be the mechanism behind inflammatory phenotypes, such as Crohn’s disease and psoriasis.

The *IL2RA* gene encodes a subunit of the CD25 receptor for interleukin 2, which resides in another AD GWAS locus - 4q27. While we do not prioritise *IL2* as a likely candidate gene for that locus in ADGAPP, the signalling pathway in which the genes are involved is worth a closer look when investigating targets for AD susceptibility. IL-2 signalling both promotes T cell growth during the early phase of the immune response and cessation of effector T cell responses at the end of the process[68]. The first mechanism involves a positive feedback loop, whereby naive CD4+ T cells are activated towards differentiation to Th1 and Th2 cells by IL-2 binding to IL2R which then induces IL-2 production and expression of IL2R by Th1 and Th2 cells. Dysregulation in either of the nodes in the IL2-IL2RA loop could increase the propensity to AD. The second mechanism involves a negative feedback loop whereby IL-2 promotes the T regulatory cell response which suppresses the effector T cell response initiated by the first mechanism, as well as Th17 differentiation through IL-2 sequestering[74]. Missense and frameshift mutations in *IL2RA* mimic Immunodysregulation polyendocrinopathy enteropathy X-linked (IPEX) syndrome which is one of the monogenic diseases illuminating aspects of AD phenotype[75] due to featuring eczematous skin and immune dysregulation[76].

**Locus 10q21.2**

Limited breadth of evidence contributes to gene prioritisation at this locus, where we evaluate 39 genes within the 3 Mbp interval around index SNP (Dataset S4). While the score of the top ranked gene, *ADO* (2-aminoethanethiol dioxygenase)*,* is 615 which makes up 61% of the top 10 gene cumulative score (Table 1), it is prioritised solely on the basis of association with the biggest number of individual variants in the whole blood[36] and monocyte[31] eQTL studies rather than formal colocalisation test as well as variants overlapping promoter regions interacting with putative enhancers in a variety of blood immune cells and associating with cardiac health phenotypes in GWAS Catalog (Dataset S5). The second and third ranked genes: *ZNF365* (zinc finger protein 365) and *EGR2* (early growth response 2) possess comparably lower scores of 101 and 90 in ADGAPP, i.e. each contributing 9-10% of cumulative score for top 10 ranked genes. Previous gene prioritisation in Paternoster et al. (2015)[3] supported *ZNF365* as the causal gene since lead SNP and credible set variants span the gene’s intronic region, where an IgE GWAS signal was previously fine-mapped to[77].

ADO encodes a thiol oxidase which catalyses the conversion of amino-terminal cysteine to cysteine sulfinic acid and shows homology to plant cystein oxidases involved in hypoxia response. In human cells, it has been shown to participate in signalling in the oxygen-regulated N-degron pathway of protein degradation as an enzymatic oxygen sensor[78].

*ZNF365* (zinc finger protein 365) encodes multiple isoforms that have been variously linked to regulation of neurogenesis, maintenance of genome stability and uric acid secretion[79]. The signal in Crohn’s disease GWAS locus has been fine-mapped to a nonsynonymous variant within *ZNF365*[80] and (based on position) the gene has been associated with several divergent phenotypes in the GWAS Catalog, ranging from breast-related, neural to chronic inflammatory disease and allergy.

Based on evidence derived solely from literature review, the third top candidate, *EGR2* appears most directly related to AD phenotype. EGR2 regulates the balance between clonal expansion and differentiation of CD4+ and CD8+ T cells[81], tipping it towards the former. Furthermore, EGR2 has been found to be a key repressor of Th17 cell pro-inflammatory program via inhibition of BATF activation[82]. Gene knockout mice show increased production of IL-17 cytokines by naïve CD4+ T cells, differentiation into Th17 cells and reduced propensity for chronic autoimmune response in the central nervous system[82,83].

**Locus 11p13**

This locus stands out as having one clearly prioritised target gene - *PRR5L* (score=598, 79% of top 10 cumulative score; Table 1), with its final score nine times higher than the second-ranked gene, *TRAF6* (score=65 - only 9%). In total, we considered 15 genes located in the 3 Mbp region around index SNP (Figure 3D & Dataset S4). We could not tease apart the target behind the secondary signal represented by lead SNP rs12295535, with the same genes prioritised as for the primary signal represented by rs2592555. rs2592555, the top prioritised variant at the locus (score=246, Dataset S4), is situated in the intron of *proline rich 5 like (PRR5L)*. We also note that ADGAPP’s second best SNP rs7925585 (score=132) for the primary signal, also positioned within the intron of *PRR5L*, was independently prioritised for eczema in FINDOR analysis[28]. Strong expression colocalisation was detected with the coloc method (Figure 2, Additional File 5: Supplementary Table 1) in: eQTLGen - whole blood (PPH4 = 95%); TwinsUK – skin (PPH4 = 91%), LCL (PPH4 = 98%); CEDAR - CD4+ T cells (PPH4 = 98%) with protective AD allele associated with increased expression. The gene’s role was also supported using the network method PrixFixe (Dataset S5) and strongest methylation signal detected through mQTL overlap with locus interval SNPs in whole blood in the GoDMC study[15]. *PRR5L* was proposed as the candidate gene in the EAGLE GWAS due to position of the lead SNP in *PRR5L’s* intron and *PRR5L* eQTL overlap with the credible set variants[3]. However, there was previously very little evidence for colocalisation of these signals.

PRR5L is part of the rapamycin complex 2 (mTORC2) which responds to extrinsic stimuli through cytoskeleton re-organisation and cell migration[84]. PRR5L specifically plays a role in regulation of fibroblast migration, and decreased expression of the gene conferred by the risk allele is predicted to lead to increase in fibroblast migration.

**Locus 11q13.1**

No single strongly prioritised candidate gene was highlighted among the 165 genes at this locus in ADGAPP (Dataset S4). The gene with the highest score of 336, *CTSW*, contributed 23% of the total score for top 10 ranked genes (Table 1), while the 2^nd^ ranked and 3^rd^ ranked *OVOL1* and *EFEMP2* scored 236 (16%) and 168 (11%), accordingly. CTSW (cathepsin W) is differentially expressed in the skin between eczema patients and controls[17,85], and numerous eQTLs in naive[39,67] and stimulated T cells[67] – including Th1, Th2, Th17, Treg, Treg memory cells, macrophages[40], and whole blood[34,36] are overlapped by variants in the locus interval, including evidence for allele-specific expression in T [22,86] and NK cells[30] (Dataset S5). However, unlike the second-ranked gene, *OVOL1* (Ovo like transcriptional repressor 1), there was no evidence of colocalisation in the datasets where this could be tested.

*OVOL1* eQTLs boast highly confident colocalisation with the AD signal in lymphocytes in GTEx (PPH4 = 99.9%; *p*-value = 2 x 10^-18^; Additional File 5: Supplementary Table 1) as well as TwinsUK using both coloc (PPH4 = 99.9%) and TWAS (*p*-value = 8 x 10^-18^) methods, with the AD risk allele correlated with downregulation of the gene. While *OVOL1* is not known to be differentially expressed in eczema, it shows differential expression between epidermal stem cells and differentiated keratinocytes with the index variant shown to lie in an enhancer contacting the gene’s promoter[5,87] (Dataset S5). The third top candidate, *EFEMP2* (EGF-containing fibulin extracellular matrix protein 2) shows weak colocalisation (PPH4 = 58%) in CD15+ granulocytes in CEDAR (Additional File 5: Supplementary Table 1), and independent variant lookups in the locus reveal eQTL overlap in whole blood[34,36], neutrophils[22], monocytes[30], naive T cells[39], macrophages[40] – similar pattern to that of *CTSW*. Likewise, *EFEMP2* is upregulated in keratinocytes versus epidermal stem cells[5] but also in eczema patients[17,85] (Dataset S5).

Prioritisation of *OVOL1* as the candidate gene at the locus in Paternoster et al. (2015)[3] relied on location of the index SNP rs10791824 within the intron of *OVOL1,* indirect evidence for regulatory role in AD-relevant cell types and mouse knockout phenotype. The mouse mutants are characterised by hyper-proliferation of keratinocytes, hair shaft abnormalities, delayed epithelial barrier formation and kidney cysts[88,89]. ADGAPP results supporting the rs10791824 variant as the most likely causal SNP (rank 1 with score of 968, Dataset S4) lend further support for potential importance of *OVOL1* at this locus. The variant shows high posterior probability (0.75-1) of being the causal SNP behind the eQTL and GWAS colocalisation signal in all the datasets tested (Additional File 5: Supplementary Table 1), and likewise through fine-mapping in Finemap, JAM and Paintor programs (Dataset S5). Its role within an enhancer is supported by Roadmap classification[29], Blueprint hQTLs for K4ME1 and K27AC[5,22,90] in blood and epidermal cell types, interaction with OVOL1 promoter in stem cell and B lymphocyte 3C data[41,91], localisation within a FAIRE-seq peak in a study of epidermal disruption[92], overlap with CTCF motif ChIP-Seq peaks in LCL[93,94], computational enhancer prediction models[95] and moderate fitness consequences[96]. Additional lookups revealed the variant to be bound by lymphocyte-specific transcription factors, such as BATF and IRF4 in ENCODE Chip-Seq[97].

Literature evidence shows the first-ranked *CTSW* to be a papain family cysteine proteinase found exclusively inside the endoplasmic reticulum of natural killer and cytotoxic T-cells, yet not essential for cytotoxicity[98]. The gene is upregulated by IL-2, which together with IL2 receptor may be prioritised as candidate genes for AD GWAS loci 4q27 and 10p15.1, respectively. Second-ranked *OVOL1* induces upregulation of filaggrin and loricrin via aryl hydrocarbon receptor activation in human keratinocytes and through binding to c-myc promoter[99,100], which provides a direct link to known atopic dermatitis gene targets. Further, nuclear translocation of OVOL1 is reduced in AD skin and normal skin treated with IL-4 (a cytokine in the Th2 pathway active in AD) providing more links of the gene to the disease. Finally, third-ranked *EFEMP2* is a member of the fibulin family of extracellular matrix proteins whose mutations cause collagen mis-assembly in humans and mice, especially affecting the skin (cutis laxa) and cardiovascular system[101].

**Locus 11q13.5**

Two genes are equally highly prioritised in ADGAPP at this locus, out of 40 genes in the 3 Mbp interval around the index SNP (Dataset S4). They are *LRRC32* and *EMSY* (previously known as *C11orf30*) which score 545 and 521, correspondingly and together make up 84% of the top 10 genes at the locus (Table 1) – for comparison, the third ranked gene (*THAP12)* had a score of only 47. The 1^st^ ranked gene *LRRC32* (leucine rich repeat containing 32) possesses the strongest colocalisation evidence, albeit in one tissue – eQTLGen whole blood at 98% posterior probability of colocalisation (Additional File 5: Supplementary Table 1), EMSY displays weaker colocalisation signal in Blueprint’s monocytes at 89%. In both cases, the AD risk allele confers reduced expression of the genes. Both genes have been shown as targets of differential enhancer interaction between naive T helper 17 cells and T regulatory cells[102] and individual variant eQTL lookups in whole blood - LRRC32 of healthy subjects[36], while EMSY of patients with RA[103] (Dataset S5). However, only LRRC32 is linked to allele-specific expression QTLs in monocytes[22] and active enhancers in keratinocytes[5]. On the other hand, EMSY shows conflicting differential expression results in the skin of eczema patients: it is upregulated in homozygous filaggrin mutation AD patients in Winge *et al.* (2011)[17], while downregulated in compound heterozygote FLG AD patients in Cole *et al.* (2014)[49].

No evidence was available to prioritise either *LRRC32* or *EMSY* as the candidate gene in Paternoster et al. (2015)[3], and all the credible set SNPs including the index SNP spanned the intergenic region between the two genes. Since then, both genes have been the focus of experimental validation[104] ^,^[105] which agrees with possible non-exclusive roles of *LRRC32* and *EMSY* in the immune T cells and skin, respectively. GARP’s (protein encoded by *LRRC32*) role in AD and inflammatory disease in general has received convincing experimental support[106]. GARP is a cell surface receptor present on activated T regulatory cells, which is bound by latent transforming growth factor-β (TGF-β). Six rare missense mutations have been identified in AD patients in a targeted resequencing study with predicted coding changes affecting protein folding and post-translational modification[104]. Characterisation of one of the substitutions, the A407T variant, revealed reduced expression of the protein on the cell surface along with TGF-β latency-associated protein, and consequently reduced differentiation of T cells into T regulatory cells in the carriers. This agrees with GARP's role in promoting Treg activity established via shRNA and antibody knockdowns[107], acting through its effect on TGF-β maturation and activation. More recently, GARP’s expression has been shown to be downregulated in another allergic disease: eosinophilic esophagitis and upregulated in oesophageal epithelial cell lines on treatment with IL-13, supporting a putative role in regulating Th2 response[108].

While data available to us does not provide any colocalisation evidence for significant role in the skin, possibly due to low expression range and hence low power for eQTL detection, common to transcriptional regulators, EMSY has recently been characterised as a potent regulator of skin barrier formation[109]. siRNA knockdown of the gene in skin organotypic culture leads to increased *FLG* expressiona and enhanced barrier function, whereas overexpression in primary human keratinocytes leads to reduction in mRNA and protein markers of barrier formation. Lastly, EMSY has been shown to activate TSLP and CCL5 expression in eosinophilic esophagitis; both genes are known to contribute to inflammatory response also in atopic dermatitis[16,24,110–115].

**Locus 11q24.3**

Amongst the 14 genes within the 3 Mbp window at this locus (Dataset S4), ADGAPP prioritised *ETS1*  with the score of 298 (75% of cumulative top 10 score, Table 1), in contrast to the scores of only 35 (9%) and 18 (5%) for the 2^nd^ (*FLI1*) and 3^rd^ (*APLP2)* ranked genes. We find modest colocalisation support for *ETS1* (ETS proto-oncogene 1, transcription factor) in whole blood in eQTLGen (PPH4 = 48%, Additional File 5: Supplementary Table 1) and stimulated monocytes (PPH4 = 75%) with the risk allele correlating with increase in gene expression. Individual lookups linked variants in the locus interval to eQTLs in the whole blood[34,36], CD4+ T cells[30] and neutrophils[116] and promoter-enhancer interactions in embryonic stem cells for *ETS1*[41] (Dataset S5). In addition, *ETS1* showed consistent differential expression in AD patients regardless of *FLG* genotype. Weak colocalisation evidence exists for *FLI1* (Fli-1 proto-oncogene, ETS transcription factor) in in Blueprint monocytes and neutrophils (54% and 46%, respectively, Dataset S5) suggesting increased expression to be associated with the AD risk allele, and individual eQTLs overlap variants in LD with focal SNP in monocytes *FLI1*[30], whereas and in LCL and CD34+ hematopoietic cells[91] for promoter-enhancer interactions.

Initial candidate gene suggestion in Paternoster et al. (2015) was for *ETS1* due to its proximity (141 kbp downstream of *ETS1*) to the fine-mapped index SNP rs7127307 and prolific immunity-related functions of this transcription factor. ADGAPP analysis likewise highlights rs7127307 as being the likely causal variant at this locus (score=310, Dataset S4) through finemapping runs in Finemap, JAM and Paintor with posterior probability of 1, 0.7 and 0.31, respectively. The variant also shows the highest posterior probability of being the causal SNP for colocalisation with *ETS1* eQTLs in stimulated monocytes and is situated in conserved genomic regulatory block of enhancers active only in immune cell types and fetal adrenal gland[117].

ETS1 has been implicated in a wide variety of functions, where it was shown to act as co-activator; however in its role in immune dysregulation, it acts as a negative regulator of inflammatory Th17 cell responses and B cell differentiation but also a positive regulator of Th1 polarity[118] acting through downregulation of the IL-6 pathway[119] – specifically, a signalling subunit for the IL-6 receptor complex gp130 protein (see also locus 1q21.3). This hypothesised role was further confirmed by increased phosphorylation of STAT3 (see also locus 17q21.2) further downstream in the Th17 pathway and increased IL-17 and IL-22 production in *Ets1*-deficient CD4+ T cells. *ETS1* gene expression levels are decreased both in atopic dermatitis and skin autoimmune disease - SLE, and *Ets1* knockout mouse phenotype mimics aspects of molecular phenotypes of both diseases[120,121]. Putative role of ETS1 in AD is also underlined by reduced IL-2 levels further affecting Th17/Treg imbalance[122] and dampened expression of skin barrier genes: filaggrin, claudin-1 and loricrin in *Ets1*-deficient CD4+ T cell mouse model. On the other hand, overexpression of *ETS1* in keratinocytes of stratified epithelium in a mouse model[123] resulted in profound abnormalities in the dermis driven by increased keratinocyte proliferation but reduced terminal differentiation: hyperplasticity, reduced granular layer, parakeratosis, compact stratum corneum and particularly interestingly for AD, increased permeability.

The 11q24 locus has been independently linked to other inflammatory diseases, amongst others, general atopy[24], RA[124], SLE[125], psoriasis, IBD and celiac disease[20]. Most of the variants map in the 3’ region of the *ETS1*, but some are placed in the region between *ETS1* and *FLI1*, so *FLI1,* another transcription factors in the ETS family, should not be dismissed as a candidate gene. *FLI1* regulates collagen deposition by repressing collagen type 1 gene and is constitutively epigenetically silenced in systemic sclerosis (scleroderma) [126], which is key to recapitulating the disease symptoms, which centre on fibrosis of the skin and organs and Th2/Th17-skewed immune activation[127].

**Locus 12q15**

At this locus (identified in the MAGENTA gene-set analysis of the GWAS), ADGAPP strongly prioritises an unexpected target – *MDM1* (score=728, 70% of the top 10 cumulative score, Table 1) rather than the cytokine *IL22* immediately neighbouring the index SNP (score=99, ranked 2^nd^ ) or the immune modulator *IFNG*, whose assumed involvement led to the identification of this locus in the GWAS (score=57, ranked 3^rd^). Altogether 34 genes were considered at the locus, within the 3 Mbp window of the lead SNP rs2227483 (Dataset S4), and 3 out of 4 closest genes are involved in immune signalling (*IL22, IFNG and IL26*). However, the top prioritised gene is the fourth-closest gene, positioned 100 kbp away - *MDM1* (Mdm1 nuclear protein). Hundreds of individual significant whole blood[34,36,103,128] and immune cell subtypes[22,30,31,67,86,129,130] eQTL lookups among interval variants, promoter-enhancer interactions target *MDM1* [91,131], and only promoter-enhancer interaction evidence[5,91,131] is present in ADGAPP for *IL22* (Dataset S5); similarly there is promoter-enhancer interaction evidence for *IFNG* but with additional prioritisation by PrixFixe network method and modest colocalisation support in activated monocytes (PPH4= 69%, direction of effect not reported)[132].

*MDM1* shows enhanced expression in multi-ciliated epithelial cells, where it localizes to centriole barrel and binds microtubules[133]. Naturally occurring mouse knockdown in the gene exhibits age-related retinal degeneration[134]. On the other hand, gene functional definitions and pathway positions rule in favour of the 2^nd^ and 3^rd^ ranked *IL22* (interleukin 22) and *IFNG* (interferon gamma, *IFN-γ*) as being more credible targets to influence AD phenotype. The expression of the Th22 cytokine IL-22 and IL-22^+^CD8^+^ T-cell frequency in adult skin lesions is positively correlated with AD disease severity characterised by increased epidermal hyperplasia and skin barrier impairment[115,135,136]. IL-22 along with other cytokines in the family induces cell proliferation, migration and drives the expression of powerful inflammatory and antimicrobial molecules in keratinocytes, while at the same prevents their terminal differentiation[137], in line with reduced expression of genes essential for cornified envelope formation, such as filaggrin. Improvement in AD severity in patients treated with IL-22 antibody (fezakinumab) gives credence to the idea that *IL-22* is the causal gene at the locus[138]. In addition, *IFNG* could also be a plausible candidate gene, given the paramount role of this cytokine in innate and adaptive immunity. Similarly to IL-22, its levels positively correlate with SCORAD scores in intrinsic AD patients[139]. As a Th1 cytokine, IFN-γ plays an important function in intrinsic AD showing higher Th1 skewing[140]. IFN-γ production by Th1 cells in the dermis promotes apoptosis of keratinocytes which leads to formation of lesions[141]. At the same time, release of IFN-γ-inducible chemokines by apoptotic keratinocytes further aggravates T cell infiltration of the dermis in a positive feedback loop resulting in increased keratinocyte apoptosis and inflammation[142]. On the other end of the endotype spectrum, particularly low expression of IFN-γ and its receptor was found among a subset of AD patients susceptible to viral skin infection eczema herpeticum[143].

**Locus 14q13.2**

Out of 70 genes considered as causal at this locus (Figure 3E & Dataset S4), we find 2 of them to have comparably high scores - *Protein phosphatase 2 regulatory subunit B''Gamma* (*PPP2R3C*) with the score of 996 (31% of top 10 cumulative score, Table 1), and *KIAA0391* with the score of 814 (25% of top 10 cumulative score).

Three (PP2R3C, KIAA0391, *FAM177A1*) out of the four top-ranking ADGAPP genes at this locus display partial co-expression. The top-ranked candidate gene *PPP2R3C* shows colocalisation (Figure 2, Additional File 5: Supplementary Table 1) in sun-exposed (PPH4 = 94%) and unexposed skin (PPH4 = 95%, *p*-value = 2 x 10^-7^ ), whole blood in GTEx (PPH4 = 97%, *p*-value = 2 x 10^-7^), CD15+ granulocytes (PPH4 = 96%) and colon (PPH4 = 96%) in CEDAR, and neutrophils (PPH4 = 93%) in Blueprint. The next best gene, *KIAA0391* recapitulates the colocalisation in the skin in GTEx (sun exposed PPH4 = 97%, unexposed PPH4 = 96%), and LCL and individual immune cell types: LCL from TwinsUK (PPH4 = 94%, *p*-value = 3 x 10^-9^), CD8+ T cells (PPH4 = 97%) and CD14+ monocytes from CEDAR cohort (PPH4 = 60%, *p*-value = 1 x 10^-4^), in addition to the spleen in GTEx (PPH4 = 95% and *p*-value = 2 x 10^-7^). Moreover, we find hundreds of individual variants in LD with the index SNP to overlap blood and skin eQTLs for those genes (Dataset S5). The orchestrated expression of these genes is underscored by their differential expression in atopic dermatitis skin: *PPP2R3C* is upregulated regardless of *FLG* genotype[17,144] and *KIAA0391* is strongly downregulated in homozygous *FLG* mutation AD patients[17]. Comparing that with our colocalisation results, we see tissue-specific regulation for *PPP2R3C,* upregulation in the skin (in line with the differential expression results) but downregulation in the blood associated with the AD risk allele. For *KIAA0391*, the downregulated expression seen in AD patients is at odds with the eQTL result, where the AD risk allele is associated with increased expression; this conflict could indicate that *KIAA0391* is not an AD susceptibility gene. The original GWAS annotation[3] also suggested *PPP2R3C, KIAA0391* and *FAM177A1* as plausible causal genes, with the 47 credible interval SNPs scattered throughout the three genes and the lead SNP mapping to an intron within *PPP2R3C*; strong colocalisation with TwinsUK microarray eQTLs in that paper was found only for *KIAA0391*.

Considering gene function, *PPP2R3C* is the most viable candidate. Targeted B-cell mouse knockout mutants show profound abnormalities in humoral immune response: reduced B cell proliferation, maturation, abnormal activation and small spleen[145]. Similarly, loss of *PPP2R3C* in T cells results in atrophy of the thymus, decreased thymocyte abundance, especially of CD4+ and CD8+ double-positive thymocytes[146]. Variants at the locus related to pleiotropic phenotype of chronic inflammatory diseases (AS, CD, psoriasis, primary sclerosing cholangitis, UC)[147] map to intron positions within *PPP2R3C*. Next, linking the gene’s mutant phenotype established in mouse to human GWAS, it has been reported that decrease in lymphocyte counts[148] significantly correlates with the risk allele at rs2038255, the index variant in AD GWAS and ADGAPP top prioritised variant (score=389, Dataset S4). *PPP2R3C* encodes a regulatory subunit of protein phosphatase 2A, known as G5PR, which associates with phosphatases PP2A, PP5, GANP protein and represses JNK and IKK*β* (inhibitor of NF-*κ*B) phosphorylation[149]. It is involved in regulating antigen-based B-cell and early T cell selection in the thymus promoting thymocyte and B cell survival. G5PR becomes upregulated in activated B cells and prevents B-cell receptor-mediated activation-induced cell death in B cells through suppression of late-phase JNK activation[150]. Overexpression results in the increase of production of non-specific B cells after immunisation and generation of autoantibodies in non-stimulated mice[151]. Therefore, *PPP2R3C* upregulation in the skin could be a contributing factor to autoimmune activation seen in a subset of severe AD patients and particularly directed against epidermal proteins[152].

*KIAA0391* in contrast does not appear to be so directly functionally linked to the AD phenotype. It encodes a component of mitochondrial RNase P complex which catalyses the last step in pre-tRNA maturation process: removal of the tRNA 5’ leader sequence[153].

**Locus 14q32.32**

Accounting for 55% of the cumulative score at the locus, *TNF receptor associated factor 3 (TRAF3)* is clearly prioritised among the 59 genes positioned within 3 Mbp of the index SNP at the locus (Figure 3F & Dataset S4). While TRAF3’s score is 848, the second-ranked AMN scores only 281 (18%) (Table 1). *TRAF3* is the only gene at the locus with direct colocalisation evidence (Additional File 5: Supplementary Table 1): in the whole blood in eQTLGen (PPH4 = 93%) and with lower confidence (PPH4 = 85%) in CD4+ T cells in Blueprint. In addition, many locus interval SNPs are possibly situated within an enhancer interacting the gene’s promoter in human embryonic stem cells[41], whole blood[154], CD34+ hematopoietic cells and lymphoblastoid cell lines[42], naïve T regulatory cells and T helper 17 cells[43] and epidermal stem cells along with keratinocytes[5] as shown by Hi-C data. In AD GWAS, risk alleles correlate with upregulated expression of TRAF3 and in IBD there is increased expression of the gene in inflamed intestinal mucosa[155]. However, changes in expression have not been consistently observed in AD lesions relative to healthy skin (Dataset S5).

The original EAGLE GWAS annotation also presents *TRAF3* as the candidate gene at the locus due to the location of the index SNP, pathway enrichment in MAGENTA gene set analysis and mouse knockout phenotype, but no eQTL evidence[3]. Two out of the 3 top prioritised SNPs at the locus in ADGAPP (1^st^ ranked rs79589176 and 3^rd^ ranked rs12880641) as well as index SNP (rs7146581) are intronic, situated within the *TRAF3* gene, whereas the remaining top 3 SNP (2^nd^ ranked rs71421262) is 2 kbp 5’ upstream of *TRAF3*. *TRAF3*’s role in signal transduction in immunity is well-established, with early studies describing a serious imbalance in T cell composition in mouse model knockouts which eventually leads to their perinatal death[156]. TRAF3 is a repressor of CD40- and B cell-activating factor-mediated signalling and limits homeostatic B cell survival[157]. *TRAF3* is also related to possible candidates at two other GWAS loci (*TRAF6* at 11p13 and *IL6R* at 1q21.3)*.*  TRAF6 positively regulates MAPK signalling and production of inflammatory cytokines and chemokines, whereas TRAF3 needs to be degradatively ubiqutinated during MyD88-dependent Toll-like receptor signalling to activate the JNK and p38 MAPK cascade[158]. TRAF3 exerts a negative effect on Th17-based inflammation by sequestering IL-17R, however, normally this is prevented by competitive binding of TRAF3 by NRD1. This allows formation of IL17R-Act1-TRAF6 complex and subsequent propagation of IL-17-induced signal down through MAPK and NF-κB pathways leading to production of pro-inflammatory molecules, including cytokine IL-6, whose receptor is prioritised in the AD GWAS at locus 1q21.3[159].

**Locus 16p13.13**

Out of 143 considered genes at the locus within the 3 Mbp interval of index SNP (Dataset S4), two top genes are prioritised equally strongly– *DEXI* (score=376, Table 1) and *CLEC16A* (score=364). Their combined score contributes to 67% of the cumulative top 10 gene locus score and the third ranked gene *RMI2* contributes only a further 10% (score=108). Colocalisation evidence at the locus is limited and prioritises *DEXI* (dexamethasone-induced protein) and *CLEC16A* (C-type lectin domain containing 16A) in CEDAR’s rectum tissue based on TWAS and modest support by coloc (PPH4= 73%), respectively. The protective AD allele confers increased expression of both genes. *DEXI* and *CLEC16A* are precisely the two genes with correlated gene expression across cell types that have been championed as the top candidate genes at this locus in association studies of inflammatory and autoimmune disease, such as T1D, MS, alopecia areata, SLE, asthma[20] (Dataset S5). Risk alleles of autoimmune disease-linked variants (T1D, MS) in the locus are associated with lower *DEXI* expression in monocytes, thymus and LCLs[160], which mirrors the effect of protective AD allele conferring higher *DEXI* expression. *CLEC16A* is differentially expressed in eczema: it is highly downregulated in *FLG* mutant eczema skin patients relative to healthy controls[17]. However, comparison of lesional vs non-lesional skin in AD patients reveals small upregulation of the gene[144].

*DEXI* represents a novel candidate gene at this locus. The original AD GWAS annotation in the EAGLE paper[3] presented *CLEC16A* as the candidate gene at the locus due to location of the two SNPs from the fine-mapped credible set within the gene’s intron and binding of STAT3 (see locus 17q21.2) in mammary epithelial cells as well as top RegulomeDB score at the lead SNP[161]. In ADGAPP, we too prioritise the index SNP rs2041733 (ranked 1^st^, score of 102; Dataset S4) thanks to strong fine-mapping evidence in Finemap and JAM (Dataset S5) but also some evidence from eQTL colocalisation (posterior probability of being a causal variant > 0.5). Regulatory function of the SNP is supported by overlap with a strong caQTL affecting regulatory DNA accessibility[162] across Roadmap cells and strong hQTL for K4ME1 in monocytes[22] and skin cells[5].

Direct looping interaction from lead SNP-harbouring *CLEC16A* intron to *DEXI* promoter has been shown with 3C in monocyte-like cell line, lung epithelium and LCL, suggesting that *CLEC16A* intron contains a *DEXI* enhancer[163]. Nevertheless, these variants are situated within intron 19 of *CLEC16A,* about 29 kbp away from the top AD SNPs (intron 23) so they could well be tagging an independent GWAS signal, as 3 independent signals at the locus have been suggested in MS[164]. CRISPR-Cas9-disrupted *DEXI* expression precipitates early onset of diabetes in T1D mouse model and it has been postulated that DEXI impacts T1D risk through changes to host metabolites and microbiome[165]. Specific targeting of beta cells in pancreas to knock down DEXI revealed reduction in interferon β expression followed by lower production of inflammatory chemokines on exposure to synthetic dsRNA and reverse in the case of DEX1 overexpression, pointing to roles in antiviral immune response[166].

*CLEC16A* is a much better characterised gene with multifarious posited functions related to autophagy and secretion. Initially, it was identified as an endosomal protein promoting murine mitophagy by interacting with the Nrdp1 E3 ubiquitin ligase, in turn a binding partner of Parkin, the key regulator of mitophagy[167]. Loss of *CLEC16A* in that context results in inefficient elimination of abnormal mitochondria through autophagy and predisposes to inflammation[168]. CLEC16A also likely negatively regulates starvation-induced autophagy[169]. Next, knockdown of the gene in mice led to halving of the number of B cells and elevated IgM levels[170]. CLEC16A functions also impact other lymphocytes: CLEC16A limits NK cytotoxicity[168,171] and fine-tunes thymic T cell selection by regulating thymic epithelial autophagy[172]. In general, the gene’s expression has a negative impact on cell surface receptor expression and cytokine (such as interferon-γ) and chemokine release. Unlike in AD, where atopic dermatitis risk correlates with low expression of *CLEC16A* in the skin tissue, patients with autoimmune MS display increased *CLEC16A* expression in monocytic and dendritic cells[173]. Of importance in MS, CLEC16A positively regulates HLA-II antigen presentation pathway through participation in late endosomal biogenesis and trafficking via binding members of the dynein complex[173].

**Locus 17q21.2**

ADGAPP was not able to provide much evidence for a single candidate gene at this locus, out of 147 genes within the 3 Mbp window of index SNP (Dataset S4). The top ranked gene *DHX58* scored only 254 (32% of the top 10 cumulative score, Table 1), while the 2^nd^, 3^rd^ and 4^th^ ranked *STAT3*, *RAB5C* and *CAVIN1* 101, 100 and 94, in that order, made up a further 37% of the cumulative score. Tentative eQTL-GWAS colocalisation evidence is only present for *STAT3* (signal transducer and activator of transcription 3) at PPH4= 50% in Sun-exposed skin in GTEx (Additional File 5: Supplementary Table 1) and for the 4^th^-ranked *CAVIN1* (caveolae associated protein 1) gene, similarly in GTEX Sun-exposed skin (68%) but also in TwinsUK LCL (55%). In both cases, the risk allele is predicted to lead to decrease in gene expression. However, it is *DHX58* (DExH-box helicase 58) that occupies the first position in the ADGAPP ranking due to multiple individual eQTLs overlapping the locus interval: in whole blood[36,128], allele-specific expression in monocytes[31] and T cells[22,39], in the skin[174], B cells and LCL[30,130], NK cells[30], Tfh cells, Treg memory cells[67] (Dataset S5). Fewer significant eQTLs in a smaller number of tissues were found for *STAT3*: whole blood[128], naive and induced monocytes[31], CD8+ T cells, NK cells[30]. Again, slightly different profile is seen for *RAB5C* (Ras-related protein Rab-5C), with eQTLs in: whole blood[36], B cells, CD4+ and CD8+ T cells, monocytes[30], macrophages[40], Tfh, Th17, Th2 cells[67]. Lastly, variants in the GWAS locus interval are also associated with the expression of *CAVIN1* (caveolae associated protein 1) in the whole blood[36], CD4+ T cells[30], and Th1 cells[67]. *CAVIN1* is the only gene of the four, where we could find evidence of differential expression in AD patients: strong downregulation in lesional skin of patients with FLG mutation relative to healthy controls[17].

The original Paternoster et al. (2015)[3] post-GWAS annotation focused on *STAT3* since the lead SNP is located in the intron of the gene, and further support was provided by eQTL lookups in relevant tissues, evidence for regulatory function of SNPs in the credible set interval and background knowledge on STAT3 function. The index SNP rs12951971 comes up top in ADGAPP too (score of 126, Dataset S4) as evidenced by high posterior probability of causality in Finemap and JAM runs and high effect mQTL mapping to STAT3. The second best SNP, rs4796791 (score=44) and also with intronic location in *STAT3*, is too supported by Finemap runs, overlaps a sentinel variant in a MS GWAS[175] and strong effect STAT3 eQTLs in whole blood[128], monocytes[86] and mQTL in whole blood.

The top prioritised hit *DHX58* encodes the LGP2 protein is involved in regulating innate immune response to virus infection[176]. LGP2 negatively regulates IFN during two early stages in the induction of interferon production[177] but has also a positive role in antiviral response through facilitating viral dsRNA recognition[178]. *STAT3*, our second ranked gene is a ubiquitous regulator and signal transducer in JAK-STAT signalling pathway, with many functions in immunity, including cancer and inflammatory skin disease, such as psoriasis and eczema[179]. Consequently, here we focus on summarising established associations of STAT3 with AD.

First of all, STAT3 is activated downstream of cytokines important for AD: IL-4, IL-13, IL-22, IL-23, IL-12, IL-6, IL-10 and IL-31[180,181]. One example of activation of STAT3 by cytokines would be in terminal keratinocyte differentiation in normal skin barrier formation, where the IL-4/IL-13-JAK-STAT3 pathway has been shown to be the master regulator. STAT3 signalling is thought to contribute to TSLP-dependent downregulation of filaggrin and anti-microbial peptides in keratinocytes in AD[182–184]. JAK-STAT3 signalling also plays a role in mediating pruritogenic effect of IL-31 in eczema[185]. Other relevant functions of STAT3 in related diseases include: promotion of dermal fibrosis in scleroderma through increased collagen and fibronectin deposition in the extracellular matrix[186] and skewing towards the IL-23/IL-17 axis in psoriasis through STAT3-dependent induction of the IL-23 receptor, RORα, RORγ and IL-17 expression[179,187,188]. Indeed, STAT3 is activated in lesions of psoriatic patients and a mouse mutant with keratinocyte-specific constitutive expression of activated STAT3 serves as a model of psoriasis[189].

On the other hand, our eQTL and colocalisation analysis links the AD risk allele to reduced expression of *STAT3*. Loss-of-function mutations in *STAT3* result in autosomal dominant hyper-IgE syndrome (HIES) featuring high IgE levels and eczema-like skin lesions plagued by recurrent infections arising due to Th1/Th2 imbalance similar to seen in AD[190,191]. However, HIES subjects entirely lack IL-17-producing Th17 cells[192]. Counter-intuitively, STAT3 serum levels and gene expression are elevated in early-onset pediatric eczema[193] relative to late-onset adult eczema cases and correlate with epidermal barrier function[194]. This could partially be due to immune activation of early-onset acute eczema resembling that of psoriasis in terms of Th17 polarisation[194,195].

The third-best prioritised gene, *RAB5C*, is primarily involved in controlling endocytosis, formation of adhesion foci and cell motility[196,197]. The fourth-ranked gene *CAVIN1* is highly expressed in fibroblasts, adipocytes, epithelial and endothelial cells where it is necessary for formation of caveolae, specialised invaginations of the plasma membrane with roles in signal transduction, lipid transport and clathrin-independent endocytosis[198,199]. Mice knockouts in the gene suffer from lipodystrophy and altered lung morphology[200], while frame-shift mutations in humans cause congenital generalized lipodystrophy type 4 with muscle rippling[201]. CAVIN1 participates in membrane repair mechanism by binding cholesterol exposed during injury and acting as a nucleation site for forming a membrane patch[202]. Of interest in AD, expression of *CAVIN1* has been linked to asthma and chronic inflammatory respiratory disease for its antifibrotic role as well as maintaining E-catherin-based cell-cell adhesion and therefore airway epithelium integrity which is diminished in scledorema patients and asthmatics[203,204], respectively. There is also evidence of role of post-translational modification of CAVIN1 on its role in asthma, with dephosphorylated CAVIN1 seemingly controlling the release of IL-33 and Th2 response in the challenge phase of mouse asthma model[200].

**Locus 17q25.3**

The 3 Mbp region around the lead SNP encompassed 41 genes which were included in our analysis (Dataset S4). The lead SNP is positioned within an intron of *PGS1,* the top prioritised gene in ADGAPP (score=205, 46% of top 10 total score; Table 1), followed by the gene’s neighbours: *DNAH17* (score=73) and *SOCS3* (score=52) which contributed 16% and 12% to the top 10 total score in the ADGAPP ranking. Colocalisation with *PGS1* (Phosphatidylglycerophosphate synthase 1) eQTLs was detected in the skin (PPH4 = 95%) and fibroblasts (PPH4 = 98%) in GTEx, and suggestively so in LCL (PPH4 = 69%) in the TwinsUK cohort (Additional File 5: Supplementary Table 1), with the AD risk allele correlating with reduced expression. In addition, 385 significant eQTLs in the locus were also found by individual variant lookups (Dataset S5). The two other top gene hits are differentially expressed in eczema patients: *DNAH17* (dynein axonemal heavy chain 17) is 2-6x downregulated[17,49] and *SOCS3* (suppressor of cytokine signaling 3) upregulated[16]. While there are 110 eQTLs associated with *DNAH17* in the region[31,34,36], only 27[30,67] were detected for *SOCS3* despite chromatin conformation capture evidence for interactions between variants in the region and gene’s promoter[5,41,91], and numerous overlapping mQTLs.

The EAGLE GWAS annotation[3] postulated that *SOCS3* is the causal gene at the locus due to MAGENTA gene set enrichment results, the gene’s regulation of cytokine signalling, case-control SNP haplotype association and differential expression in AD.

Despite eQTL-GWAS colocalisation signal observed for *PGS1*, any possible link to AD is not immediately apparent for this enzyme catalysing the first step in production of cardiolipin [205], an integral lipid component of the inner mitochondrial membrane[206]. However, anticardiolipin antibodies are present in child patients with extrinsic AD[207,208]. The antibodies are reported also commonly in autoimmune diseases, e.g. in antiphospholipid syndrome (APS) or SLE. *DNAH17* is predicted to encode a chain of dynein motor protein implicated in cilia assembly and motility[209].

Only the third-ranked gene – *SOCS3*, despite low evidence for prioritisation, appears to be directly involved in AD-related cellular processes. SOCS3 exerts a significant negative regulatory role in IL-1, IL-6, IL-12 and IL-23 cytokine signal transduction, among other regulatory functions relating to cytokines, hormones and growth factors[210]. Loss of SOCS3 has severe consequences, with deletion resulting in embryonic lethality. Increased *SOCS3* expression in the gut has been observed in IBD patients where it correlated with inflammation severity[211,212]. Similarly, for asthma and atopic dermatitis, *SOCS3* expression in CD3+ T cells positively correlates with disease pathology. Constitutive expression of *Socs3* in murine T cells increased Th2 skewing and enhanced airway hypersensitivity[213], and the reverse in conditional knockout mutants in T cells[214]. Keratinocyte-specific knockout of *Socs3* results in abnormal wound healing characterised by hyperproliferative epidermis and neutrophil infiltration[215]

**Locus 19p13.2**

ADGAPP provides limited evidence for the top 3 prioritised genes at this locus, out of the 30 genes positioned within the 3 Mbp window around index SNP and included in the analysis (Dataset S4). The intergenic index SNP at the locus is situated between *ACTL9* (ranked 1^st^, score 115 and 41% of top 10 cumulative total; Table 1) and *ADAMTS10* (ranked 2^nd^, score 57 and 20% of top 10 cumulative total). The third ranked MAP2K7 scored only 34, 12% of top 10 cumulative total. While *ACTL9* (actin like 9) is promoted as the top hit in ADGAPP, it is only due to numerous promoter-enhancer interactions[41,91,131] and mQTLs linked to this gene (Dataset S5). Second ranked *ADAMTS10* (ADAM metallopeptidase with thrombospondin type 1 motif 10) is downregulated in the skin of homozygous *FLG* mutation AD patients[17] and individual look-ups show 37 eQTL hits in the locus interval in whole blood, monocytes and T cells[30,34,36,216] . Third ranked gene, *MAP2K7* (mitogen-activated protein kinase 7) is the only gene with colocalisation evidence at the locus (Additional File 5: Supplementary Table 1). Posterior probability of colocalisation is 90% in CD4+ T cells in CEDAR with risk allele associated with reduction in expression, and the gene is also downregulated in homozygous and heterozygous *FLG* mutation AD patients[17], as well as on differentiation from epidermal stem cells to keratinocytes[5] (Dataset S5). No clear candidate gene was presented in the Paternoster et al. (2015) AD GWAS annotation[3].

*ACTL9* encodes an uncharacterised protein from the family of actin-related proteins. *ADAMTS10* belongs to a family of 19 secreted zinc-dependent metalloproteinases that participate in connective tissue remodelling. Murine knockouts in the gene show multiple abnormalities, relating to ciliary body, epidermal-dermal junction and skeletal muscle morphology [88,217]. In humans, mutations in the gene have been mapped to autosomal recessive Weill-Marchesani Syndrome[218], characterised by generalized mesodermal dysplasia affecting bones, eyes and resulting in pachyderma[219]. Molecular characterisation revealed that ADAMTS10 specifically helps in formation of fibrillin microfibrils in the extracellular matrix[220] and thus promotes focal adhesions and epithelial cell-cell junction formation[221].

MAP2K7 is a kinase in the environmental stress-activated protein kinase/c-Jun N-terminal kinases (SAP/JNK) signalling pathway and relays signal in the JNK pathway triggered by proinflammatory cytokines, specifically IL1-α[222,223], crucial in skin inflammation and whose expression positively correlates with AD symptoms[222]. In dendritic cells within tumour microenvironment, MAP2K7 is positioned downstream of TGF-β[224], which upregulates the expression of miR-27a that silences, among others, *MAP2K7*. This results in inhibition in expression of key pro-inflammatory cytokines and in parallel accumulation of T regulatory cells along with reduction in differentiation to Th1 and Th17 cells, suggesting dual role of JNK pathway kinases in regulating cytokine expression; however, the Treg/Th17 balance observed in *MAP2K7* knockouts is the direct opposite of the one seen in AD[225,226].

**Locus 20q13.33**

At this locus, we tested 109 potential candidate genes situated within the 3 Mbp interval around index SNP (Dataset S4). The three top-ranked genes are novel candidates which show consistent support in independent eQTL colocalisation analyses in whole blood, immune cell types and skin (Additional File 5: Supplementary Table 1): *STMN3* (score=608, 27% of top 10 cumulative total; Table 1), *LIME1* (score=473, 21% of top 10 cumulative total)*,* and *ARFRP1* (score=257, 12% of top 10 cumulative total).

*STMN3* (stathmin 3) eQTLs from sun-exposed/unexposed skin (PPH4 = 99.2% in coloc and *p*-value of 2 x 10^-8^ in TWAS; protective allele correlating with increased expression; Additional File 5: Supplementary Table 1) and whole blood in GTEx (PPH4 = 98%, reduced expression), and Blueprint’s monocytes (PPH4 = 91%, reduced expression) show strong evidence of colocalisation with AD GWAS, and TwinsUK’s skin shows slightly weaker evidence of colocalisation (PPH4 = 87%, increased expression). In a similar way to *STMN3*, eQTLs in *LIME1* (Lck-interacting transmembrane adaptor 1) colocalise in GTEx sun-exposed/unexposed skin (PPH4 = 87% and PPH4 = 90%, protective allele correlating with increased expression), whole blood (PPH4 = 91%, increased expression), CEDAR CD4+ T cells (PPH4 = 92%, reduced expression), TwinsUK skin (PPH4 = 99.2%, increased expression) and eQTLGen whole blood (PPH4 = 89%, increased expression). Third ranked, *ARFRP1* (ADP ribosylation factor related protein 1) shows good evidence for colocalisation in GTEx: whole blood (PPH4 = 88%, protective allele correlates with increased expression), fibroblasts (PPH4 = 83%, reduced expression) and in Blueprint neutrophils (PPH4 = 91%, increased expression).

This is the only locus, where we detected pQTL colocalisation in the Sun *et al*. (2018) dataset – however, the posterior probability of colocalisation for *TNFRSF6B* (TNF receptor superfamily member 6b, ranked 5^th^) was moderate at 72% (Additional File 5: Supplementary Table 1). The protective allele correlates with increased abundance of the protein product DcR3. Four additional genes showed eQTL colocalisation in single tissues: *GMEB2* (glucocorticoid modulatory element binding protein 2) – ranked 4^th^, *SLC2A4RG* (SLC2A4 regulator) – ranked 7^th^, *UCKL1* (uridine-cytidine kinase 1 like 1) – ranked 13^th^ and *RBBP8NL* (RBBP8 N-terminal like) – ranked 16^th^. Complex pattern of gene co-regulation at the locus does not make it easy to distil the likely GWAS target(s) but the three top prioritised genes along with *TNFRSF6B* offer the most ample, albeit differing across tissues, evidence of relationship with the AD protective variant. Similarly, the original GWAS analysis in Paternoster et al. (2015)[3] did not provide a clear candidate gene for the locus, and mentions *TNFRSF6B*, *RTEL1*, *LIME1*, *SLC2A4RG* as potential targets.

STMN3 is a member of the stathmin-like family of proteins, which are involved in regulating microtubule dynamics and inhibit tubulin polymerization[227]. STMN3, also known as SLIP, is especially intensively expressed in central nervous system and highly proliferative cancer cells, associated with neural cell growth[228], as well as tumour cell survival and migration[228–230]. Its knockdown in breast cancer cells causes loss of epithelial morphology, concomitant with reduced cadherin expression[229].

*LIME1* is a poorly characterised transmembrane adaptor phosphoprotein that relays T cell receptor stimulation to downstream signalling pathways following its phosphorylation by the Src family kinases[231,232]. The protein is highly and specifically expressed in nasal respiratory epithelium, followed by cytotoxic CD8+ T cells, helper CD4+ T cells, B-lymphocytes and NK cells[233] where it moves to immunological synapse on contact with antigen-presenting cells and is later downregulated after T cell activation. Overexpression of the gene in T cells induces *IL-2* promoter activity[231], which connects the gene more closely to AD phenotype, because of the importance of Th2 response in AD development, further underlined by possible direct role for interleukin-2 (AD GWAS locus 4q27) and/or its receptor (AD GWAS locus 10p15.1) in mediating AD risk.

ARFRP1 is a small GTP-ase which is associated with the *trans*-Golgi, where it regulates intracellular protein trafficking, such as of E-catherin through the Golgi to plasma membrane and in particular, plays a key role in lipoprotein maturation and formation of lipid droplets in adipocytes[234–237].

*TNFRSF6B* encodes a decoy protein DcR3 which binds to FAS ligand and LIGHT and neutralizes their action. Binding of DcR3 to FAS ligand, which is produced by activated T cells and NK cells and promotes cytotoxic killing of cells, antagonizes apoptosis of such cells[238]. In addition to inducing apoptosis, LIGHT promotes T cell activation and regulates airway modelling in asthma patients[239,240]. Independently of action on FASL and LIGHT, TNFRSF6B is thought to positively regulate dendritic cell differentiation and Th2 polarization. TNFRSF6B serum concentration is increased in inflammatory disease, including allergic disease: atopic dermatitis and asthma[239,241–243], which does not agree with the direction of effect seen in pQTLs, whereby a protective allele would lead to higher protein expression. A synonymous variant in the gene has been previously reported as a top SNP in another AD GWAS[243].

60. Gupta S, Deepti A, Deegan S, Lisbona F, Hetz C, Samali A. HSP72 protects cells from ER stress-induced apoptosis via enhancement of IRE1α-xbp1 signaling through a physical interaction. PLoS Biol. 2010;8.

100. Hirano A, Goto M, Mitsui T, Hashimoto-Hachiya A, Tsuji G, Furue M. Antioxidant Artemisia princeps Extract Enhances the Expression of Filaggrin and Loricrin via the AHR/OVOL1 Pathway. Int J Mol Sci. 2017;18.

117. Harmston N, Ing-Simmons E, Tan G, Perry M, Merkenschlager M, Lenhard B. Topologically associating domains are ancient features that coincide with Metazoan clusters of extreme noncoding conservation. Nat Commun. Springer US; 2017;8.

136. Kim JE, Kim JS, Cho DH, Park HJ. Molecular mechanisms of cutaneous inflammatory disorder: Atopic dermatitis. Int J Mol Sci. 2016;17.

193. Amano W, Nakajima S, Kunugi H, Numata Y. Atopic dermatitis and skin disease The Janus kinase inhibitor JTE-052 improves skin barrier function through suppressing signal transducer and activator of transcription 3 signaling. J Allergy Clin Immunol. Elsevier Ltd; 136:667-677.e7.

206. Shen Z, Ye C, Mccain K, Greenberg ML. The Role of Cardiolipin in Cardiovascular Health. Biomed Res Int. 2015;2015.

241. Kowal K, Pampuch A, Sacharzewska E, Kowal-Bielecka O. Serum concentration of TNFSF14/LIGHT receptors DcR3 and HVEM in asthma patients. J Allergy Clin Immunol. Elsevier Limited; 2019;143:AB8.

242. Chen CC, Yang YH, Lin YT, Hsieh SL, Chiang B-L. Soluble decoy receptor 3: increased levels in atopic patients. J Allergy Clin Immunol. Elsevier Limited; 2004;114:195.

243. Ellinghaus D, Baurecht H, Esparza-Gordillo J, Rodríguez E, Matanovic A, Marenholz I, et al. High-density genotyping study identifies four new susceptibility loci for atopic dermatitis. Nat Genet. 2013;45:808–12.
