## Additional File 8 Supplementary Figure 4 for "Triangulating molecular evidence to prioritise candidate causal genes at established atopic dermatitis loci"

| | coloc | TWAS | Hi−C | eQTL | DGE | regfm | PrixFixe | mQTL | pQTL | hQTL |
|---|---|---|---|---|---|---|---|---|---|---|
| 1q21.3a / rs61813875 HRNR | | | 334 | 79 | | | | 4 | | |
| 1q21.3a / rs61813875 RPTN | | | 279 | 0 | 4 | | | 2 | | |
| 1q21.3a / rs61813875 CRNN | | 38 | 206 | 0 | 4 | | | 0 | | |
| 1q21.3b / rs12730935 IL6R | 320 | 70 | 83 | 109 | 19 | | | 6 | 52 | |
| 1q21.3b / rs12730935 UBE2Q1 | | | 22 | 46 | 8 | | | 6 | | |
| 1q21.3b / rs12730935 ADAR | | | 7 | 39 | 9 | | | 5 | | |
| 2p13.3 / rs112111458 CD207 | 115 | 76 | 28 | 21 | | | | 5 | | |
| 2p13.3 / rs112111458 CLEC4F | | | | 60 | | | | 2 | | |
| 2p13.3 / rs112111458 VAX2 | | | 41 | | 4 | | | 9 | | |
| 2q12.1 / rs6419573 IL18R1 | 265 | 77 | 22 | 900 | 14 | | | 37 | 3 | |
| 2q12.1 / rs6419573 IL18RAP | 215 | 186 | 41 | 685 | 18 | | | 26 | 4 | |
| 2q12.1 / rs6419573 IL1RL1 | 73 | | 33 | 39 | 10 | | | 21 | 28 | |
| 2q37.1 / rs1057258 INPP5D | 60 | | 16 | 157 | 16 | | 5 | 8 | | |
| 2q37.1 / rs1057258 ATG16L1 | | | 72 | 1 | 13 | | 3 | 14 | | |
| 2q37.1 / rs1057258 RN7SL32P | | | | 25 | | | | | | |
| 4q27 / rs6827756 KIAA1109 | | | 43 | 152 | 4 | | 4 | 6 | | |
| 4q27 / rs6827756 BBS12 | 78 | | 8 | 19 | 3 | | | 0 | | |
| 4q27 / rs6827756 TRPC3 | 57 | | 21 | 8 | 3 | | | 7 | | |
| 5p13.2 / rs10214237 IL7R | | | 299 | 447 | 16 | | 6 | 19 | 35 | |
| 5p13.2 / rs10214237 SPEF2 | | | 158 | 7 | 12 | | 3 | 10 | | |
| 5p13.2 / rs10214237 UGT3A2 | 19 | | 2 | 8 | 9 | 48 | | 0 | | |
| 5q31.1a / rs12188917 SLC22A5 | | 57 | | 355 | 20 | | | 4 | | |
| 5q31.1a / rs12188917 IRF1 | | | 46 | 116 | 39 | | | 11 | | |
| 5q31.1a / rs12188917 RAD50 | 38 | 34 | 7 | 20 | 15 | | | | | |
| 5q31.1b / rs4705962 KIF3A | 61 | | 9 | 143 | 9 | | | 4 | | |
| 5q31.1b / rs4705962 SLC22A5 | 71 | | | 28 | 17 | 115 | | 3 | | |
| 5q31.1b / rs4705962 PDLIM4 | 71 | | 6 | 55 | | | | 4 | | |
| 6p21.32 / rs4713555 HLA−DRA | 13 | | 1186 | 63 | 15 | | | 5 | | 4 |
| 6p21.32 / rs4713555 HLA−DQB1 | | | 112 | 422 | 23 | | | 6 | | 10 |
| 6p21.32 / rs4713555 HLA−DRB1 | | | 65 | 389 | 5 | | | 5 | | 6 |
| 6p21.33 / rs41293864 HSPA1B | 114 | 28 | 1 | 8 | 10 | | | 3 | 0 | 0 |
| 6p21.33 / rs41293864 HCG27 | | | 19 | 107 | 12 | | | 4 | | 5 |
| 6p21.33 / rs41293864 CSNK2B | 51 | 65 | 4 | 7 | 20 | | | 1 | 0 | 0 |
| 8q21.13 / rs6473227 ZBTB10 | 66 | | 10 | 31 | 10 | 58 | 4 | 4 | | |
| 8q21.13 / rs6473227 TPD52 | | | 31 | 11 | 11 | | | 12 | | |
| 8q21.13 / rs6473227 PAG1 | | | 6 | 3 | 15 | 42 | | | | |
| 10p15.1 / rs6602364 IL2RA | 103 | | 118 | 60 | 7 | | 5 | 18 | | |
| 10p15.1 / rs6602364 RBM17 | 23 | 24 | 3 | 29 | 5 | | 3 | 16 | | |
| 10p15.1 / rs6602364 PFKFB3 | | | 11 | 9 | 8 | | | 17 | | |
| 10q21.2 / rs2944542 ADO | | | 132 | 406 | 11 | | | 32 | | |
| 10q21.2 / rs2944542 ZNF365 | | | 54 | 13 | 9 | | 4 | 17 | | |
| 10q21.2 / rs2944542 EGR2 | | | 50 | 6 | 10 | | | 17 | | |
| 11p13 / rs2592555 PRR5L | 351 | | 34 | 131 | 10 | | 4 | 33 | | |
| 11p13 / rs2592555 TRAF6 | 40 | | 9 | 4 | 7 | | | 2 | | |
| 11p13 / rs2592555 COMMD9 | | | 9 | 1 | 7 | | 2 | 11 | | |
| 11q13.1 / rs10791824 CTSW | | | 6 | 279 | 22 | | 6 | 3 | | |
| 11q13.1 / rs10791824 OVOL1 | 115 | 67 | 10 | 15 | 5 | | 4 | 3 | | 11 |
| 11q13.1 / rs10791824 EFEMP2 | 9 | 26 | 13 | 76 | 27 | | | 3 | | 2 |
| 11q13.5 / rs2212434 LRRC32 | 102 | | 383 | 22 | 4 | | | 18 | | |
| 11q13.5 / rs2212434 EMSY | 72 | | 232 | 94 | 14 | | | 29 | | |
| 11q13.5 / rs2212434 THAP12 | | | 11 | 14 | 6 | | | 14 | | |
| 11q24.3 / rs7127307 ETS1 | 110 | | 24 | 35 | 26 | | | 78 | | |
| 11q24.3 / rs7127307 FLII | 35 | | | | | | | | | |
| 11q24.3 / rs7127307 APLP2 | | | 5 | 4 | 9 | | | | | |
| 12q15 / rs2227483 MDM1 | | | 47 | 532 | 9 | | | 55 | | |
| 12q15 / rs2227483 IL22 | | | 51 | 6 | 7 | | 3 | 29 | | |
| 12q15 / rs2227483 IFNG | 40 | | 6 | 0 | 5 | | 5 | | | |
| 14q13.2 / rs2038255 PPP2R3C | 277 | 104 | 18 | 485 | 28 | | | | 3 | 3 |
| 14q13.2 / rs2038255 KIAA0391 | 309 | 195 | 27 | 223 | 13 | | | 10 | | 2 |
| 14q13.2 / rs2038255 SRP54 | | | 60 | 307 | 35 | | | 1 | | 2 |
| 14q32.32 / rs7146581 TRAF3 | 136 | | 364 | 253 | 10 | | 6 | 14 | | |
| 14q32.32 / rs7146581 AMN | | | 224 | 24 | 3 | | 3 | 10 | | |
| 14q32.32 / rs7146581 CDC42BPB | | | 15 | 132 | 9 | | 4 | 13 | | |
| 16p13.13 / rs2041733 DEXI | | 29 | 55 | 213 | 9 | | | 15 | | |
| 16p13.13 / rs2041733 CLEC16A | 29 | | 129 | 89 | 24 | | 4 | 31 | | 21 |
| 16p13.13 / rs2041733 RMI2 | | | 18 | 42 | 9 | | | 21 | | 10 |
| 17q21.2 / rs12951971 DHX58 | | | 3 | 216 | 11 | | | 4 | | |
| 17q21.2 / rs12951971 STAT3 | 45 | | 9 | 27 | 5 | | | 4 | 5 | |
| 17q21.2 / rs12951971 RAB5C | | | 4 | 62 | 4 | | | 1 | | |
| 17q25.3 / rs11657987 PGS1 | | | 42 | 69 | 3 | | | 35 | | |
| 17q25.3 / rs11657987 DNAH17 | | | 2 | 25 | 11 | | | 27 | | |
| 17q25.3 / rs11657987 SOCS3 | | | 11 | 3 | 15 | | | 19 | | |
| 19p13.2 / rs2918307 ACTL9 | | | 98 | | | | | 16 | | |
| 19p13.2 / rs2918307 ADAMTS10 | | | 1 | 38 | 8 | | | 3 | | |
| 19p13.2 / rs2918307 MAP2K7 | 15 | | | 2 | 13 | | | | | |
| 20q13.33 / rs4809219 STMN3 | 171 | 16 | | 342 | 13 | | | 7 | | |
| 20q13.33 / rs4809219 LIME1 | 246 | 112 | 11 | 59 | 10 | | | 5 | | |
| 20q13.33 / rs4809219 ARFRP1 | 109 | 12 | 1 | 32 | 4 | | | | | |

score: 1100, 1000, 900, 800, 700, 600, 500, 400, 300, 200, 100, 0
