## Additional File 9 Supplementary File for "Triangulating molecular evidence to prioritise candidate causal genes at established atopic dermatitis loci"

**Supplementary Table 3**

*Genes frequently associated with atopic dermatitis, and not prioritised as our top candidate genes*(1–4)*.*

TSLP

TSLPR

TLR2

IFNGR1

SPINK5

SPRR3

FLG

FLG2

FCER1G

FCER1A

IL4

IL31

IVL

LOR

CLDN1

DSG1

TMEM79

STAT6

1.	Hoffjan S, Stemmler S. Unravelling the complex genetic background of atopic dermatitis : from genetic association results towards novel therapeutic strategies. Arch Dermatol Res [Internet]. 2015;659–70. Available from: http://dx.doi.org/10.1007/s00403-015-1550-6

2.	Bin L, Leung DYMM. Genetic and epigenetic studies of atopic dermatitis. Allergy, Asthma Clin Immunol. 2016;12(1):1–14.

3.	Kim JEJS, Kim JEJS, Cho DH, Park HJ. Molecular mechanisms of cutaneous inflammatory disorder: Atopic dermatitis. Int J Mol Sci. 2016;17(8).

4.	Liang Y, Chang C, Lu Q. The Genetics and Epigenetics of Atopic Dermatitis—Filaggrin and Other Polymorphisms. Clin Rev Allergy Immunol [Internet]. 2016;51(3):315–28. Available from: http://dx.doi.org/10.1007/s12016-015-8508-5
