## Supplementary material for "Triangulating molecular evidence to prioritise candidate causal genes at established atopic dermatitis loci": Consortium members

eQTLGen Consortium – Author information

Mawussé Agbessi^1^, Habibul Ahsan^2^, Isabel Alves^1^, Anand Kumar Andiappan^3^, Wibowo Arindrarto^4^, Philip Awadalla^1^, Alexis Battle^5,6^, Frank Beutner^7^, Marc Jan Bonder^8,9^, Dorret I. Boomsma^10^, Mark W. Christiansen^11^, Annique Claringbould^8,12^, Patrick Deelen^8,13,12,14^, Tõnu Esko^15^, Marie-Julie Favé^1^, Lude Franke^8,12^, Timothy Frayling^16^, Sina A. Gharib^11,17^, Greg Gibson^18^, Bastiaan T. Heijmans^4^, Gibran Hemani^19^, Rick Jansen^20^, Mika Kähönen^21^, Anette Kalnapenkis^15^, Silva Kasela^15^, Johannes Kettunen^22^, Yungil Kim^23,5^, Holger Kirsten^24^, Peter Kovacs^25^, Knut Krohn^26^, Jaanika Kronberg^15^, Viktorija Kukushkina^15^, Zoltan Kutalik^27^, Bernett Lee^3^, Terho Lehtimäki^28^, Markus Loeffler^24^, Urko M. Marigorta^18,29,30^, Hailang Mei^31^, Lili Milani^15^, Grant W. Montgomery^32^, Martina Müller-Nurasyid^33,34,35^, Matthias Nauck^36,37^, Michel G. Nivard^38^, Brenda Penninx^20^, Markus Perola^39^, Natalia Pervjakova^15^, Brandon L. Pierce^2^, Joseph Powell^40^, Holger Prokisch^41,42^, Bruce M. Psaty^11,43^, Olli T. Raitakari^44^, Samuli Ripatti^45^, Olaf Rotzschke^3^, Sina Rüeger^27^, Ashis Saha^5^, Markus Scholz^24^, Katharina Schramm^46,34^, Ilkka Seppälä^28^, Eline P. Slagboom^4^, Coen D.A. Stehouwer^47^, Michael Stumvoll^48^, Patrick Sullivan^49^, Peter A.C. ‘t Hoen^50^, Alexander Teumer^51^, Joachim Thiery^52^, Lin Tong^2^, Anke Tönjes^48^, Jenny van Dongen^10^, Maarten van Iterson^4^, Joyce van Meurs^53^, Jan H. Veldink^54^, Joost Verlouw^53^, Peter M. Visscher^32^, Uwe Völker^55^, Urmo Võsa^8,15^, Harm-Jan Westra^8,12^, Cisca Wijmenga^8^, Hanieh Yaghootkar^16,56,57^, Jian Yang^32,58^, Biao Zeng^18^, Futao Zhang^32^

Author list is ordered alphabetically

1. Computational Biology, Ontario Institute for Cancer Research, Toronto, Canada

2. Department of Public Health Sciences, University of Chicago, Chicago, United States of America

3. Singapore Immunology Network, Agency for Science, Technology and Research, Singapore, Singapore

4. Leiden University Medical Center, Leiden, The Netherlands

5. Department of Computer Science, Johns Hopkins University, Baltimore, United States of America

6. Departments of Biomedical Engineering, Johns Hopkins University, Baltimore, United States of America

7. Heart Center Leipzig, Universität Leipzig, Leipzig, Germany

8. Department of Genetics, University Medical Centre Groningen, Groningen, The Netherlands

9. European Molecular Biology Laboratory, Genome Biology Unit, 69117 Heidelberg, Germany

10. Netherlands Twin Register, Department of Biological Psychology, Vrije Universiteit Amsterdam, Amsterdam Public Health research institute and Amsterdam Neuroscience, the Netherlands

11. Cardiovascular Health Research Unit, University of Washington, Seattle, United States of America

12. Oncode Institute

13. Genomics Coordination Center, University Medical Centre Groningen, Groningen, The Netherlands

14. Department of Genetics, University Medical Centre Utrecht, P.O. Box 85500, 3508 GA, Utrecht, The Netherlands

15. Estonian Genome Center, Institute of Genomics, University of Tartu, Tartu 51010, Estonia

16. Genetics of Complex Traits, University of Exeter Medical School, Royal Devon & Exeter Hospital, Exeter, United Kingdom

17. Department of Medicine, University of Washington, Seattle, United States of America

18. School of Biological Sciences, Georgia Tech, Atlanta, United States of America

19. MRC Integrative Epidemiology Unit, University of Bristol, Bristol, United Kingdom

20. Amsterdam UMC, Vrije Universiteit, Department of Psychiatry, Amsterdam Public Health research institute and Amsterdam Neuroscience, The Netherlands

21. Department of Clinical Physiology, Tampere University Hospital and Faculty of Medicine and Health Technology, Tampere University, Tampere, Finland

22. University of Helsinki, Helsinki, Finland

23. Genetics and Genomic Science Department, Icahn School of Medicine at Mount Sinai, New York, United States of America

24. Institut für Medizinische InformatiK, Statistik und Epidemiologie, LIFE – Leipzig Research Center for Civilization Diseases, Universität Leipzig, Leipzig, Germany

25. IFB Adiposity Diseases, Universität Leipzig, Leipzig, Germany

26. Interdisciplinary Center for Clinical Research, Faculty of Medicine, Universität Leipzig, Leipzig, Germany

27. Lausanne University Hospital, Lausanne, Switzerland

28. Department of Clinical Chemistry, Fimlab Laboratories and Finnish Cardiovascular Research Center-Tampere, Faculty of Medicine and Health Technology, Tampere University, Tampere, Finland

29. Integrative Genomics Lab, CIC bioGUNE, Bizkaia Science and Technology Park, Derio, Bizkaia, Basque Country, Spain

30. IKERBASQUE, Basque Foundation for Science, Bilbao, Spain

31. Department of Medical Statistics and Bioinformatics, Leiden University Medical Center, Leiden, The Netherlands

32. Institute for Molecular Bioscience, University of Queensland, Brisbane, Australia

33. Institute of Genetic Epidemiology, Helmholtz Zentrum München - German Research Center for Environmental Health, Neuherberg, Germany

34. Department of Medicine I, University Hospital Munich, Ludwig Maximilian’s University, München, Germany

35. DZHK (German Centre for Cardiovascular Research), partner site Munich Heart Alliance, Munich, Germany

36. Institute of Clinical Chemistry and Laboratory Medicine, Greifswald University Hospital, Greifswald, Germany

37. German Center for Cardiovascular Research (partner site Greifswald), Greifswald, Germany

38. Department of Biological Psychology, Faculty of Behaviour and Movement Sciences, VU, Amsterdam, The Netherlands

39. National Institute for Health and Welfare, University of Helsinki, Helsinki, Finland

40. Garvan Institute of Medical Research, Garvan-Weizmann Centre for Cellular Genomics, Sydney, Australia

41. Institute of Human Genetics, Helmholtz Zentrum München, Neuherberg, Germany

42. Institute of Human Genetics, Technical University Munich, Munich, Germany

43. Kaiser Permanente Washington Health Research Institute, Seattle, WA, United States of America

44. Centre for Population Health Research, Department of Clinical Physiology and Nuclear Medicine, Turku University Hospital and University of Turku, Turku, Finland

45. Statistical and Translational Genetics, University of Helsinki, Helsinki, Finland

46. Institute of Genetic Epidemiology, Helmholtz Zentrum München - German Research Center for Environmental Health, Neuherberg, Germany

47. Department of Internal Medicine and School for Cardiovascular Diseases (CARIM), Maastricht University Medical Center, Maastricht, The Netherlands

48. Department of Medicine, Universität Leipzig, Leipzig, Germany

49. Department of Medical Epidemiology and Biostatistics, Karolinska Institutet, Stockholm, Sweden

50. Center for Molecular and Biomolecular Informatics, Radboud Institute for Molecular Life Sciences, Radboud University Medical Center Nijmegen, Nijmegen, The Netherlands

51. Institute of Clinical Chemistry and Laboratory Medicine, University Medicine Greifswald, Greifswald, Germany

52. Institute for Laboratory Medicine, LIFE – Leipzig Research Center for Civilization Diseases, Universität Leipzig, Leipzig, Germany

53. Department of Internal Medicine, Erasmus Medical Centre, Rotterdam, The Netherlands

54. UMC Utrecht Brain Center, University Medical Center Utrecht, Department of Neurology, Utrecht University, Utrecht, The Netherlands

55. Interfaculty Institute for Genetics and Functional Genomics, University Medicine Greifswald, Greifswald, Germany

56. School of Life Sciences, College of Liberal Arts and Science, University of Westminster, 115 New Cavendish Street, London, United Kingdom

57. Division of Medical Sciences, Department of Health Sciences, Luleå University of Technology, Luleå, Sweden

58. Institute for Advanced Research, Wenzhou Medical University, Wenzhou, Zhejiang 325027, China

**BIOS Consortium (Biobank-based Integrative Omics Study) – Author information**

**Management Team** Bastiaan T. Heijmans (chair)^1^, Peter A.C. ’t Hoen^2^, Joyce van Meurs^3^, Aaron Isaacs^4^, Rick Jansen^5^, Lude Franke^6^.
Cohort collection Dorret I. Boomsma^7^, René Pool^7^, Jenny van Dongen^7^, Jouke J. Hottenga^7^ (Netherlands Twin Register); Marleen MJ van Greevenbroek^8^, Coen D.A. Stehouwer^8^, Carla J.H. van der Kallen^8^, Casper G. Schalkwijk^8^ (Cohort study on Diabetes and Atherosclerosis Maastricht); Cisca Wijmenga^6^, Lude Franke^6^, Sasha Zhernakova^6^, Ettje F. Tigchelaar^6^ (LifeLines Deep); P. Eline Slagboom^1^, Marian Beekman^1^, Joris Deelen^1^, Diana van Heemst^9^ (Leiden Longevity Study); Jan H. Veldink^10^, Leonard H. van den Berg^10^ (Prospective ALS Study Netherlands); Cornelia M. van Duijn^4^, Bert A. Hofman^11^, Aaron Isaacs^4^, André G. Uitterlinden^3^ (Rotterdam Study).
Data Generation Joyce van Meurs (Chair)^3^, P. Mila Jhamai^3^, Michael Verbiest^3^, H. Eka D.
Suchiman^1^, Marijn Verkerk^3^, Ruud van der Breggen^1^, Jeroen van Rooij^3^, Nico Lakenberg^1^.

**Data management and computational infrastructure** Hailiang Mei (Chair)^12^, Maarten van Iterson^1^, Michiel van Galen^2^, Jan Bot^13^, Dasha V. Zhernakova^6^, Rick Jansen^5^, Peter van ’t Hof^12^, Patrick Deelen^6^, Irene Nooren^13^, Peter A.C. ’t Hoen^2^, Bastiaan T. Heijmans^1^, Matthijs Moed^1^.
Data Analysis Group Lude Franke (Co-Chair)^6^, Martijn Vermaat^2^, Dasha V. Zhernakova^6^, René Luijk^1^, Marc Jan Bonder^6^, Maarten van Iterson^1^, Patrick Deelen^6^, Freerk van Dijk^14^, Michiel van Galen^2^, Wibowo Arindrarto^12^, Szymon M. Kielbasa^15^, Morris A. Swertz^14^, Erik. W van Zwet^15^, Rick Jansen^5^, Peter-Bram ’t Hoen (Co-Chair)^2^, Bastiaan T. Heijmans (Co-Chair)^1^.

1. Molecular Epidemiology Section, Department of Medical Statistics and Bioinformatics, Leiden University Medical Center, Leiden, The Netherlands
2. Department of Human Genetics, Leiden University Medical Center, Leiden, The Netherlands
3. Department of Internal Medicine, ErasmusMC, Rotterdam, The Netherlands
4. Department of Genetic Epidemiology, ErasmusMC, Rotterdam, The Netherlands
5. Department of Psychiatry, VU University Medical Center, Neuroscience Campus Amsterdam, Amsterdam, The Netherlands
6. Department of Genetics, University of Groningen, University Medical Centre Groningen,
Groningen, The Netherlands
7. Department of Biological Psychology, VU University Amsterdam, Neuroscience Campus
Amsterdam, Amsterdam, The Netherlands
8. Department of Internal Medicine and School for Cardiovascular Diseases (CARIM), Maastricht University Medical Center, Maastricht, The Netherlands
9. Department of Gerontology and Geriatrics, Leiden University Medical Center, Leiden, The
Netherlands
10. Department of Neurology, Brain Center Rudolf Magnus, University Medical Center Utrecht, Utrecht, The Netherlands
11. Department of Epidemiology, ErasmusMC, Rotterdam, The Netherlands
12. Sequence Analysis Support Core, Leiden University Medical Center, Leiden, The Netherlands
13. SURFsara, Amsterdam, the Netherlands
14. Genomics Coordination Center, University Medical Center Groningen, University of Groningen, Groningen, the Netherlands
15. Medical Statistics Section, Department of Medical Statistics and Bioinformatics, Leiden University Medical Center, Leiden, The Netherlands
